# A Genomic Risk Score Identifies Individuals at High Risk for Intracerebral Hemorrhage

**DOI:** 10.1101/2022.05.05.22274399

**Authors:** Evangelos Pavlos Myserlis, Marios K. Georgakis, Stacie L. Demel, Padmini Sekar, Jaeyoon Chung, Rainer Malik, Hyacinth I. Hyacinth, Mary E. Comeau, Guido Falcone, Carl D. Langefeld, Jonathan Rosand, Daniel Woo, Christopher D. Anderson

## Abstract

**Background:** Intracerebral hemorrhage (ICH), the most fatal form of stroke, has an estimated heritability of 29%. Applying a meta-scoring approach, we developed a genomic risk score for ICH and determined its predictive power in comparison to standard clinical risk factors.

**Methods:** Using a meta-analytic approach, we combined genome-wide association data from individuals of European ancestry for ICH and ICH-related traits in a meta-genomic risk score (metaGRS) consisting of 2.6 million variants. We tested associations with ICH and the predictive performance of the metaGRS in addition to clinical risk factors in a held-out validation dataset (842 cases and 796 controls). Furthermore, we tested associations with risk of incident ICH in the population-based UK Biobank cohort (486,784 individuals, 1,526 events, median follow-up 11.3 years).

**Results:** One SD increment in the metaGRS was significantly associated with 45% higher odds for ICH (OR 1.45; 95%CI: 1.30-1.63) in age- and sex-adjusted models and 31% higher odds for ICH (OR: 1.31, 95%CI: 1.16-1.48) in models further adjusted for clinical risk factors. The metaGRS identified individuals with almost 5-fold higher odds for ICH in the top score percentile (OR: 4.83, 95%CI: 1.56-21.2). Predictive models for ICH incorporating the metaGRS in addition to clinical predictors showed superior performance compared with clinical risk factors alone (c-index: 0.695 vs. 0.686). The metaGRS showed similar associations for both lobar and non-lobar ICH, which were independent of the known *APOE* risk locus for lobar ICH. In the UK Biobank, the metaGRS was associated with higher risk of incident ICH (HR: 1.15, 95%CI: 1.09-1.21). The associations were significant within both a relatively high-risk population of users of antithrombotic medications, as well as among a relatively low-risk population with a good control of vascular risk factors and no use of anticoagulants.

**Conclusions:** We developed and validated a genomic risk score that predicts lifetime risk of ICH beyond established clinical risk factors among individuals of European ancestry. Whether implementation of the score in risk prognostication models for high-risk populations, such as patients under antithrombotic treatment, could improve clinical decision making should be explored in future studies.

## INTRODUCTION

Intracerebral hemorrhage (ICH) is the most devastating type of stroke. Although it accounts for 10-20% of all acute cerebrovascular events, it is responsible for almost 50% of stroke-related morbidity and mortality.^1^ Given the lack of effective treatments and the devastating outcome of ICH, primary and secondary prevention is critical. As with many complex human diseases, ICH risk is comprised of both environmental and genetic factors. Early genome-wide association studies (GWAS) of ICH estimated a heritability of 29% and revealed a polygenic architecture,^2, 3^ whereas large-scale GWAS for established risk factors for ICH, such as hypertension and smoking, have identified hundreds of associated genomic loci that cumulatively explain a large proportion of the variance of these traits.^4–9^

Beyond defining heritability and providing insight into biological mechanisms, GWAS findings have begun to show promise for disease risk prediction. Genomic risk scores (GRS), biomarkers representing the aggregated effect of many genetic variants on a given trait, have been proposed as powerful tools for identifying individuals at high risk for complex traits, with a predictive performance at times comparable to that of rare monogenic mutations.^10^ A GRS for ICH could have clinical utility in decision making algorithms, such as for complementing risk-benefit calculation tools in patients prescribed antithrombotic medications. Despite acknowledged limitations, pertaining mainly to translation of the accuracy of GRS findings from the cohort-to the individual-based level, as well as sex- and ancestry-specific predictive differences,^11, 12^ if constructed according to best practices,^12–15^ an ICH GRS could have important implications for patient selection for clinical trials. However, as opposed to other vascular diseases,^10^ efforts to construct a GRS for ICH lag behind, likely because of the lower statistical power in available datasets due to the relative rarity of the disease.

Recent advances in analytical approaches offer more efficient alternative methods for GRS construction that allow to utilize data from multiple GWAS in order to overcome power limitations of genomic datasets of rare diseases.^16, 17^ Based on the assumption that the majority of genetic variants exert their effects on a given disease by affecting intermediate traits, constructing a GRS based on genomic data of traits in the causal pathway of the disease of interest can improve both power and prediction.^16^ In this study, we sought to investigate whether combining genetic liability for possible ICH risk factors and traits reflecting pathologies underlying ICH into a meta-Genomic Risk Score (metaGRS) could improve our ability to predict ICH events among individuals of European ancestry. To further explore the potential utility of such a genomic score for clinical risk prediction, we also assessed whether the metaGRS improves ICH risk prediction beyond established clinical risk factors.

## METHODS

### Data availability statement

The data that support the findings of this study will be available from the corresponding author upon reasonable request. Single nucleotide polymorphism (SNP)-specific weights for the ICH metaGRS will be made publicly available at The Polygenic Score (PGS) Catalog (https://www.pgscatalog.org).

### Study design and participating studies

As our primary data source, we used genotype and phenotype data from 1,861 ICH cases and 1,722 ICH-free controls from three independent GWAS datasets: the North American (USA) multi-center Genetics of Cerebral Hemorrhage on Anticoagulation (GOCHA) study, a prospectively collected case-control study of European ancestry subjects aged > 55 years with primary ICH^18^; the European member sites contributing ICH cases and controls to the International Stroke Genetics Consortium (EUR/ISGC); and the Genetic and Environmental Risk Factors for Hemorrhagic Stroke (GERFHS) study, a prospectively collected case-control study of subjects > 18 years of age with spontaneous ICH in the Greater Cincinnati region^19^. GOCHA and EUR/ISGC were used as the training datasets, whereas GERFHS was our primary validation dataset. Furthermore, we performed an external validation of the derived score for incident ICH events in the UK Biobank (UKBB) cohort, over a median follow-up of 11.3 years among 486,623 individuals aged 40-69 years at recruitment without a prior history of ICH.^20^ The Institutional Review Boards (IRB) of Massachusetts General Hospital, Mayo Clinic (FL), University of Virginia Health System, University of Florida College of Medicine, University of Michigan Health System, Beth Israel Deaconess Medical Center approved the GOCHA study.^21^ IRBs of 16 hospital systems within a 50-mile radius from the University of Cincinnati approved the GERFHS study.^19^ The information used in the Hospital del Mar Intracerebral Hemorrhage (HM-ICH) study was collected from the prospective clinical protocols of the Hospital del Mar, which fulfilled the local ethical guidelines. The identities of the individual patients were kept anonymous. Therefore, patients signed no specific informed consent.^22^ For the Vall d’Hebron Hospital ICH (VVH-ICH) study, informed written consent was obtained from all subjects, and the local Ethics Committee approved the study.^23^ The ethics committee of the University Hospital, Krakow, Poland approved the Jagiellonian University Hemorrhagic Stroke (JUHS) Study.^24^ The Lund Stroke Register (LSR) study in Lund, Sweden was approved by the Lund University Ethics Committee. Informed consent was obtained from the prospectively included patients (or in some cases from next of kin).^25^

### Ascertainment of cases and controls

Primary ICH cases in all studies were recruited through participating hospitals and were defined as a new and acute neurological deficit with confirmation through computed tomography or magnetic resonance imaging and without evidence of trauma, brain tumor, hemorrhagic transformation of a cerebral infarction, vascular malformation, or any other cause of secondary ICH. ICH-free controls were recruited from the same populations through inpatient recruitment, ambulatory centers in the local communities, and random digit dialing in the same population or population-based cohorts. A detailed description of case and control ascertainment and inclusion and exclusion criteria for the participating studies is described in the **Supplemental Material**. Details about sample recruitment as well as about genotyping and quality control (QC) of the individuals in the three studies have been previously described.^2, 26, 27^ For the purposes of the current study, we excluded patients with missing data on ICH status, age, sex and principal components (PCs) reflecting ancestry. We included only primary ICH cases, after applying previously described methods of enrollment and inclusion/exclusion criteria.^26^

### Imputation of main GWAS datasets

Samples in the training and validation GWAS datasets of ICH were separately imputed using the Haplotype Reference Consortium (HRC) reference panel on the Michigan Imputation Server (https://imputationserver.sph.umich.edu). We retained only SNPs with minor allele frequency (MAF) >0.01 and high imputation quality (R^2^ >0.4). We excluded multi-allelic variants according to previously published methodologies.^3^ In the UKBB, we used the version 3 genotypes, which were genotyped on the UKB Axiom array and imputed to the HRC.^28^ To examine the metaGRS in the UKBB, we considered only genotyped or HRC-imputed SNPs with imputation INFO > 0.01 and MAF >0.001.

### Trait-specific genomic risk scores (GRS) construction

We used GOCHA (436 ICH cases and 405 controls) and EUR/ISGC (577 ICH cases and 523 controls) as training datasets to develop GRS for 21 traits associated with ICH risk. We leveraged publicly available GWAS summary-level data from international consortia, as detailed in **Supplementary Table 1.**^29–42^ For all traits, we used data from European-only populations and excluded duplicate and ambiguous AT/GC SNPs and SNPs with MAF≤1%. No GOCHA or EUR/ISGC cases or controls were included in the selected studies. For each trait, we generated a range of candidate GRS based on a combination of different r^2^ (0.1, 0.3, 0.5) and p-value thresholds (1×10^−8^ to 1) within each derivation dataset separately, using the association estimates (betas for each trait) as weights and a linkage disequilibrium (LD)-driven clumping procedure (European 1000 Genomes phase 3 was our reference LD panel).^43, 44^ Then, we merged the two training datasets and standardized (zero mean, unit standard deviation) the candidate GRS over the entire training set. Next, we selected the best-performing GRS for each trait (optimized GRS), based on the highest area under the receiving-operating characteristics curve (AUC) of a logistic regression model for ICH that included the candidate GRS, age, sex and the first two principal components of population structure as predictors. In case two or more models resulted in the same AUC, we chose the GRS with the largest effect size estimate for ICH risk, irrespective of directionality of effect (**Supplementary Tables 2-22**). This procedure resulted in a single optimized GRS representing each trait (**Supplementary Table 23).**

### Construction of the main ICH meta-Genomic Risk Score (metaGRS)

In order to generate an ICH metaGRS, we followed a standard meta-analytic approach, creating a weighted average of the trait-specific optimized GRS (**Supplementary Table 23)**.^45, 46^ Similar approaches have previously been used for ischemic stroke as well as for coronary artery disease (CAD).^45, 46^ We used GOCHA and EUR/ISGC to calculate logORs of the optimized GRS with ICH status (adjusted for age, sex, and two PCs) and pairwise Pearson correlations (ρ) between all GRS. Pooled logORs and correlations were generated after meta-analyzing under inverse-variance random-effects models across the two training datasets. We then used GERFHS (848 ICH cases and 794 ICH-free controls) as our validation dataset, where we calculated the ICH metaGRS as a weighted average of the standardized scores using the following formula:^45, 46^

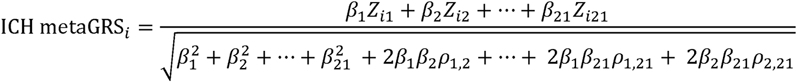

where *Z_i_*_1_,*Z_i_*_2_,…,*Z_i_*_21_ are the 21 different optimized GRS for the *i-th* individual in the GERFHS validation dataset, respectively; *β*_1_,*β*_2_,…,*β*_21_ are the meta-analyzed logORs of each score with ICH status in the training datasets; and *ρ_i,j_* are the meta-analyzed correlations between the *i-th* and *j-th* scores in the training datasets.

### Sensitivity analyses with alternative metaGRS

To uncover any underlying structure of groups of GRS affecting ICH risk, we performed exploratory maximum-likelihood factor analysis on the correlation matrix generated from the meta-analyzed Pearson correlations among the optimized GRS in the training datasets. To derive the appropriate number of factors, we used the Optimal Coordinate method as a non-graphical solution to the Cartell’s scree test.^47^ We applied varimax rotation to make the identified factors orthogonal, using a loading threshold of 0.4 for GRS inclusion.

In order to explore how inclusion and weighting of different GRS affect ICH risk, we constructed different versions of the metaGRS in the primary validation dataset, based on complementary GRS selection and weighting procedures in the training datasets. (‘causal metaGRS’, ‘factor metaGRS’, ‘stepwise metaGRS’, ‘lasso metaGRS’). First, we generated a metaGRS restricted to GRS of traits with prior significant associations with ICH risk in published studies, through either Mendelian Randomization (MR) or GRS analyses (‘causal metaGRS’).^48–52^ Next, we generated a metaGRS that contained only the GRS with the highest loadings within each identified factor from our exploratory factor analysis (‘factor metaGRS’). For these two metaGRS versions, the GRS weights corresponded to the univariate meta-analyzed logORs of the respective GRS in the training datasets. Then, using the R package ‘MASS’ ^53^, we employed stepwise logistic regression of ICH status on the 21 optimized GRS in the merged training datasets, using the Akaike Information Criterion (AIC) to identify the best-performing model containing the least number of GRS (‘stepwise metaGRS’). Additionally, we performed lasso regression using the R package ‘glmnet’ to model the associations between the 21 optimized GRS and ICH risk in the merged training datasets ^54^. Using 10-fold cross-validation in the merged training dataset, we estimated the value of lambda (coefficient shrinkage) that minimized the cross-validated prediction error rate (lambda = min), in order to find the most accurate model from the set of the 21 optimized GRS. Using this lambda value, we generated the next version of the metaGRS (‘lasso metaGRS’). For the last two metaGRS versions, the GRS weights corresponded to the adjusted and penalized logORs of the stepwise and lasso regressions, respectively, in the merged training datasets. All pairwise Pearson correlations among GRS for all metaGRS versions are estimates from random-effects meta-analysis across the training datasets.

### ICH metaGRS performance and clinical evaluation in GERFHS

We explored the metaGRS performance in the validation dataset and its comparison with clinical predictors. The primary metaGRS and its alternative versions were entered as linear predictors (1-standard deviation (SD) increment) in logistic regression models for ICH in the primary validation dataset. We performed several complementary analyses to investigate the association between the metaGRS and ICH. First, we assessed the odds of ICH in progressively higher metaGRS distributions, relative to the remainder of the sample. Next, we split the validation dataset into similar-sized metaGRS percentile groups (0%-15%, 15%-30%, 30%-45%, 55%-70%, 70%-85%, 85%-100%) and assessed the odds of ICH relative to the middle decile of the distribution (45%-55% group). To investigate potential deviations from linearity, we assessed how the odds of ICH changes in 5%-metaGRS-percentile increments, relative to the lowest 0%-10% of the metaGRS distribution. All analyses were adjusted for age, sex, and two PCs.^55^

In order to evaluate the potential clinical utility of the metaGRS, we first assessed whether the metaGRS is independently associated with ICH risk in a multivariable logistic regression model controlling for established clinical risk factors for ICH (BMI, history of ischemic stroke, hypertension, diabetes, high cholesterol, smoking history, alcohol use, anticoagulant medication use).^56, 57^ Four cases were excluded from this analysis – three had a previous ICH and one ICH case was a cavernoma. To restrict our clinical variables to those most strongly associated with ICH, we performed backward elimination on the logistic regression model containing the clinical risk factors alone. Factors with p<0.1 were included in the final restricted clinical model. To assess whether ICH risk modeling improves with the addition of genomic risk information, we calculated the AIC of the model containing only the clinical variables (‘All clinical’ model), the AIC of the model containing the clinical variables and the metaGRS (‘All clinical + metaGRS’ model), and compared the two nested models, utilizing the likelihood ratio test (LRT). These models also included age, sex, and 2 PCs as baseline predictors. In an alternative model, we explored whether the metaGRS is associated with ICH risk independently of *APOE* genotype, a known risk locus for lobar ICH.^58, 59^ *APOE* genotype was modeled as two variables, ε2 and ε4, coded for allele counts (0, 1, 2) in additive models referent to the wildtype ε3 allele.^59^ We explored potential interactions between the two *APOE* variables and the metaGRS. Finally, in order to evaluate and compare the predictive performances of the clinical variables with that of the metaGRS, we calculated the c-index and 95% confidence interval (CI) of models including the clinical risk factors separately, the ‘All-clinical’ model, and the ‘All-clinical + metaGRS’ model. The reference model contained only age, sex, and two PCs, and all models contained these variables as baseline predictors. 95% CIs were calculated after bootstrapping over 1,000 iterations in the derivation dataset. We generated percentile confidence intervals in bootstrapping. Similar analyses were performed for the different versions of the metaGRS. Complementary analyses were performed for patients with lobar and non-lobar ICH separately to assess whether the metaGRS was more predictive for specific subgroups of ICH patients.

### External validation of the metaGRS in UKBB

Lastly, we sought to externally validate the metaGRS in the UKBB, after calculating the metaGRS per variant score. To examine the derived score in a population-based setting, we calculated the metaGRS for participants without a history of ICH in the UKBB and explored associations with incident ICH events over follow-up. To calculate the metaGRS for each participant in the UKBB we transformed the per-GRS logOR weights to a per-variant score via a weighted sum using the following formula:^16^

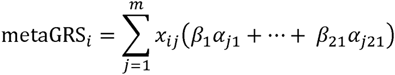

where *β*_1_,…,*β*_21_ are the standardized logORs (per-SD increment) of the 21 different optimized GRS for ICH in the training datasets (**Supplementary Table 23**), *a_j_*_1_,…,*a_j_*_21_ are the variant-specific effect sizes (from the GWAS summary statistics) for the *j-th* variant in each of the optimized GRS (*a_j_* was set to 0 for variants not included in the respective GRS), and *x_ij_* is the imputed or genotyped dose for the *i-th* individual’s *j-th* variant in the UKBB dataset. This resulted in a total of 2.6 million SNPs to be included in the final metaGRS.

The records of UKBB participants were linked with inpatient hospital episode statistics (HES), primary general practitioner data, and death registry for longitudinal follow-up. For the current study, we explored incident ICH, which was captured by the diagnostic algorithm for stroke in the UKB (detailed here: https://biobank.ndph.ox.ac.uk/showcase/ukb/docs/alg_outcome_stroke.pdf) up to December 2018. For events occurring after 2018 up to the end of follow-up (June 2020), we captured ICH events by the respective ICD-9 and ICD-10 codes in HES or death registry data (ICD-9 431.X and ICD-10 I61).

We next explored the effect of the metaGRS (per-SD increment) on incident ICH in a Cox proportional hazards model adjusted for age, sex, the first 10 PCs, genetic ancestry (White vs Other), kinship, and genotyping chip (UKB vs BiLEVE). We repeated this analysis in a high-risk group of individuals reporting use of antiplatelet and/or anticoagulant agents at baseline, as well as among a low-risk group with well-controlled vascular risk factors at baseline (blood pressure <140/90 mmHg, hemoglobin A1c <6.4%, no smoking, LDL-cholesterol <130 mg/dL, and no reported use of anticoagulant agents). Because several of the trait-specific GRS were constructed based partly on data derived from UKBB (**Supplementary Table 1**), we did not perform analyses including additional clinical predictors in order to avoid bias. To account for death as a potential competing risk, we applied the Fine-Gray subdistribution hazard approach.^60^

The UK Biobank has approval from the Northwest Multi-Center Research Ethics Committee. All participants provided written informed consent. We accessed the data following approval of an application by the UK Biobank Ethics and Governance Council (application # 36993).

Analyses were performed using R software version 3.6.1 (R Foundation for Statistical Computing)^61^ and SAS 9.4 (Cary, NC). Two-tailed p-values<0.05 were considered statistically significant.

## RESULTS

### Construction of a metaGRS for ICH in the training dataset

A schematic of our study design is provided in **Figure 1**. Following quality checks, we developed 21 optimized GRS for ICH-associated traits on the basis of associations with ICH in GOCHA and EUR/ISGC. The numbers of variants included in these GRS ranged from 213 to 1,148,192 (**Supplementary Table 23**). There was a high degree of correlation among the different trait-specific GRS in the training datasets (**Figure 2A**). In an attempt to clarify the correlation structure between the trait-specific GRS, we applied the Optimal Coordinate solution, which resulted in five orthogonal factors explaining 38.4% of the variance in the GRS correlation matrix (**Supplementary Figures 1-2**, **Supplementary Tables 24-25**). Applying a loading threshold of >0.4 resulted in factor 1 broadly representing cardiometabolic factors [waist-hip ratio (WHR), body mass index (BMI), smoking, HbA1c, high density lipoprotein (HDL), urine albumin-to-creatinine ratio (UACR), educational attainment (EA)]; factor 2 representing blood pressure (BP) traits (systolic and diastolic BP); factor 3 representing lipid traits [total cholesterol (TC) and low-density lipoprotein (LDL)]; factor 4 representing sleep traits (sleep duration and insomnia); and factor 5 representing pulse pressure.

**Figure 1.**
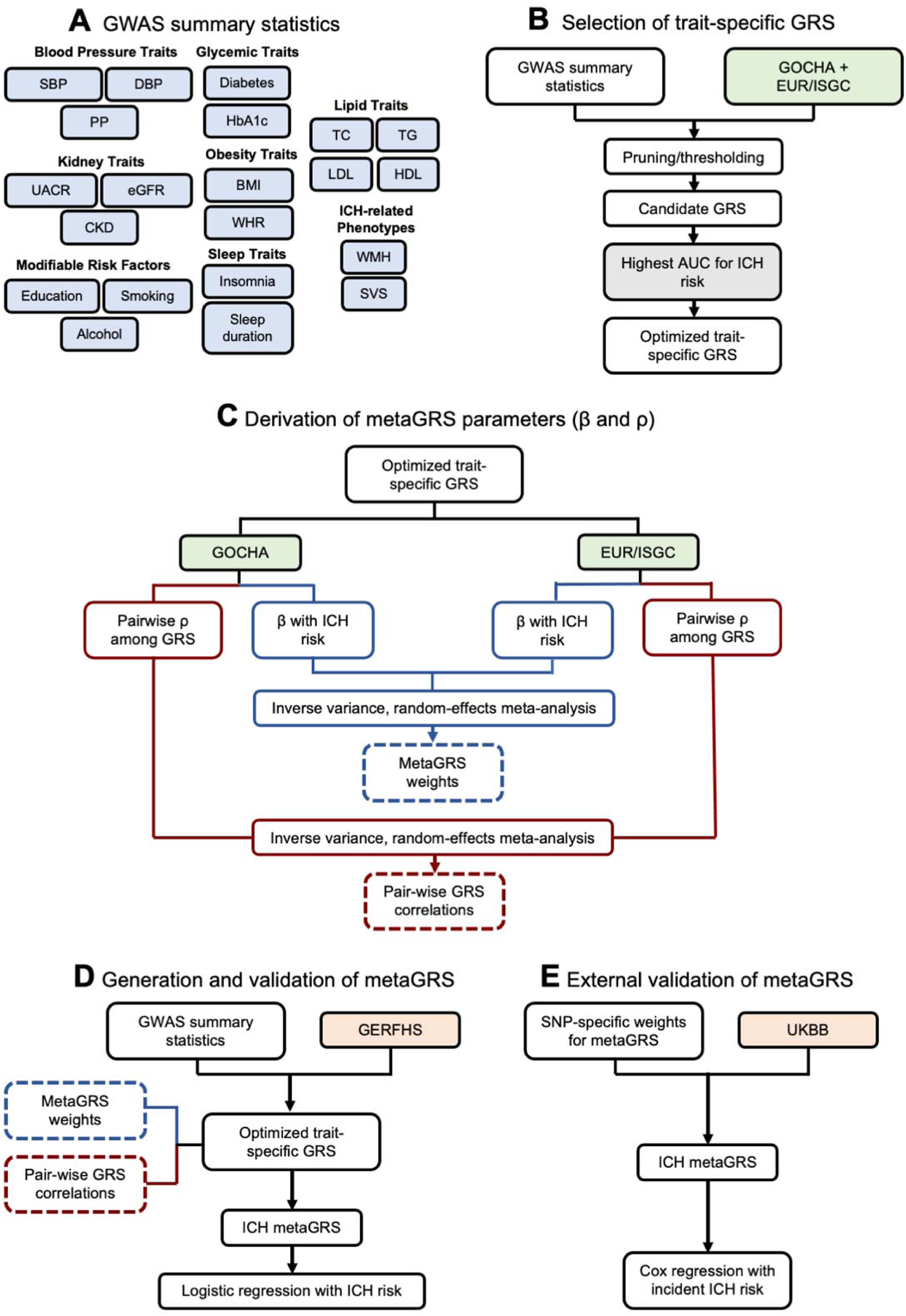
Study design. (**A**) Individual genomic risk scores (GRS) were derived for intracerebral hemorrhage (ICH)-related traits from publicly available summary statistics. (**B**) The GRS were optimized in the combined training dataset of GOCHA and EUR/ISGC (1013 cases, 928 controls). (**C**) MetaGRS parameters were determined on the basis of association with ICH in the training datasets. (**D**) The metaGRS was compiled in the GERFHS validation dataset (842 cases and 796 controls) and associations with ICH were explored using logistic regression. (**E**) The metaGRS was externally validated in the general population-based UK Biobank cohort among 486,623 individuals without a history of ICH over a median follow-up period of 11.3 years (1,526 incident events) using Cox proportional hazards regression analyses. SBP systolic blood pressure; DBP diastolic blood pressure; PP pulse pressure; WMH white matter hyperintensities; CKD chronic kidney disease; eGFR estimated glomerular filtration rate; UACR urine albumin-to-creatinine ratio; TC total cholesterol; TG triglycerides; LDL low-density lipoprotein; HDL high-density lipoprotein; T2D type 2 diabetes mellitus; HbA1c hemoglobin A1c; BMI body mass index; WHR waist-to-hip ratio; SVS small vessel stroke; GRS genomic risk score; GOCHA Genetics of Cerebral Hemorrhage on Anticoagulation; GERFHS Genetic and Environmental Risk Factors for Hemorrhagic Stroke; EUR/ISGC European member sites contributing to the International Stroke Genetics Consortium

**Figure 2.**
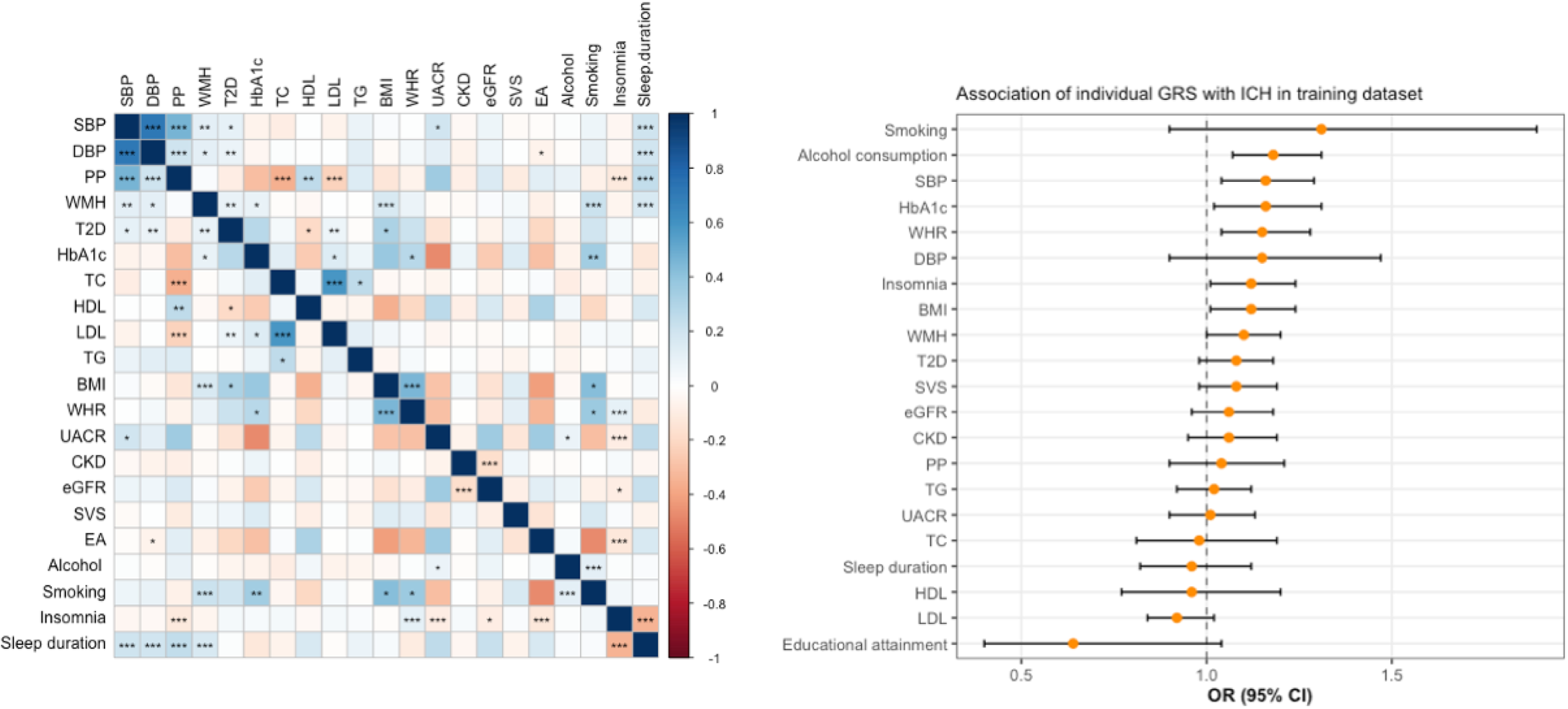
Individual genomic risk scores (GRS) for intracerebral hemorrhage (ICH)-related traits. **A.** Pairwise Pearson correlations among trait-specific GRS in training datasets (1,013 cases, 928 controls). FDR-p ***<0.001, **<0.01, *<0.05. **B.** Associations between trait-specific GRS (1 standard deviation increment) and ICH in training datasets (Odds Ratios, OR), as derived by logistic regression analyses adjusted for age, sex, and 2 principal components in training datasets (1,013 cases, 928 controls). Detailed estimates are presented in **Supplementary table 23**. SBP systolic blood pressure; DBP diastolic blood pressure; PP pulse pressure; WMH white matter hyperintensities; CKD chronic kidney disease; eGFR estimated glomerular filtration rate; UACR urine albumin-to-creatinine ratio; TC total cholesterol; TG triglycerides; LDL low-density lipoprotein; HDL high-density lipoprotein; T2D type 2 diabetes mellitus; HbA1c hemoglobin A1c; BMI body mass index; WHR waist-to-hip ratio; SVS small vessel stroke; EA educational attainment; GRS genomic risk score.

**Figure 2B** depicts the association estimates of the optimized GRS in the meta-analysis of the two training datasets, which we then used as weights to construct the metaGRS for ICH in the validation set (**Supplementary Table 23**). To avoid over-fitting and explore whether our approach for constructing the primary metaGRS influences associations with ICH, we also constructed alternative versions of the metaGRS following different GRS inclusion and weighting procedures: a ‘Causal metaGRS’ included 8 GRS, a ‘Factor metaGRS’ included 6 GRS, a ‘Stepwise metaGRS’ included 9 GRS with adjusted weights, and a ‘Lasso metaGRS’ included 20 GRS with penalized weights (**Supplementary Figure 3, Supplementary Table 25**).

### Associations between the metaGRS and ICH in the validation dataset

We explored associations between the derived metaGRS and ICH risk in GERFHS. The demographic and clinical characteristics of ICH cases and ICH-free controls in GERFHS are presented in **Table 1**. After adjusting for age, sex, and two PCs, a one SD increase in the metaGRS was associated with 45% higher odds of ICH (OR 1.45; 95% CI: 1.30-1.63; p=6.2×10^−11^). Patients in higher thresholds of the metaGRS distribution were at progressively higher risk for ICH (**Figure 3**). Notably, patients in the top 2.5% and 1% had substantially increased odds of ICH, respectively, compared to the rest of sample (OR 3.72; 95% CI: 1.84-8.33; p=5.77×10^−4^, and OR 4.83; 95% CI: 1.56-21.2; p=0.01, respectively). We observed an expected gradual change in ICH odds when moving to either the higher or lower ends of the metaGRS distribution from the middle decile (**Supplementary Figure 4**). Modeling risk of ICH relative to the bottom 10% of the metaGRS loading did not reveal any significant non-linear effects (**Supplementary Figure 5**).

**Figure 3.**
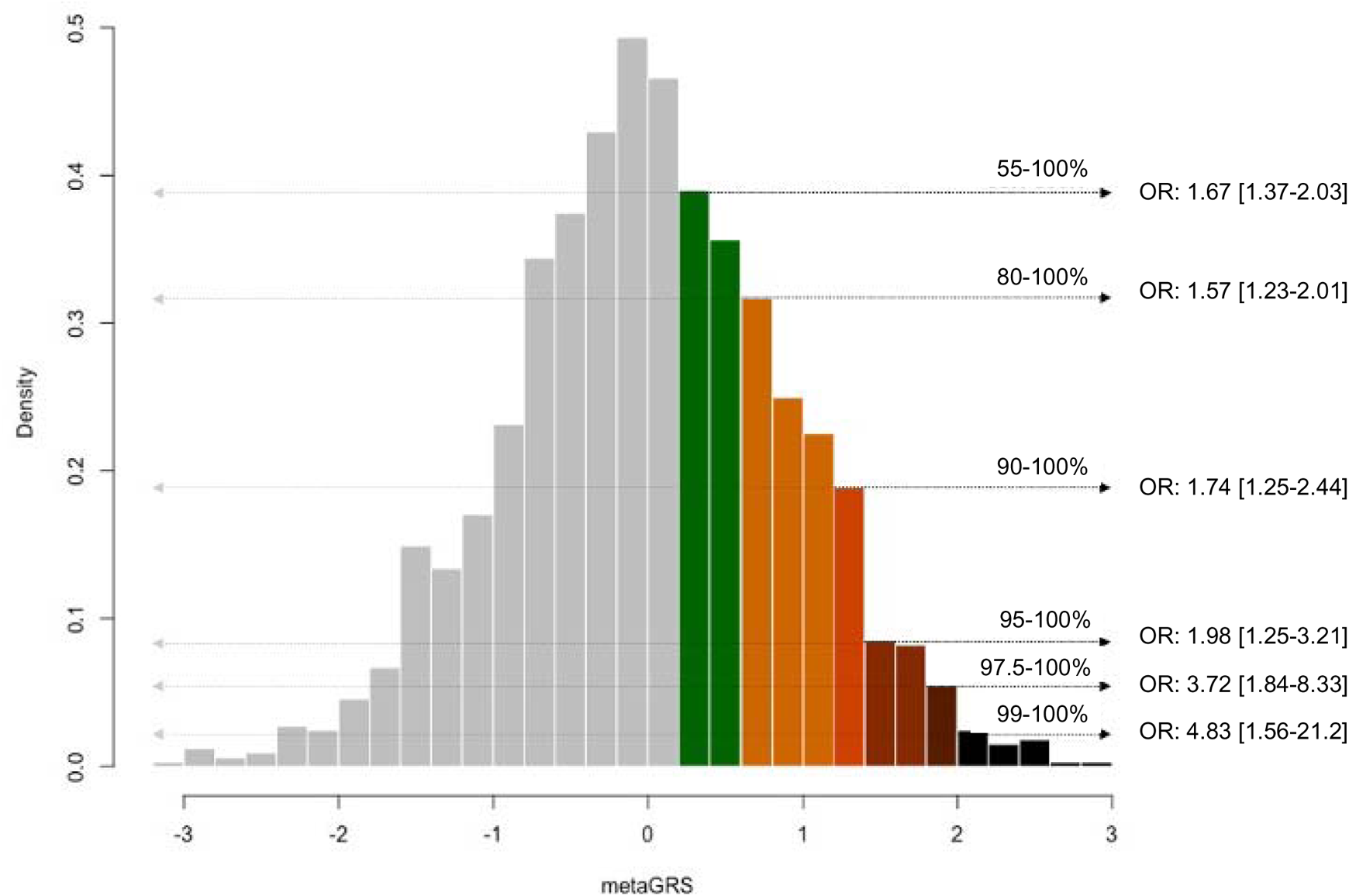
Odds for intracerebral hemorrhage (ICH) across the metaGRS distribution. Depicted is the metaGRS distribution (centered around a mean of 0 and a standard deviation of 1) in the GERFHS validation dataset (848 cases and 794 controls) and the odds ratios (OR) for ICH per percentile group, relative the rest of the sample, as derived from logistic regression models adjusted for age, sex and the first 2 principal components of the population structure.

**Table 1.** Clinical characteristics of the GERFHS validation dataset for intracerebral hemorrhage (ICH) cases and ICH-free controls.

| Variable | Controls N=796 | Cases N=842 | p-value |
| --- | --- | --- | --- |
| Age: mean (SD) | 68.4 (13.4) | 69.2 (14.1) | 0.2248 |
| Sex: female | 400 (50.2) | 417 (49.5) | 0.7689 |
| Race: white | 796 (100.0) | 841 (99.9) | - |
| Education: >high school | 439 (55.2) | 294 (35.2) | <0.0001 |
| Hypertension |  |  | <0.0001 |
| Treated hypertension | 393 (49.4) | 403 (48.4) |  |
| Untreated hypertension | 29 (3.6) | 159 (19.1) |  |
| Diabetes mellitus | 131 (16.5) | 190 (22.6) | 0.0019 |
| Hypercholesterolemia | 407 (51.5) | 366 (44.9) | 0.008 |
| History of ischemic stroke | 21 (2.6) | 76 (9.0) | <0.0001 |
| BMI: median (IQR) | 27.1 (23.8, 31.0) | 26.7 (22.9, 31.2) | 0.1459 |
| Smoking |  |  | 0.1477 |
| Never | 373 (46.9) | 365 (43.6) |  |
| Former | 325 (40.9) | 345 (41.2) |  |
| Current | 97 (12.2) | 128 (15.3) |  |
| Heavy alcohol use | 38 (4.8) | 54 (6.8) | 0.0812 |
| Anticoagulant medications | 51 (6.4) | 143 (17.0) | <0.0001 |
| ICH location: lobar |  | 334 (39.7) | - |

The values correspond to N (%), except as otherwise stated. Two-sided p-values are presented as derived by chi-square test for independence in contingency tables, Mann-Whitney U test, as appropriate.
GERFHS: Genetic and Environmental Risk Factors for Hemorrhagic Stroke, SD: standard deviation, IQR: interquartile range.

In sensitivity analyses comparing ICH odds between the primary and the alternative metaGRS, we overall found that the primary metaGRS achieved the best performance, indicating that information contained within GRS that were excluded from the restricted metaGRS versions contributed independently to ICH odds (**Supplementary Figure 6**). However, these metaGRS with penalized and adjusted weights still demonstrated significant associations with ICH risk, albeit with weaker effect size estimates and lower predictive performances (**Supplementary Figure 6** and **Supplementary Table 26**).

### Predictive performance of metaGRS for ICH in comparison with clinical risk factors

We next explored the performance of the metaGRS with established clinical risk factors for predicting ICH in GERFHS. We performed backward elimination to identify the optimal set of clinical risk factors to be included in the model and found history of ischemic stroke, hypertension, diabetes, high cholesterol, heavy alcohol use, anticoagulant use, and education less than high school to be most strongly associated with ICH risk (**Supplementary Table 27**). After adjusting for these clinical risk factors in a multivariable logistic regression model, the metaGRS continued to be independently associated with ICH risk (OR = 1.31 per one standard deviation of the metaGRS; 95% CI 1.16 – 1.48 p < 0.0001). Adding the metaGRS to a model including the clinical risk factors significantly improved the model fit (AIC of clinical predictors = 1989.65, AIC of clinical predictors + metaGRS = 1972.86, LRT = 4×10^−5^).

Comparing the c-indices of models including individual clinical risk factors, the metaGRS showed comparable predictive performance to hypertension, and higher than the remaining clinical risk factors apart from education (**Figure 4**). Importantly, the c-index of a model including the entire set of clinical risk factors (C: 0.686, 95%CI: 0.663-0.718) increased after including the metaGRS in the model (C: 0.695, 95%CI: 0.673 – 0.727, **Figure 4**). Similar results were observed for the alternative metaGRS versions, which were each independently associated with ICH and contributed significantly to clinical risk models (**Supplementary Figure 7 and Supplementary Table 28**).

**Figure 4.**
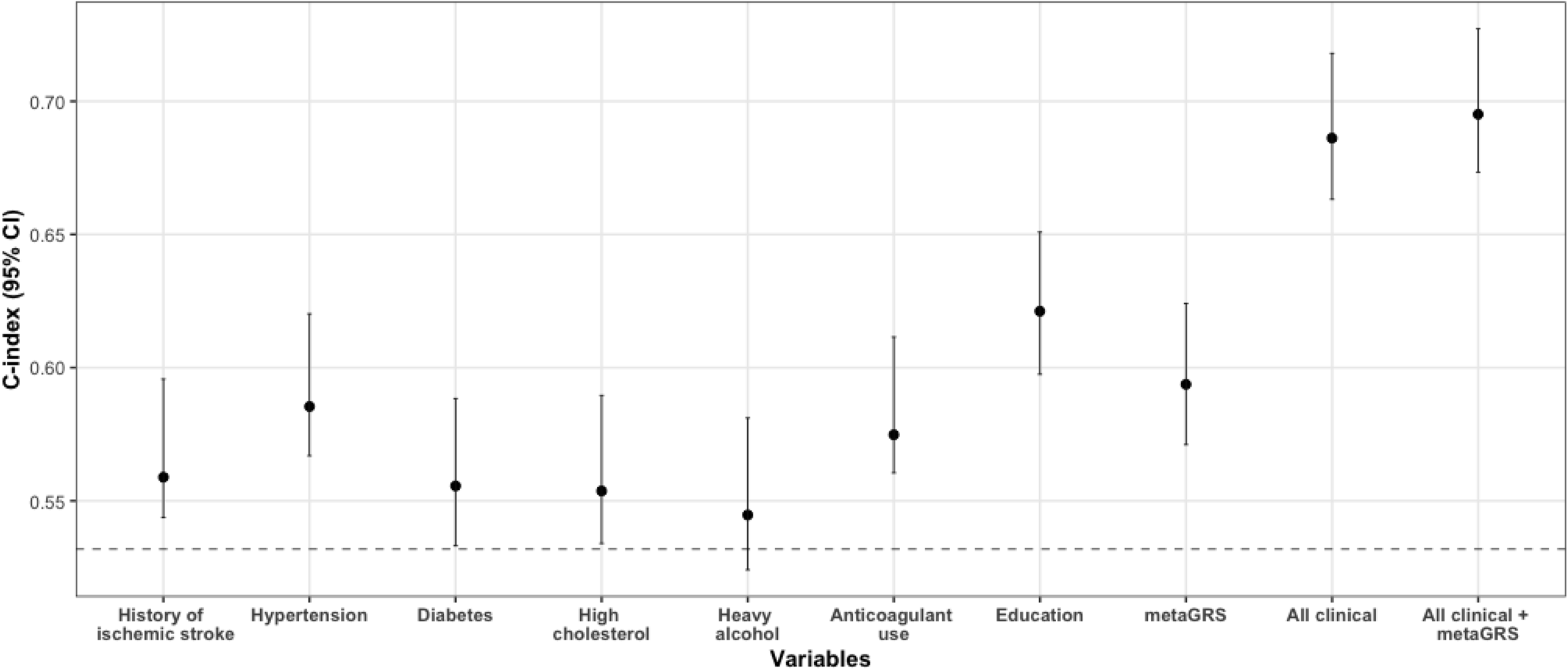
Performance of clinical risk factors, metaGRS, and their combination for predicting odds for intracerebral hemorrhage (ICH). Depicted are the c-indices derived from logistic regression models including age, sex, 2 principal components of population structure (baseline model, dotted line) and additionally in successive models the reported clinical variables (history of ischemic stroke, hypertension, diabetes, high cholesterol, heavy alcohol use, anticoagulant medication use, and education) and the metaGRS in the validation GERFHS dataset (842 cases, 796 controls). Percentile confidence intervals of the c-indices were calculated after bootstrapping over 1000 iterations.

Further, we explored whether the metaGRS is differentially associated with location-specific subtypes of ICH related to different underlying pathologies. Lobar and non-lobar characteristics as well as significant clinical predictors for location-specific ICH subtypes are presented in **Supplementary Tables 29-32**. We found higher odds of both non-lobar (OR 1.37; 95%CI: 1.18-1.58; p=2.4×10^−5^) and lobar ICH (OR 1.32; 95%CI: 1.13-1.54; p=5×10^−4^) per one SD increment in metaGRS following adjustment for clinical risk factors (**Table 2**). Inclusion of the metaGRS to models already adjusted for the entire set of clinical risk factors led to significant decreases in AIC of the models for each subtype (**Table 2**).

**Table 2.** Associations between the metaGRS (1-SD increment) with odds of lobar and non-lobar intracerebral hemorrhage (ICH) in the GERFHS validation dataset.

|  | **Lobar ICH (324 cases vs 794 controls)** |  |  |
| --- | --- | --- | --- |
|  | **OR (95% CI)** | **P-value** | **AIC<sub>clin</sub> , AIC<sub>clin+metaGRS</sub>, LRT** |
| All trait metaGRS | 1.32 (1.13-1.54) | 0.0005 | 1271.20, 1260.85, 0.001291245 |
|  | **Non-lobar ICH (446 cases vs 788 controls)** |  |  |
|  | **OR (95% CI)** | **P-value** | **AIC<sub>clin</sub> , AIC<sub>clin+metaGRS</sub>, LRT** |
| All-trait metaGRS | 1.37 (1.18-1.58) | $2.4 \times 10^{-5}$ | 1465.15, 1449.95, 0.000096755 |

Lobar ICH model is adjusted for age, sex, 2 principal components (PC), body mass index, history of ischemic stroke, anticoagulant medication use, and education less than high school. Non-lobar ICH model is adjusted for history of ischemic stroke, hypertension, diabetes, high cholesterol, heavy alcohol use, anticoagulant use, and education less than high school.
AIC = Akaike information criterion. AIC<sub>clin</sub> = AIC of model containing baseline (age, sex, 2 PCs) and ICH-subtype specific clinical predictors. AIC<sub>clin+metaGRS</sub> = AIC of model containing baseline, sub-type specific clinical predictors and All-trait metaGRS. GERFHS: Genetic and Environmental Risk Factors for Hemorrhagic Stroke. LRT = likelihood ratio test.

When exploring in the same model the metaGRS and *APOE* genotype, a known risk locus for ICH, we found that both were independently associated with the odds of ICH (**Supplementary Table 33**). The metaGRS was associated with the odds of both lobar and non-lobar ICH when adjusting for APOE genotype, whereas the ε2 and the ε4 alleles of the APOE locus were associated only with the odds of lobar ICH. There was no evidence of an interaction between the two genomic markers (**Supplementary Table 33**).

### Validation of the metaGRS in the UK Biobank population

As a final step, we explored associations between the metaGRS and incident ICH risk in a general population sample. In the prospective population-based UKBB cohort, a total of 486,623 participants without a history of ICH (mean age 56.5 ± 8.1 years, 54.2% females), were followed-up for a median of 11.3 years (IQR: 10.6-11.1 years). The baseline characteristics of the study participants are presented in **Table 3**. We again found the metaGRS to be associated with a higher risk of incident ICH (HR per SD increment: 1.15, 95%CI: 1.09-1.21, p=7×10^−7^) in a Cox proportional hazards model adjusted for age, sex, the first 10 PCs of population structure, kinship, genotyping chip, and genetic ancestry. Accounting for death as a competing risk with the subdistribution hazard approach also did not substantially alter the results (HR per SD increment of metaGRS: 1.14, 95%CI: 1.08-1.20, p=4×10^−6^). **Figure 5A** presents the Kaplan-Meier curves with age as the time variable for individuals at upper, median, and lower quantiles of the metaGRS. We explored whether the metaGRS retains association with incident ICH risk among groups of patients with different levels of baseline risk. Within both high-risk individuals using antithrombotic medications at baseline, as well as low-risk individuals with well-controlled vascular risk factors at baseline and no antithrombotic medications, the metaGRS retained association with incident ICH risk (**Figures 5B and 5C**).

**Figure 5.**
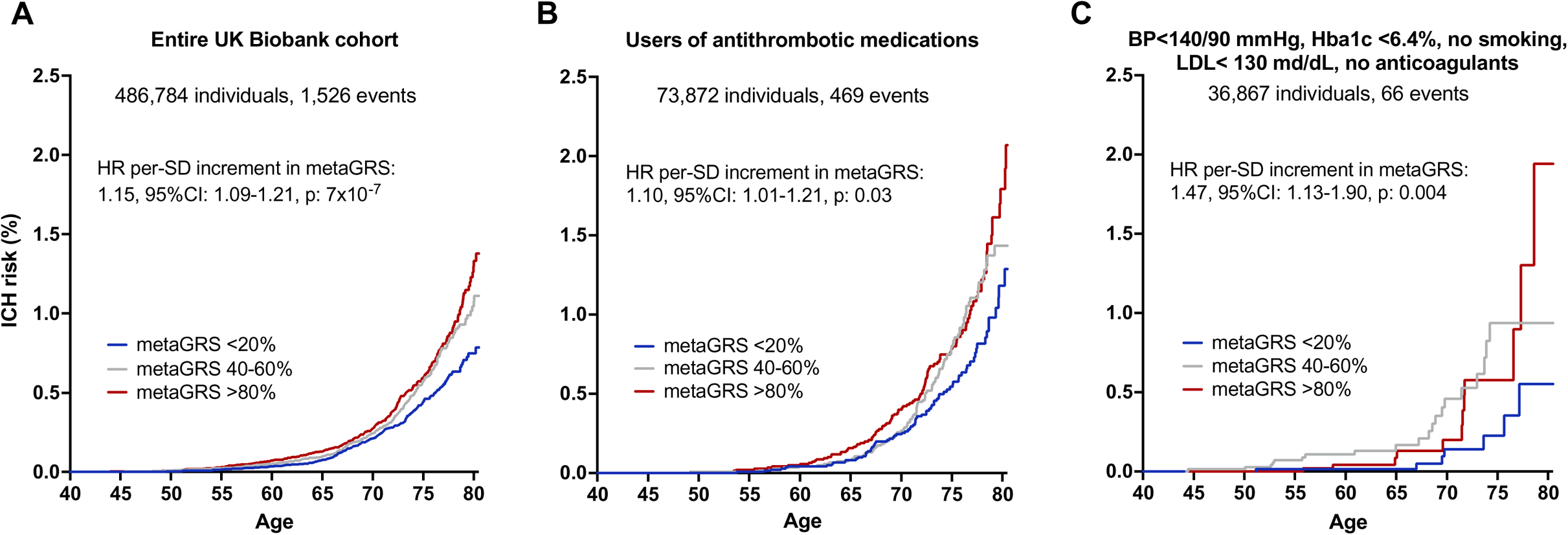
MetaGRS and cumulative risk of incident intracerebral hemorrhage (ICH) in the population-based UK Biobank sample. The results are derived from (A) entire UK Biobank population, (B) users of antithrombotic medications at baseline, and (C) low-risk individuals with conventional vascular risk factors under control and no use of anticoagulant medications after excluding prevalent cases of ICH. Depicted are the Kaplan-Meier curves for different metaGRS quantiles, as well as the hazard ratios (HR) per standard deviation (SD) increment in metaGRS, as derived from Cox proportional hazards regression models adjusted for baseline age, sex, the first 10 principal components of the population structure, genetic ancestry, genotyping chip, and kinshi

**Table 3.**
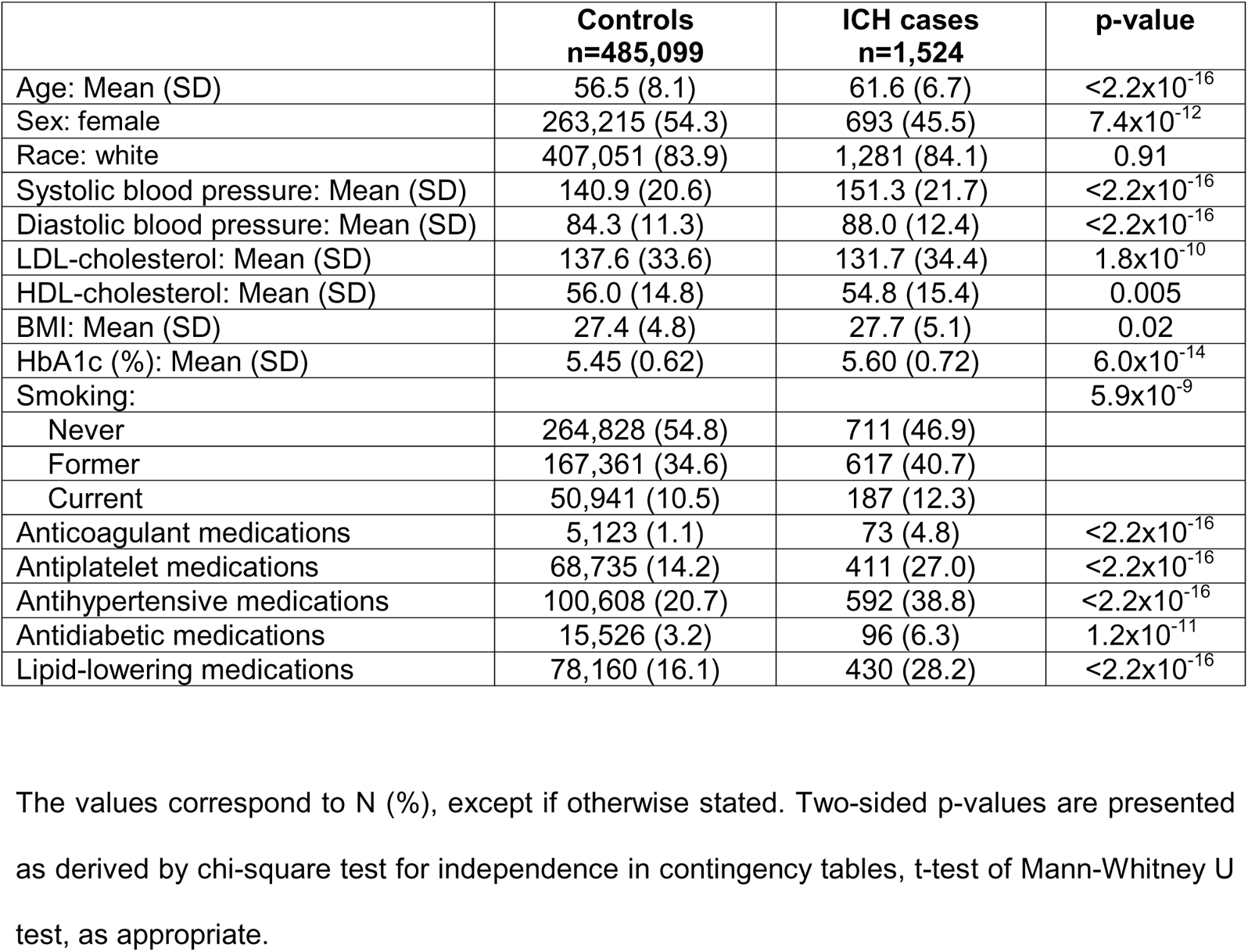
Baseline characteristics of the external UK Biobank dataset for individuals who developed incident intracerebral hemorrhage (ICH) over a median 11.3-year period of follow-up and ICH-free controls.

|  | **Controls**<br>**n=485,099** | **ICH cases**<br>**n=1,524** | **p-value** |
| --- | --- | --- | --- |
| Age: Mean (SD) | 56.5 (8.1) | 61.6 (6.7) | $<2.2 \times 10^{-16}$ |
| Sex: female | 263,215 (54.3) | 693 (45.5) | $7.4 \times 10^{-12}$ |
| Race: white | 407,051 (83.9) | 1,281 (84.1) | 0.91 |
| Systolic blood pressure: Mean (SD) | 140.9 (20.6) | 151.3 (21.7) | $<2.2 \times 10^{-16}$ |
| Diastolic blood pressure: Mean (SD) | 84.3 (11.3) | 88.0 (12.4) | $<2.2 \times 10^{-16}$ |
| LDL-cholesterol: Mean (SD) | 137.6 (33.6) | 131.7 (34.4) | $1.8 \times 10^{-10}$ |
| HDL-cholesterol: Mean (SD) | 56.0 (14.8) | 54.8 (15.4) | 0.005 |
| BMI: Mean (SD) | 27.4 (4.8) | 27.7 (5.1) | 0.02 |
| HbA1c (%): Mean (SD) | 5.45 (0.62) | 5.60 (0.72) | $6.0 \times 10^{-14}$ |
| Smoking: | | | $5.9 \times 10^{-9}$ |
| Never | 264,828 (54.8) | 711 (46.9) |  |
| Former | 167,361 (34.6) | 617 (40.7) |  |
| Current | 50,941 (10.5) | 187 (12.3) |  |
| Anticoagulant medications | 5,123 (1.1) | 73 (4.8) | $<2.2 \times 10^{-16}$ |
| Antiplatelet medications | 68,735 (14.2) | 411 (27.0) | $<2.2 \times 10^{-16}$ |
| Antihypertensive medications | 100,608 (20.7) | 592 (38.8) | $<2.2 \times 10^{-16}$ |
| Antidiabetic medications | 15,526 (3.2) | 96 (6.3) | $1.2 \times 10^{-11}$ |
| Lipid-lowering medications | 78,160 (16.1) | 430 (28.2) | $<2.2 \times 10^{-16}$ |

The values correspond to N (%), except if otherwise stated. Two-sided p-values are presented as derived by chi-square test for independence in contingency tables, t-test of Mann-Whitney U test, as appropriate.

## DISCUSSION

We developed a genomic risk score for ICH in a training dataset of 1,013 cases and 928 controls based on GWAS data for 21 ICH-related traits. We found the derived metaGRS to be significantly associated with the odds of ICH in an independent validation dataset of 842 ICH cases and 796 ICH-free controls. The metaGRS was independent of traditional clinical risk factors of ICH and improved model performance in prediction of ICH. Furthermore, the score was significantly associated with incident ICH risk in a population-based cohort study of 480,000 individuals followed-up for a median of 11 years (1,500 incident ICH events). Our results provide important insights into genomic prediction for ICH and could have implications for clinical practice.

First, the metaGRS identified individuals at very high risk for ICH. For example, individuals at the top percentile had almost 5-fold increased odds for ICH, as compared to the rest of the population. While it remains to be clarified how these individuals would benefit from potential primary preventive interventions, this information could be useful both for screening for hypertension, the main clinical risk factor for ICH, and early initiation of antihypertensive treatment, as well as for decision making when considering initiation of antiplatelet or anticoagulation treatments that might increase ICH risk. These risk stratification strategies based on genomic information are increasingly important as millions of persons in the US and around the world have been genotyped by direct-to-consumer genotyping companies.

Second, the metaGRS improved risk discrimination for ICH when compared to classical clinical predictors. Specifically, it was associated with ICH risk independently of vascular risk factors and was found to have a predictive value superior to all predictors except for education. The predictive power of the metaGRS was comparable to that of hypertension, the most well-established clinical risk factor for ICH that explains the most variance in the trait.^62^ These findings support the incorporation of genetic information into clinical tools aiming to quantify ICH risk within specific patient subgroups. A post hoc analysis of trial data showed that among patients with atrial fibrillation and a CHA2-DS2-VASc score of 2, a high genomic risk score for ischemic stroke led to an absolute ischemic stroke risk equivalent to those with a higher score.^63^ Whether integration of a genomic risk score for ICH in such analyses could lead to a more precise assessment of the risk-benefit ratio for specific patients remains to be determined. Along these lines, the several clinical trials currently evaluating anticoagulation as a secondary prevention strategy after ICH constitute a unique opportunity for genomic-based risk-stratification, as a portion of them have built-in biobanks that are collecting DNA samples.

Third, despite its rarity (0.3% in the UKBB sample), we found the metaGRS to be significantly associated with prospective ICH risk in the general population. The metaGRS was associated with a higher risk of ICH even among individuals with evidence-based control of relevant risk factors, who were not actively smoking, had blood pressure of 140/90 mmHg or less, no evidence of diabetes, normal BMI, and who reported no use of anticoagulants. While such analyses are restricted by lack of power, our results suggest that for specific individuals with a high genetic risk, the recommended treatment targets for modifiable risk factors might not be sufficient for primary ICH prevention. The importance of this observation lies on the fact that the genomic information is available long before vascular risk factors are present and could thus be used for earlier risk stratification in otherwise low-risk individuals. Concomitantly, the metaGRS was also associated with a higher risk of ICH even among a high-risk group of individuals using antithrombotic medications, indicating its potential utility among a relevant group of patients for whom bedside calculation of ICH risk might be particularly relevant to clinical care.

Our study has limitations. First, the sample sizes of the available genetic datasets for ICH are limited, as compared to other clinical endpoints. This introduces uncertainty to the association estimates between the genetic variants and ICH risk, which were used to construct the described metaGRS, and thus impacts negatively on its predictive performance. Indeed, when compared with metaGRS for other traits that have been developed in larger datasets, such as for coronary artery disease and ischemic stroke,^16, 64^ the association with incident ICH events is weaker. Second, ICH is a phenotypically heterogeneous disease, with the most common etiologies being hypertensive small vessel disease (typically in non-lobar locations) and cerebral amyloid angiopathy (typically in lobar locations). To maximize the power of our approach, we have pooled cases, which could have negatively impacted the predictive performance for specific ICH etiologies. While our score was predictive for both non-lobar and lobar ICHs, developing etiology-specific scores might be of more relevance for specific clinical scenarios. Third, while the metaGRS showed significant associations with risk of incident ICH in the UKBB, we could not explore its effects in concert with other clinical predictors, because the metaGRS was generated using associations with these predictors in datasets including data from the UKBB. Therefore, independent validation either of a score trained in an entirely UK Biobank-independent dataset or of the described metaGRS in another external cohort would be necessary. Fourth, the metaGRS was constructed solely on the basis of data from individuals of European genetic ancestry, and may thus not be applicable for individuals of other ancestries. Larger multi-ethnic GWAS studies of ICH currently underway will facilitate the generation of ancestry-specific GWAS datasets. Last, overarching limitations and challenges still exist on the generation and validation of GRS across disease states, which apply to our metaGRS as well. Some of them include the possible differences in sex-specific predictive performances, the translation of GRS estimates from the cohort-to the individual-specific level which has been suggested to introduce additional variability, as well as the heterogeneity of the different methods for GRS construction which could ultimately hinder clinical application.^11, 12^ Towards that end, efforts are currently underway to standardize and delineate procedures surrounding GRS construction and reporting, such that prediction models incorporating GRS-based estimations can be leveraged in a consistent and reproducible manner.^65^

In conclusion, our study represents the first comprehensive attempt to develop and validate a genomic risk score for ICH. Our results demonstrate that the incorporation of genomic information in clinical prediction models for ICH could enhance predictive performance. As such, it lays the groundwork for future analyses in larger genetic datasets for ICH to optimally combine genomic information to maximize predictive benefit. Exploration of the performance of genomic risk scores for ICH in clinical trials of patients receiving antithrombotic medications could offer useful insights in risk prediction of ICH in this high-risk population with potential relevance for clinical decision making.

## Supporting information

MetaGRS_ICH_Supplemental Material

## Abbreviations

ICH: intracerebral hemorrhage
GWAS: genome-wide association studyGRS genomic risk score
SNP: single nucleotide polymorphism
GOCHA: Genetics of Cerebral Hemorrhage on Anticoagulation
GERFHS: Genetic and Environmental Risk Factors for Hemorrhagic Stroke
EUR/ISGC: European member sites contributing to the International Stroke Genetics Consortium
UKBB: UK Biobank
QC: quality control
PC: principal component
HRC: Haplotype Reference Consortium
MAF: minor allele frequency
LD: linkage disequilibrium
AUC: area under the receiving-operating characteristics curve
CAD: coronary artery disease
AIC: Akaike Information Criterion
HES: hospital episode statistics
SBP: systolic blood pressure
DBP: diastolic blood pressure
PP: pulse pressure
WMH: white matter hyperintensities
CKD: chronic kidney disease
eGFR: estimated glomerular filtration rate
UACR: urine albumin-to-creatinine ratio
TC: total cholesterol
TG: triglycerides
LDL: low-density lipoprotein
HDL: high-density lipoprotein
T2D: type 2 diabetes mellitus
HbA1c: hemoglobin A1c
BMI: body mass index
WHR: waist-to-hip ratio
SVS: small vessel stroke
EA: educational attainment

## Acknowledgements

**Funding:** MKG is supported by a Walter-Benjamin fellowship from the German Research Foundation (Deutsche Forschungsgemeinschaft [DFG], GZ: GE 3461/1-1) and the FöFoLe program of LMU Munich (FöFoLe-Forschungsprojekt Reg.-Nr. 1120). HIH is suported by NIH R01HL138423, R01HL156024, R01HL138423-05S1 and 1R01AG072592. GJF is supported by the NIH (K76AG059992, R03NS112859), the American Heart Association (18IDDG34280056 and 817874), and a pilot grant from the Claude D. Pepper Older Americans Independence Center at Yale (P30AG021342). JR and CDA are supported by NIH R01NS103924, U01NS069673, AHA 18SFRN34250007, and AHA-Bugher 21SFRN812095. GOCHA acknowledges funding from the NIH-National Institute of Neurologic Disorders and Stroke grants K23NS042695, R01NS059727 and 5R01NS042147, the Deane Institute for Integrative Research in Atrial Fibrillation and Stroke, the Keane Stroke Genetics Research Fund, the Edward and Maybeth Sonn Research Fund, the University of Michigan General Clinical Research Center (M01 RR000042) and a grant from the National Center for Research Resources. GERFHS acknowledges funding from the NIH grants NS36695, NS30678, and the Greater Cincinnati Foundation Grant (Cincinnati Control Cohort). The European ISGC studies acknowledge funding from the Ministerio de Sanidad y Consumo de España, Instituto de Salud Carlos III with the grants: “Registro BASICMAR” Funding for Research in Health (PI051737), “GWALA project” from Fondos de Investigación Sanitaria ISC III (PI10/02064), Fondos FEDER/EDRF Red de Investigación Cardiovascular (RD12/0042); Contract for Research Training for Professionals with Specialty (CM06100067); “Ramon y Cajal” Postdoctoral Contract and Grant from Spanish Research Networks “Red HERACLES” (RD06/ 0009) (HM-ICH); the Spanish government grants (Geno-tPA project-FIS PJ060586 and GRECAS project EC08/00137) (VHH-ICH); the Polish Ministry of Education grant N402 083934 (JUHSS); the Lund University, Region Skåne and the Swedish Medical Research Council (K2007-61X - 20378-01-3, K2010-61X-20378-04-3), the Swedish Stroke Association, the Freemasons Lodge of Instruction EOS in Lund, and the King Gustaf V and Queen Victoria’s foundations. Biobank services and genotyping were performed at Region Skåne Competence Centre (RSKC Malmö), Skåne university hospital, Malmö, Sweden (LUHSS); the Austrian Science Fond (FWF, P20545-P05 and P13180) and the Medical University of Graz supports the databank of the ASPS (MUC-ICH).

## Supplemental Material

Table of Contents Supplemental Methods

Supplemental Figures and Figure Legends

Supplemental Tables and Supporting Information

Supplemental References

