## Supplementary material for "A Genomic Risk Score Identifies Individuals at High Risk for Intracerebral Hemorrhage": MetaGRS_ICH_Supplemental Material

### Supplemental Methods: participating studies

#### Genetics of Cerebral Hemorrhage with Anticoagulation (GOCHA) Study

GOCHA is a multi-center case-control study of primary intracerebral hemorrhage (ICH) conducted in the Unites States[1]. Massachusetts General Hospital (MGH) was the coordinating center along with five additional participating centers: Mayo Clinic (Jacksonville, FL), University of Virginia Health System, University of Florida College of Medicine, University of Michigan Health System, and Beth Israel Deaconess Medical Center. Subjects were recruited between 1999 and 2010, on the basis of one of the following categories: warfarin-related ICH patients, matched warfarin-related controls with no history of ICH, no-treatment ICH (patient with ICH not taking warfarin at time of hemorrhage), and no-treatment controls. Inclusion criteria for cases were the following: symptomatic ICH, sufficiently severe to cause patient to seek medical care; age > 55, either gender without upper age limit to participation; and ability and willingness of the patient or appropriate surrogate to provide informed consent. Inclusion criteria for controls were the same with the exception that subjects did not have symptomatic ICH. Case exclusion criteria included the following: if taking warfarin, initial INR at presentation < 1.4; concussive head trauma in the 24 hours before presentation (unless it was a clear result of the ICH); clinical, radiographic, or pathological evidence of any of the following: ischemic stroke, occurring during the 2 weeks prior to presentation, primary or metastatic intracerebral tumor, intracerebral vascular malformation, or vasculitis of the central nervous system; antecedent use of cocaine or sympathomimetic drug; antecedent alcohol abuse; medical history of primary coagulopathy, blood dyscrasia, or active liver disease; greater than 1 week interval between symptom onset and presentation at enrolling center. Exclusion criteria for controls included the following: history of symptomatic ICH; if taking warfarin, initial INR at presentation < 1.4; antecedent use of cocaine or sympathomimetic drug; antecedent alcohol abuse; medical history of primary coagulopathy, blood dyscrasia, or active liver disease. Cases were identified through presentation or transfer to participating institutions, based on the aforementioned criteria. Control subjects were selected through either the MGH

Anticoagulation Unit or review of medical records via the Research Patient Database Registry and were matched for race, ethnicity, and gender.

**Genetic and Environmental Risk Factors for Hemorrhagic Stroke (GERFHS) Study**

GERFHS is a case-control study of primary ICH in the Greater Cincinnati/Northern Kentucky region[2]. Subjects were recruited from all 16 adult hospitals within a 50-mile radius from the University of Cincinnati. Inclusion criteria were spontaneous ICH; age ≥18 years; resident for 6 months within 50 miles of the University of Cincinnati; no evidence of trauma, brain tumor, or vascular malformation as a cause of hemorrhage; and ability to obtain informed consent from the patient or an appropriate surrogate. Cases would not be enrolled if they were contacted > 90 days from the day of stroke. Controls were matched by age (+/- 5 years), race, and gender from the same population, sampled by random digit dialing.

**European member sites contributing to the International Stroke Genetics Consortium (EUR/ISGC)**

These included the Hospital del Mar Intracerebral Hemorrhage (HM-ICH) and the Vall d'Hebron Hospital ICH (VVH-ICH) studies in Barcelona, Spain; the Jagiellonian University Hemorrhagic Stroke (JUHS) Study in Krakow, Poland; the Lund Stroke Register (LSR) study in Lund, Sweden, and the Medical University of Graz ICH study (MUG-ICH) in Graz, Austria. Due to their limited sample sizes, data from these sites were analyzed together for the purposes of quality control, imputation, and association testing in the primary genome-wide association meta-analysis[3]. The HM-ICH study was a retrospective study of primary, nontraumatic ICH cases enrolled in a stroke registry. Exclusion criteria included mRS > 1, ICH due to anticoagulant treatment, multiple ICH, primary intraventricular hemorrhage (IVH), and ICH secondary to tumor and vascular malformations[4]. The VVH-ICH study recruited patients with age ≥ 55 years, diagnosed with lobar ICH. Patients were excluded if they had lobar ICH related to vascular malformation, impaired coagulation or oral anticoagulation use, traumatic brain injury, tumoral bleed, if they underwent a surgical procedure, if they died within the first 6 months after the ICH and if they had multiple lobar and deep microbleeds on MRI[5]. The JHUS study recruited consecutively admitted primary ICH patients of Caucasian origin without evidence of isolated intraventricular hemorrhage, head trauma, vascular malformations, malignancy, hemorrhagic transformation of ischemic stroke, or blood dyscrasias and age- and sex-matched controls[6]. The LSR study was a prospective population-based study of stroke in the Lund-Orup district. Cases were ascertained mainly (90.4%) through prospective methods, including regular queries of patient lists in the Emergency Unit at Lund University Hospital, community health care centers and general practitioners. Cases included only first-ever strokes.[7] The MUG-ICH study included only consecutive primary non-traumatic ICH cases confirmed on a computed tomography scan and without evidence of hemorrhagic transformation of ischemic stroke, tumor, or vascular malformation[8].

### Supplemental Figures and Figure Legends

**Supplementary Figure 1. Non-graphical solutions to Scree Test. The optimal coordinates method resulted in n=5 factors representing the GRS correlation matrix.**

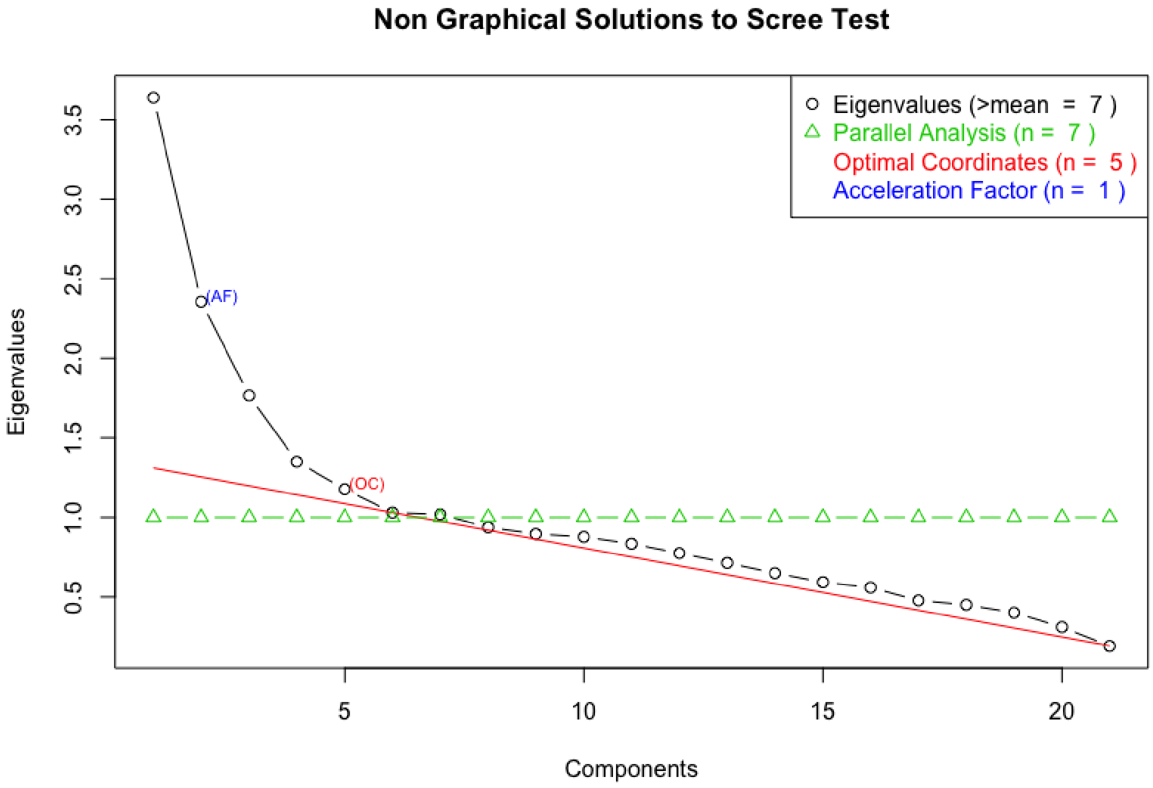

**Supplementary Figure 2. Exploratory factor analysis of trait-specific GRS.**

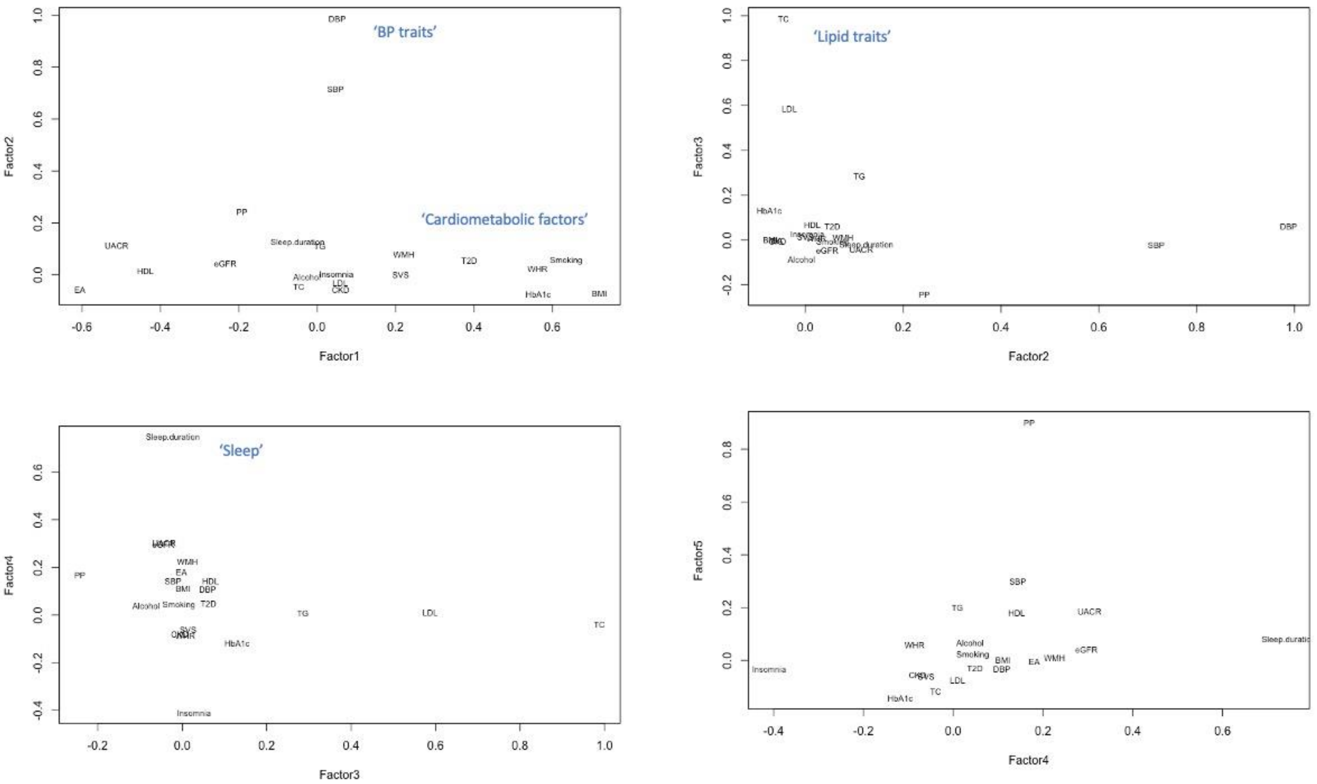

**Supplementary Figure 3. Weights and GRS included in the different versions of the metaGRS.** Estimates from the 'All-trait metaGRS', 'Causal metaGRS', and 'Factor metaGRS' are from inverse-variance random-effects meta-analysis across the training datasets. Estimates from 'Stepwise metaGRS' and 'Lasso metaGRS' are from merged training datasets. Estimates for the 'All-trait metaGRS', 'Causal metaGRS' and 'Factor metaGRS' are adjusted for age, sex, and two principal components.

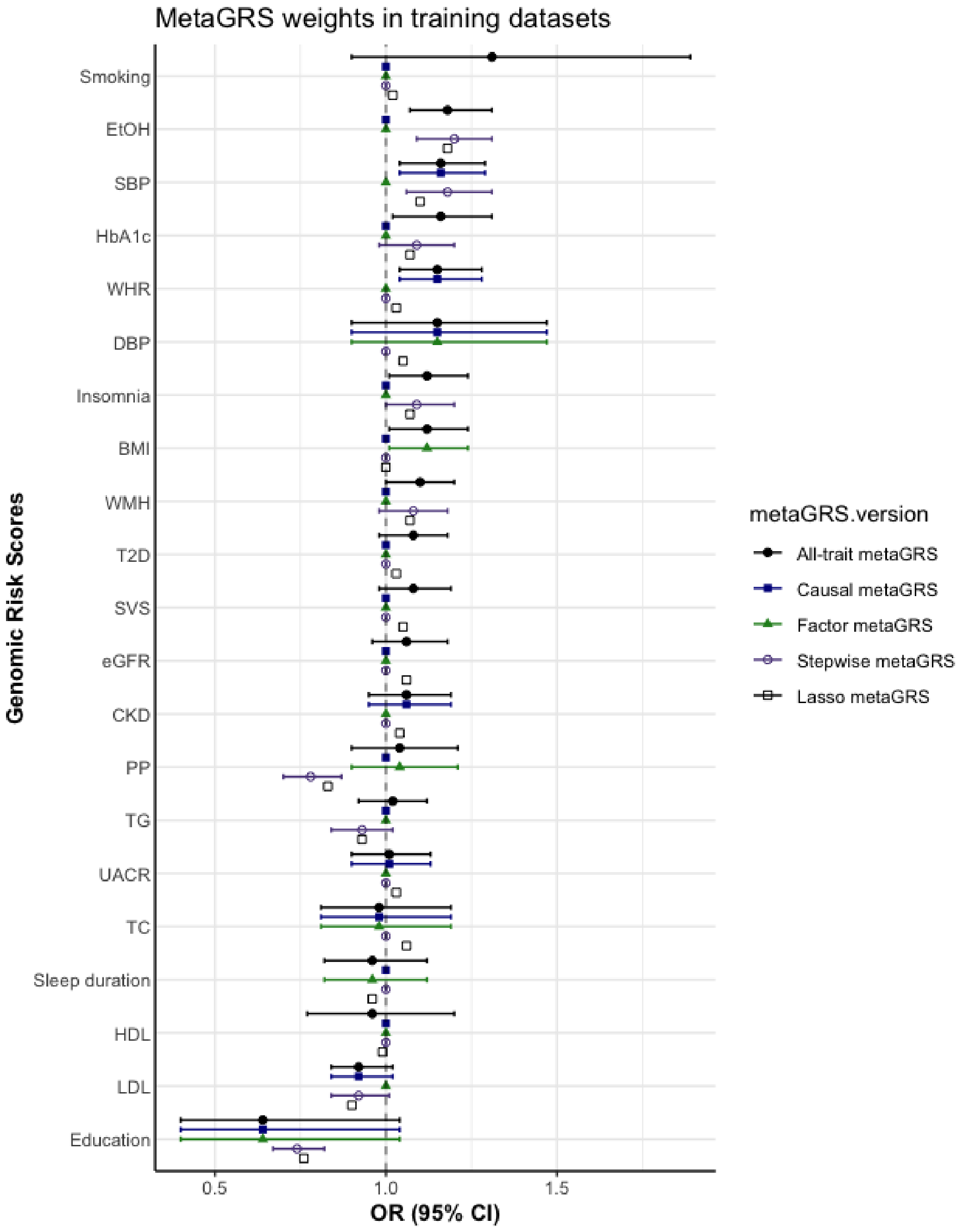

SBP systolic blood pressure; DBP diastolic blood pressure; PP pulse pressure; WMH white matter hyperintensities; CKD chronic kidney disease; eGFR estimated glomerular filtration rate; UACR urine albumin-to-creatinine ratio; TC total cholesterol; TG triglycerides; LDL low-density lipoprotein; HDL high-density lipoprotein; T2D type 2 diabetes mellitus; HbA1c hemoglobin A1c; BMI body mass index; WHR waist-to-hip ratio; SVS small vessel stroke; GRS genomic risk score.

**Supplementary Figure 4. Risk of ICH per metaGRS percentile groups, relative to the middle decile of the metaGRS distribution.**

Each effect size estimate and 95% CIs are from logistic regression models comparing the respective percentile groups (lower bound is inclusive, higher bound is exclusive) to the middle decile of the metaGRS distribution. All models are adjusted for age, sex, and two principal components.

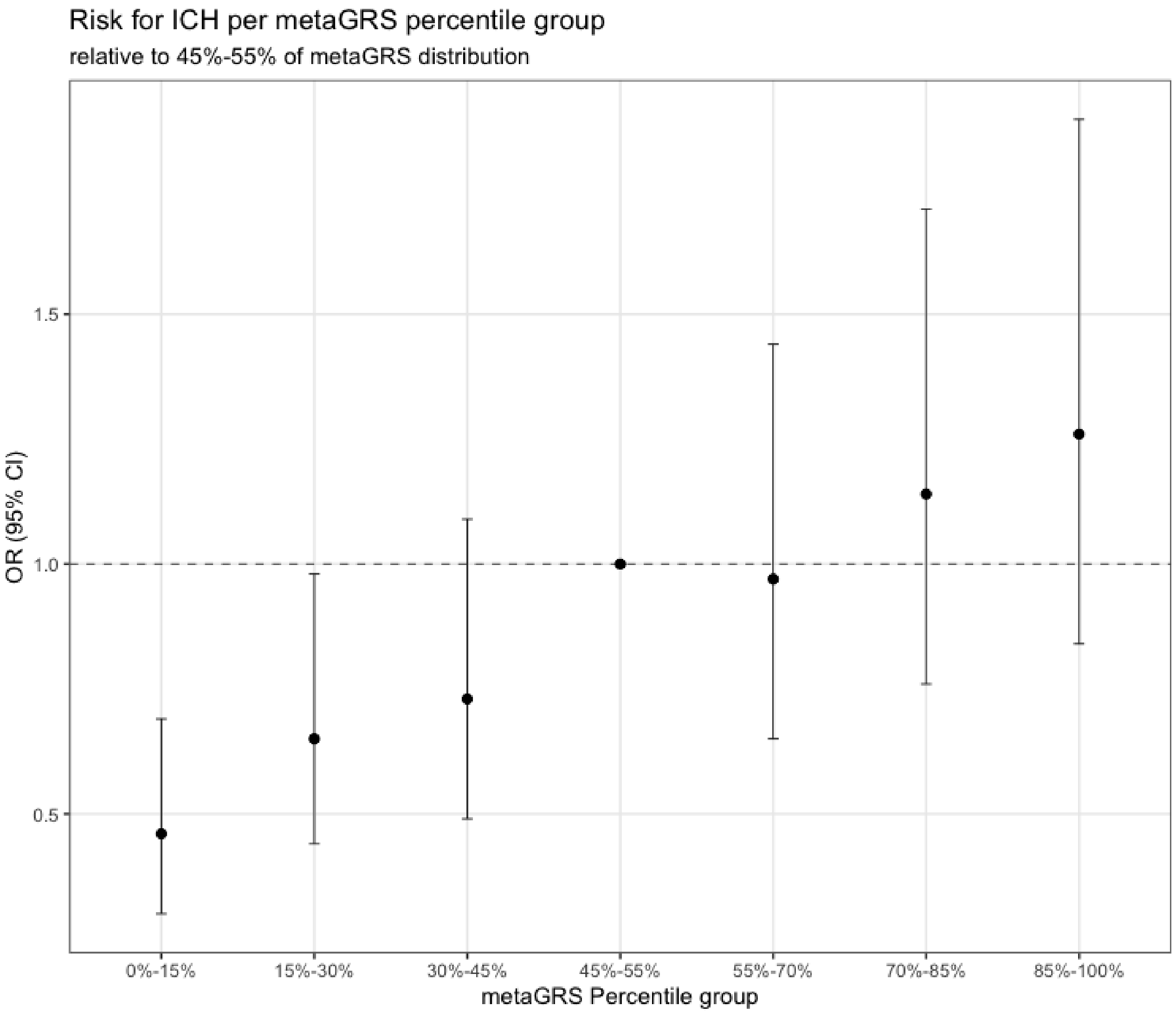

**Supplementary Figure 5. Risk of ICH per 5% sliding-widows metaGRS percentile groups, relative to the bottom 10% of the metaGRS distribution.**

Each effect size estimate is from logistic regression models comparing the 5% metaGRS percentile groups, relative to the lowest 0%-10% of the metaGRS distribution in a sliding-window fashion (11%-16% vs 0%-10%, 12%-17% vs 0%-10% etc). All models are adjusted for age, sex, and two principal components.

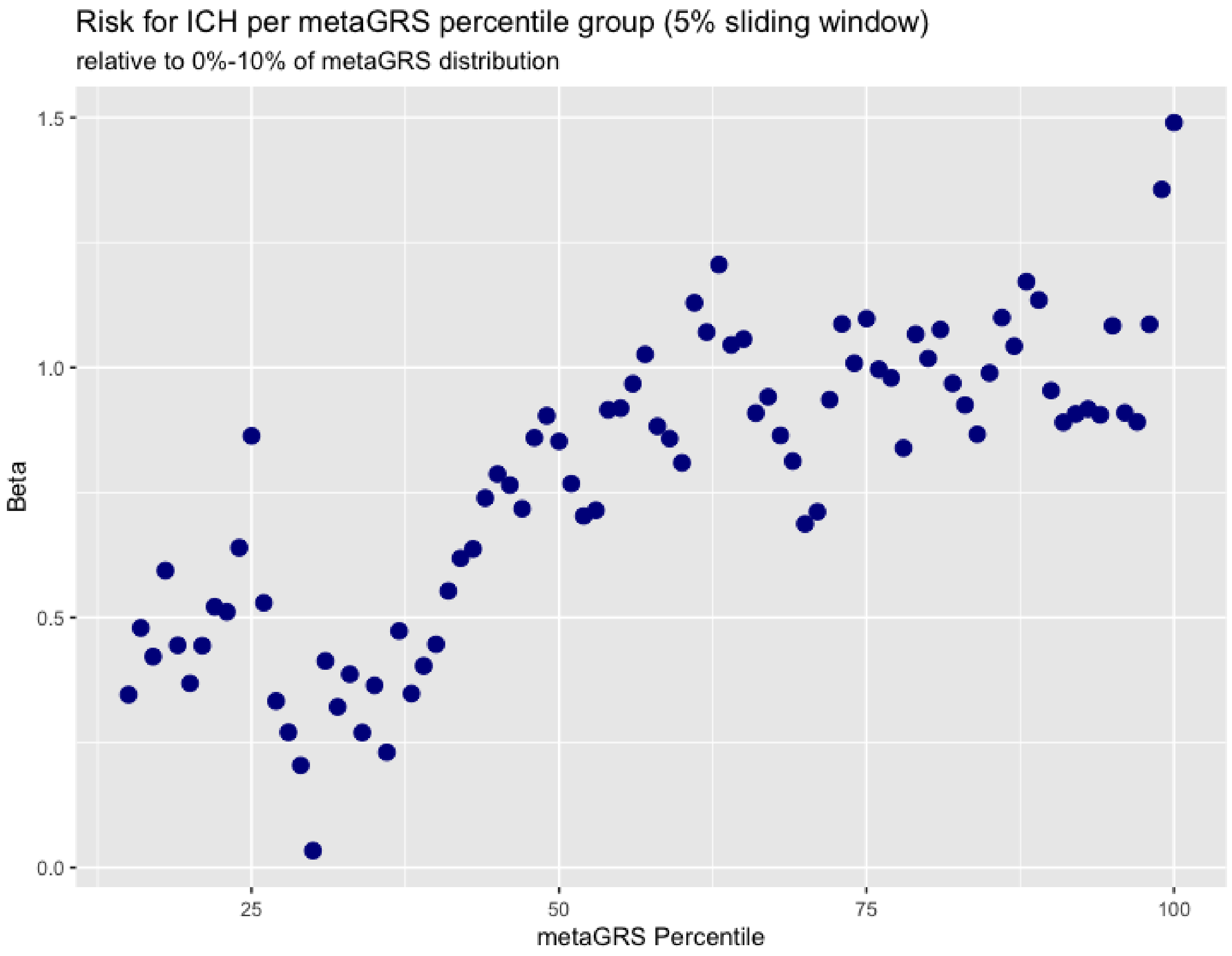

**Supplementary Figure 6. Comparison of (A) odds ratios and (B) c-indices of the different versions of the metaGRS in the validation dataset.**

All estimates in panel A are adjusted for age, sex, and two PCs. For panel B, 95% CIs for the c-indices are calculated after bootstrapping over 1000 iterations. The baseline ICH risk model for the c-index comparison contains only age, sex, and two PCs (c-index= 0.5291).

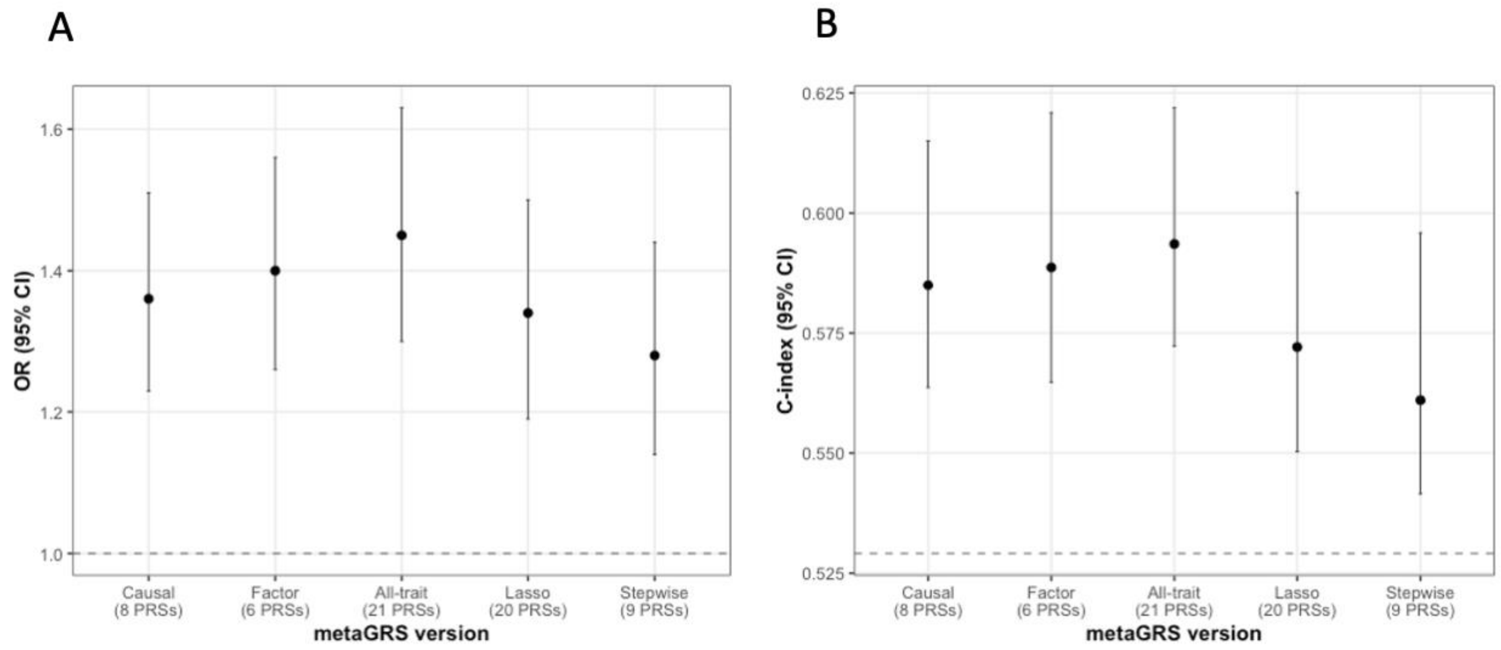

**Supplementary Figure 7. A-E. Comparison of predictive performance of different metaGRS versions with ICH clinical risk factors and with model containing clinical risk factors and metaGRS.**

**A. All-trait metaGRS**

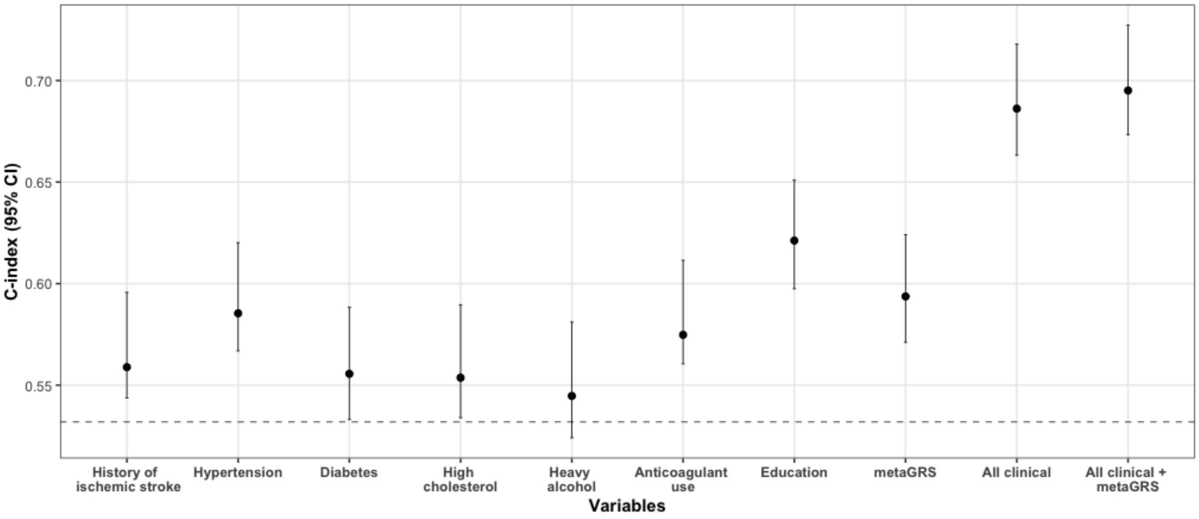

**B. Causal metaGRS**

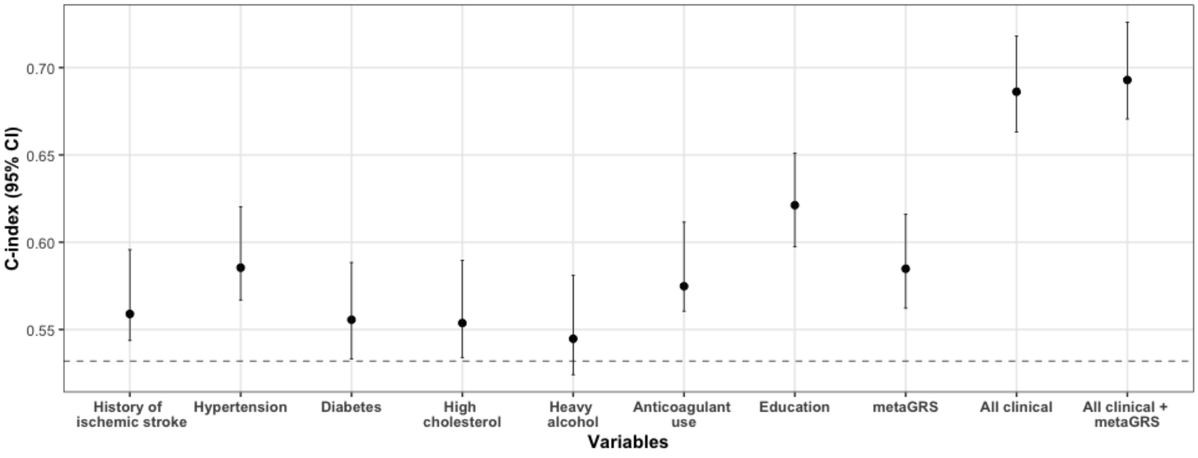

### C. Factor metaGRS

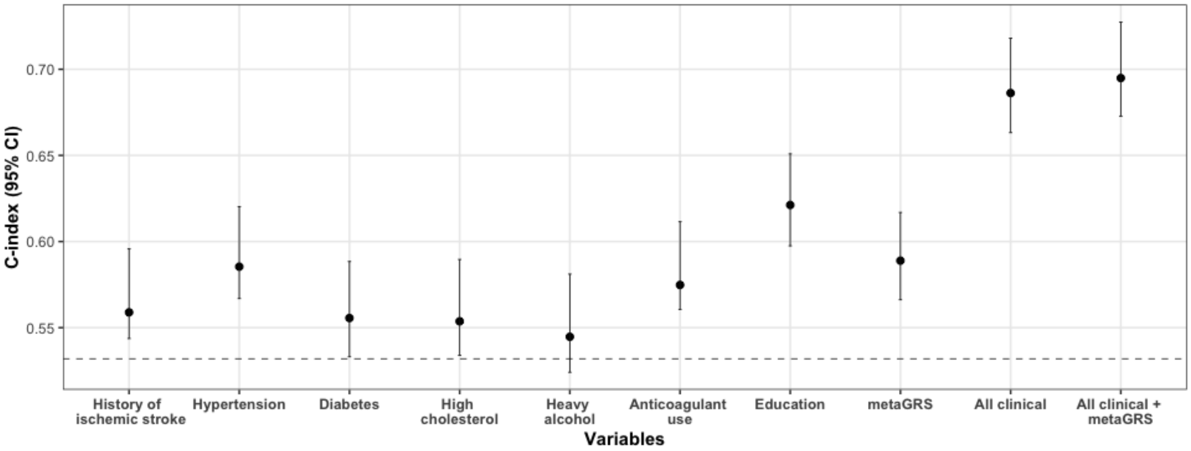

### D. Stepwise metaGRS

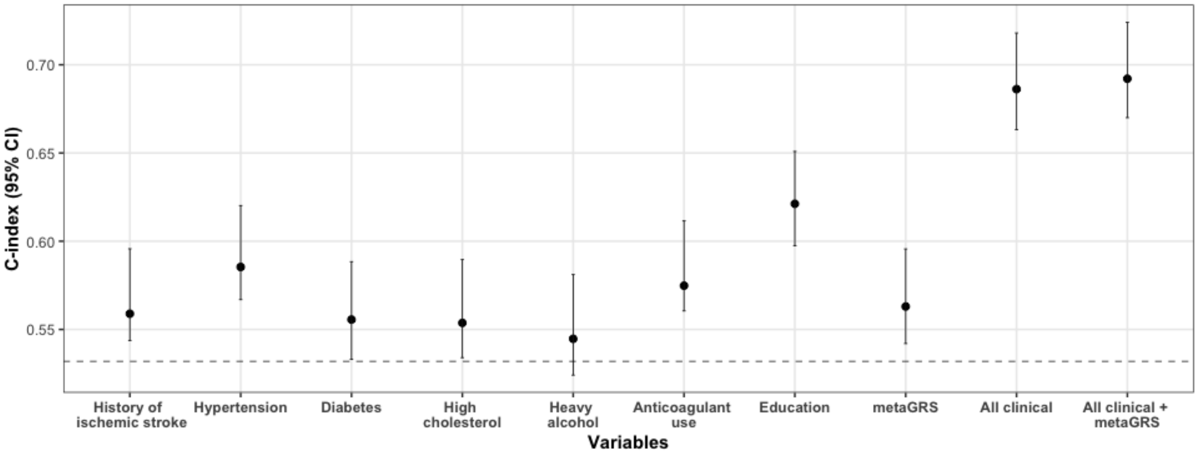

### E. Lasso metaGRS

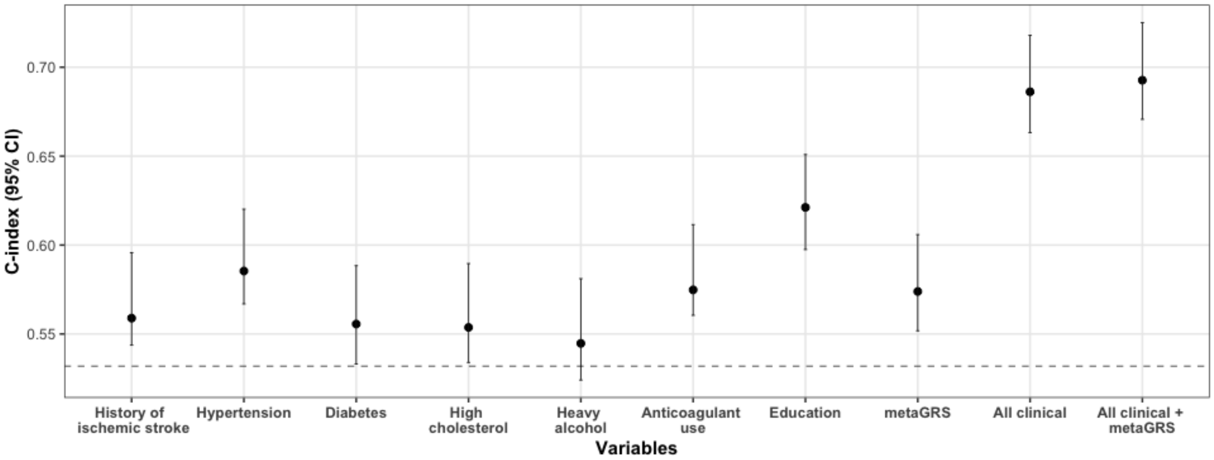

### Supplemental Tables and supporting information

**Supplementary Table 1. Description of GWAS samples used for trait-specific GRS construction.**

| **Phenotypes** | **Source** | **N (Total or Cases/Controls)** | **Reference** |
|---|---|---|---|
| SBP/DBP/PP | UK Biobank and ICBP | 757,601 | *Evangelou E et al. 2018* |
| Type 2 diabetes mellitus | DIAGRAM Consortium | 74,124/824,006 | *Mahajan A et al. 2017* |
| HbA1c | MAGIC Consortium | 123,665 | *Wheeler E et al. 2017* |
| TC/HDL/LDL/TG | Global Lipids Genetics Consortium | 94,595 | *Willer CJ et al. 2013* |
| BMI | UK Biobank and GIANT Consortium | 806,834 | *Pulit SL et al. 2019* |
| WHR | UK Biobank and GIANT Consortium | 697,734 | *Pulit SL et al. 2019* |
| CKD | CKDGen Consortium | 41,395/439,303 | *Wuttke M et al. 2019* |
| eGFR | CKDGen Consortium | 567,460 | *Wuttke M et al. 2019* |
| UACR | CKDGen Consortium | 547,361 | *Teumer A et al. 2019* |
| WMH | UK Biobank | 18,381 | *Persyn E et al. 2020* |
| SVS | MEGASTROKE | 5,386/406,111 | *Malik R et al. 2018* |
| Educational attainment | Social Science Genetic Association Consortium | 766,345 | *Lee JJ et al. 2018* |
| Alcohol consumption | GWAS and Sequencing Consortium of Alcohol and Nicotine use | 941,280 | *Liu M et al. 2019* |
| Smoking | UK Biobank | 462,690 | *Wootton RE et al. 2019* |
| Insomnia | UK Biobank | 453,379 | *Lane JM et al. 2019* |
| Sleep duration | UK Biobank | 446,118 | *Dashti HS et al. 2019* |

eGFR GWAS units: log-transformed eGFR. WMH GWAS units: logarithmic transformation and normalization by intracranial volume. Smoking GWAS units: lifetime smoking index. Alcohol consumption GWAS units: drinks per week. Data from 23andMe were excluded from the respective GWASs.

SBP systolic blood pressure; DBP diastolic blood pressure; PP pulse pressure; WMH white matter hyperintensities; CKD chronic kidney disease; eGFR estimated glomerular filtration rate; UACR urine albumin-to-creatinine ratio; TC total cholesterol; TG triglycerides; LDL low-density lipoprotein; HDL high-density lipoprotein; T2D type 2 diabetes mellitus; HbA1c hemoglobin A1c; BMI body mass index; WHR waist-to-hip ratio; SVS small vessel stroke; GRS genomic risk score

**Supplementary Tables 2-22. Selection of trait-specific GRS in training datasets.**

AUCs are from merged GOCHA and EUR/ISGC training datasets. ORs and 95% CIs are from merged GOCHA and EUR/ISGC training datasets. The final GRS with the largest AUC for each trait (see methods) is indicated with bold.

SBP systolic blood pressure; DBP diastolic blood pressure; PP pulse pressure; WMH white matter hyperintensities; CKD chronic kidney disease; eGFR estimated glomerular filtration rate; UACR urine albumin-to-creatinine ratio; TC total cholesterol; TG triglycerides; LDL low-density lipoprotein; HDL high-density lipoprotein; T2D type 2 diabetes mellitus; HbA1c hemoglobin A1c; BMI body mass index; WHR waist-to-hip ratio; SVS small vessel stroke; GRS genomic risk score; AUC are under the receiving-operating characteristics curve

| **Supplementary table 2. Selection of SBP GRS** | | | |
|---|---|---|---|
| **$r^2$** | **P-value threshold** | **OR per SD** | **AUC** |
| 0.3 | $1x10^{-8}$ | 1.08 (0.99-1.18) | 0.5696 |
| 0.3 | $5x10^{-8}$ | 1.09 (1.00-1.20) | 0.571 |
| 0.3 | $1x10^{-7}$ | 1.09 (1.00-1.20) | 0.5715 |
| 0.3 | $5x10^{-7}$ | 1.09 (1.00-1.20) | 0.571 |
| 0.3 | $1x10^{-6}$ | 1.09 (1.00-1.19) | 0.5706 |
| 0.3 | $5x10^{-6}$ | 1.08 (0.98-1.18) | 0.5694 |
| 0.3 | $1x10^{-5}$ | 1.08 (0.99-1.19) | 0.57 |
| 0.3 | $5x10^{-5}$ | 1.07 (0.98-1.17) | 0.5688 |
| 0.3 | $1x10^{-4}$ | 1.07 (0.98-1.17) | 0.5684 |
| 0.3 | $5x10^{-4}$ | 1.05 (0.96-1.15) | 0.5673 |
| 0.3 | $1x10^{-3}$ | 1.04 (0.95-1.14) | 0.567 |
| 0.3 | $5x10^{-3}$ | 1.01 (0.92-1.10) | 0.5666 |
| 0.3 | $1x10^{-2}$ | 1.00 (0.90-1.09) | 0.5666 |
| 0.3 | $5x10^{-2}$ | 0.96 (0.87-1.06) | 0.567 |
| 0.3 | 0.1 | 0.95 (0.86-1.04) | 0.5675 |
| 0.3 | 0.2 | 0.92 (0.84-1.02) | 0.5689 |
| 0.3 | 0.4 | 0.90 (0.82-0.99) | 0.5712 |
| 0.3 | 0.6 | 0.90 (0.81-0.99) | 0.5715 |
| 0.3 | 0.8 | 0.90 (0.81-0.99) | 0.5715 |
| 0.3 | 1.0 | 0.90 (0.81-0.98) | 0.5716 |
| 0.5 | $1x10^{-8}$ | 1.09 (0.99-1.19) | 0.5701 |
| 0.5 | $5x10^{-8}$ | 1.09 (1.00-1.20) | 0.5709 |
| 0.5 | $1x10^{-7}$ | 1.10 (1.00-1.20) | 0.5711 |
| 0.5 | $5x10^{-7}$ | 1.09 (1.00-1.19) | 0.5705 |
| 0.5 | $1x10^{-6}$ | 1.09 (1.00-1.19) | 0.5705 |

| | | | |
|---|---|---|---|
| 0.5 | $5x10^{-6}$ | 1.08 (0.99-1.19) | 0.5701 |
| 0.5 | $1x10^{-5}$ | 1.09 (0.99-1.19) | 0.5702 |
| 0.5 | $5x10^{-5}$ | 1.08 (0.98-1.18) | 0.5692 |
| 0.5 | $1x10^{-4}$ | 1.07 (0.98-1.17) | 0.5686 |
| 0.5 | $5x10^{-4}$ | 1.06 (0.97-1.16) | 0.5679 |
| 0.5 | $1x10^{-3}$ | 1.05 (0.96-1.15) | 0.5674 |
| 0.5 | $5x10^{-3}$ | 1.03 (0.94-1.12) | 0.5666 |
| 0.5 | $1x10^{-2}$ | 1.01 (0.92-1.11) | 0.5667 |
| 0.5 | $5x10^{-2}$ | 0.97 (0.89-1.07) | 0.5665 |
| 0.5 | 0.1 | 0.96 (0.87-1.06) | 0.5669 |
| 0.5 | 0.2 | 0.94 (0.85-1.03) | 0.5679 |
| 0.5 | 0.4 | 0.91 (0.83-1.01) | 0.5698 |
| 0.5 | 0.6 | 0.91 (0.83-1.00) | 0.5699 |
| 0.5 | 0.8 | 0.91 (0.83-1.00) | 0.57 |
| 0.5 | 1.0 | 0.91 (0.83-1.00) | 0.57 |
| 0.1 | $1x10^{-8}$ | 1.08 (0.98-1.18) | 0.5698 |
| 0.1 | $5x10^{-8}$ | 1.09 (1.00-1.20) | 0.571 |
| 0.1 | $1x10^{-7}$ | 1.10 (1.01-1.21) | 0.5727 |
| 0.1 | $5x10^{-7}$ | 1.11 (1.01-1.21) | 0.5726 |
| 0.1 | $1x10^{-6}$ | 1.11 (1.01-1.21) | 0.5729 |
| 0.1 | $5x10^{-6}$ | 1.11 (1.01-1.21) | 0.5727 |
| **0.1** | **$1x10^{-5}$** | **1.11 (1.01-1.22)** | **0.5734** |
| 0.1 | $5x10^{-5}$ | 1.10 (1.00-1.20) | 0.571 |
| 0.1 | $1x10^{-4}$ | 1.09 (0.99-1.19) | 0.5697 |
| 0.1 | $5x10^{-4}$ | 1.05 (0.96-1.15) | 0.5669 |
| 0.1 | $1x10^{-3}$ | 1.04 (0.94-1.14) | 0.5666 |
| 0.1 | $5x10^{-3}$ | 0.99 (0.90-1.09) | 0.5669 |
| 0.1 | $1x10^{-2}$ | 0.97 (0.88-1.06) | 0.567 |
| 0.1 | $5x10^{-2}$ | 0.93 (0.85-1.03) | 0.5681 |
| 0.1 | 0.1 | 0.92 (0.83-1.01) | 0.5689 |
| 0.1 | 0.2 | 0.90 (0.82-0.99) | 0.5711 |
| 0.1 | 0.4 | 0.89 (0.81-0.98) | 0.5729 |
| 0.1 | 0.6 | 0.89 (0.81-0.98) | 0.5731 |
| 0.1 | 0.8 | 0.89 (0.81-0.98) | 0.573 |
| 0.1 | 1.0 | 0.89 (0.81-0.98) | 0.573 |

| Supplementary table 3. Selection of DBP GRS | | | |
|---|---|---|---|
| $r^2$ | P-value threshold | OR per SD | AUC |
| 0.3 | $1x10^{-8}$ | 1.11 (1.01-1.21) | 0.572 |
| 0.3 | $5x10^{-8}$ | 1.12 (1.02-1.23) | 0.574 |
| 0.3 | $1x10^{-7}$ | 1.12 (1.02-1.23) | 0.5734 |
| 0.3 | $5x10^{-7}$ | 1.11 (1.01-1.22) | 0.5725 |
| 0.3 | $1x10^{-6}$ | 1.10 (1.01-1.21) | 0.5716 |
| 0.3 | $5x10^{-6}$ | 1.10 (1.01-1.21) | 0.572 |
| 0.3 | $1x10^{-5}$ | 1.10 (1.01-1.21) | 0.5717 |
| 0.3 | $5x10^{-5}$ | 1.11 (1.01-1.21) | 0.5728 |
| 0.3 | $1x10^{-4}$ | 1.10 (1.00-1.20) | 0.5722 |
| 0.3 | $5x10^{-4}$ | 1.09 (1.00-1.19) | 0.5711 |
| 0.3 | $1x10^{-3}$ | 1.09 (0.99-1.19) | 0.5708 |
| 0.3 | $5x10^{-3}$ | 1.09 (0.99-1.19) | 0.5704 |
| 0.3 | $1x10^{-2}$ | 1.06 (0.97-1.16) | 0.5679 |
| 0.3 | $5x10^{-2}$ | 1.03 (0.94-1.13) | 0.5669 |
| 0.3 | 0.1 | 1.00 (0.91-1.09) | 0.5666 |
| 0.3 | 0.2 | 0.98 (0.89-1.07) | 0.5674 |
| 0.3 | 0.4 | 0.96 (0.87-1.05) | 0.5685 |
| 0.3 | 0.6 | 0.95 (0.87-1.04) | 0.569 |
| 0.3 | 0.8 | 0.95 (0.87-1.04) | 0.5688 |
| 0.3 | 1.0 | 0.95 (0.87-1.04) | 0.5688 |
| 0.5 | $1x10^{-8}$ | 1.11 (1.01-1.21) | 0.5718 |
| 0.5 | $5x10^{-8}$ | 1.12 (1.02-1.22) | 0.5729 |
| 0.5 | $1x10^{-7}$ | 1.12 (1.02-1.22) | 0.5727 |
| 0.5 | $5x10^{-7}$ | 1.11 (1.01-1.22) | 0.5725 |
| 0.5 | $1x10^{-6}$ | 1.11 (1.01 -1.21) | 0.572 |
| 0.5 | $5x10^{-6}$ | 1.11 (1.01-1.21) | 0.5722 |
| 0.5 | $1x10^{-5}$ | 1.10 (1.01-1.21) | 0.572 |
| 0.5 | $5x10^{-5}$ | 1.11 (1.01-1.21) | 0.5728 |
| 0.5 | $1x10^{-4}$ | 1.10 (1.01-1.21) | 0.5725 |
| 0.5 | $5x10^{-4}$ | 1.10 (1.01-1.21) | 0.5725 |
| 0.5 | $1x10^{-3}$ | 1.10 (1.00-1.20) | 0.5717 |
| 0.5 | $5x10^{-3}$ | 1.09 (1.00-1.19) | 0.5711 |
| 0.5 | $1x10^{-2}$ | 1.07 (0.98-1.18) | 0.5691 |
| 0.5 | $5x10^{-2}$ | 1.04 (0.95-1.14) | 0.5671 |
| 0.5 | 0.1 | 1.01 (0.92-1.11) | 0.5666 |
| 0.5 | 0.2 | 1.00 (0.91-1.09) | 0.5667 |
| 0.5 | 0.4 | 0.97 (0.89-1.07) | 0.5675 |
| 0.5 | 0.6 | 0.97 (0.88-1.06) | 0.5677 |
| 0.5 | 0.8 | 0.97 (0.89-1.06) | 0.5675 |
| 0.5 | 1.0 | 0.97 (0.89-1.06) | 0.5676 |
| 0.1 | $1x10^{-8}$ | 1.10 (1.01-1.21) | 0.5704 |
| 0.1 | $5x10^{-8}$ | 1.11 (1.01-1.21) | 0.5706 |
| 0.1 | $1x10^{-7}$ | 1.11 (1.02-1.22) | 0.5713 |
| 0.1 | $5x10^{-7}$ | 1.12 (1.02-1.22) | 0.5722 |
| 0.1 | $1x10^{-6}$ | 1.10 (1.01-1.21) | 0.5707 |
| 0.1 | $5x10^{-6}$ | 1.12 (1.02-1.21) | 0.573 |
| 0.1 | $1x10^{-5}$ | 1.11 (1.02-1.22) | 0.5721 |
| **0.1** | **$5x10^{-5}$** | **1.13 (1.04-1.24)** | **0.5752** |
| 0.1 | $1x10^{-4}$ | 1.12 (1.02-1.23) | 0.5742 |
| 0.1 | $5x10^{-4}$ | 1.10 (1.00-1.20) | 0.5718 |

| | | | |
|---|---|---|---|
| 0.1 | $1x10^{-3}$ | 1.10 (1.00-1.20) | 0.5711 |
| 0.1 | $5x10^{-3}$ | 1.06 (0.96-1.16) | 0.5675 |
| 0.1 | $1x10^{-2}$ | 1.01 (0.93-1.11) | 0.5663 |
| 0.1 | $5x10^{-2}$ | 0.98 (0.90-1.08) | 0.5672 |
| 0.1 | 0.1 | 0.95 (0.87-1.04) | 0.5693 |
| 0.1 | 0.2 | 0.94 (0.85-1.02) | 0.5705 |
| 0.1 | 0.4 | 0.92 (0.84-1.01) | 0.5722 |
| 0.1 | 0.6 | 0.92 (0.84-1.01) | 0.5724 |
| 0.1 | 0.8 | 0.92 (0.84-1.01) | 0.5723 |
| 0.1 | 1.0 | 0.92 (0.84-1.01) | 0.5723 |

| Supplementary table 4. Selection of PP GRS | | | |
|---|---|---|---|
| **$r^2$** | **P-value threshold** | **OR per SD** | **AUC** |
| 0.3 | $1x10^{-8}$ | 1.00 (0.91-1.19) | 0.5666 |
| 0.3 | $5x10^{-8}$ | 1.01 (0.92-1.11) | 0.5666 |
| 0.3 | $1x10^{-7}$ | 1.01 (0.92-1.11) | 0.5666 |
| 0.3 | $5x10^{-7}$ | 1.01 (0.92-1.10) | 0.5665 |
| 0.3 | $1x10^{-6}$ | 1.01 (0.92-1.10) | 0.5665 |
| 0.3 | $5x10^{-6}$ | 1.01 (0.92-1.10) | 0.5664 |
| 0.3 | $1x10^{-5}$ | 0.99 (0.90-1.09) | 0.5666 |
| 0.3 | $5x10^{-5}$ | 0.99 (0.91-1.09) | 0.5666 |
| 0.3 | $1x10^{-4}$ | 1.00 (0.91-1.10) | 0.5666 |
| 0.3 | $5x10^{-4}$ | 0.99 (0.90-1.08) | 0.5665 |
| 0.3 | $1x10^{-3}$ | 0.97 (0.89-1.07) | 0.5666 |
| 0.3 | $5x10^{-3}$ | 0.94 (0.86-1.03) | 0.5678 |
| 0.3 | $1x10^{-2}$ | 0.94 (0.86-1.03) | 0.5678 |
| 0.3 | $5x10^{-2}$ | 0.92 (0.83-1.01) | 0.5688 |
| 0.3 | 0.1 | 0.90 (0.81-0.99) | 0.5703 |
| 0.3 | 0.2 | 0.88 (0.80-0.98) | 0.5712 |
| 0.3 | 0.4 | 0.89 (0.81-0.98) | 0.5701 |
| 0.3 | 0.6 | 0.88 (0.80-0.97) | 0.5717 |
| 0.3 | 0.8 | 0.88 (0.80-0.97) | 0.5716 |
| 0.3 | 1.0 | 0.88 (0.80-0.97) | 0.5714 |
| 0.5 | $1x10^{-8}$ | 1.03 (0.94-1.13) | 0.5671 |
| 0.5 | $5x10^{-8}$ | 1.03 (0.94-1.13) | 0.5668 |
| 0.5 | $1x10^{-7}$ | 1.03 (0.94-1.13) | 0.567 |
| 0.5 | $5x10^{-7}$ | 1.02 (0.93-1.12) | 0.5668 |
| 0.5 | $1x10^{-6}$ | 1.01 (0.93-1.11) | 0.5666 |
| 0.5 | $5x10^{-6}$ | 1.02 (0.93-1.11) | 0.5667 |
| 0.5 | $1x10^{-5}$ | 1.01 (0.92-1.11) | 0.5665 |
| 0.5 | $5x10^{-5}$ | 1.00 (0.92-1.10) | 0.5665 |
| 0.5 | $1x10^{-4}$ | 1.01 (0.92-1.11) | 0.5665 |
| 0.5 | $5x10^{-4}$ | 1.00 (0.91-1.09) | 0.5665 |
| 0.5 | $1x10^{-3}$ | 0.98 (0.89-1.08) | 0.5665 |
| 0.5 | $5x10^{-3}$ | 0.96 (0.87-1.05) | 0.5671 |
| 0.5 | $1x10^{-2}$ | 0.95 (0.86-1.04) | 0.5674 |
| 0.5 | $5x10^{-2}$ | 0.91 (0.83-1.00) | 0.5698 |
| 0.5 | 0.1 | 0.89 (0.81-0.98) | 0.5712 |
| 0.5 | 0.2 | 0.89 (0.80-0.98) | 0.5715 |
| 0.5 | 0.4 | 0.89 (0.81-0.98) | 0.5707 |
| 0.5 | 0.6 | 0.88 (0.80-0.97) | 0.5718 |
| 0.5 | 0.8 | 0.88 (0.80-0.97) | 0.5717 |
| 0.5 | 1.0 | 0.88 (0.80-0.97) | 0.5717 |
| 0.1 | $1x10^{-8}$ | 0.99 (0.91-1.09) | 0.5666 |
| 0.1 | $5x10^{-8}$ | 0.98 (0.90-1.08) | 0.5667 |
| 0.1 | $1x10^{-7}$ | 0.99 (0.91-1.09) | 0.5665 |
| 0.1 | $5x10^{-7}$ | 0.99 (0.90-1.08) | 0.5667 |
| 0.1 | $1x10^{-6}$ | 0.98 (0.90-1.08) | 0.5669 |
| 0.1 | $5x10^{-6}$ | 0.98 (0.89-1.07) | 0.5668 |
| 0.1 | $1x10^{-5}$ | 0.97 (0.88-1.06) | 0.5676 |
| 0.1 | $5x10^{-5}$ | 0.95 (0.87-1.05) | 0.5682 |
| 0.1 | $1x10^{-4}$ | 0.98 (0.89-1.07) | 0.5668 |
| 0.1 | $5x10^{-4}$ | 0.97 (0.88-1.06) | 0.5668 |

| | | | |
|---|---|---|---|
| 0.1 | $1x10^{-3}$ | 0.95 (0.86-1.04) | 0.5674 |
| 0.1 | $5x10^{-3}$ | 0.91 (0.83-1.00) | 0.5699 |
| 0.1 | $1x10^{-2}$ | 0.90 (0.81-0.99) | 0.5712 |
| 0.1 | $5x10^{-2}$ | 0.90 (0.81-0.99) | 0.5706 |
| 0.1 | 0.1 | 0.89 (0.80-0.97) | 0.572 |
| 0.1 | 0.2 | 0.87 (0.78-0.96) | 0.5741 |
| 0.1 | 0.4 | 0.88 (0.79-0.97) | 0.5716 |
| 0.1 | 0.6 | 0.86 (0.78-0.95) | 0.5745 |
| **0.1** | **0.8** | **0.86 (0.78-0.95)** | **0.5748** |
| 0.1 | 1.0 | 0.86 (0.78-0.95) | 0.5748 |

| **Supplementary table 5. Selection of WMH GRS** | | | |
|---|---|---|---|
| **$r^2$** | **P-value threshold** | **OR per SD** | **AUC** |
| 0.3 | $1x10^{-8}$ | 1.04 (0.95-1.14) | 0.5674 |
| 0.3 | $5x10^{-8}$ | 1.07 (0.98-1.17) | 0.5677 |
| 0.3 | $1x10^{-7}$ | 1.07 (0.98-1.17) | 0.5676 |
| 0.3 | $5x10^{-7}$ | 1.05 (0.96-1.15) | 0.5669 |
| 0.3 | $1x10^{-6}$ | 1.04 (0.95-1.14) | 0.5669 |
| 0.3 | $5x10^{-6}$ | 1.04 (0.95-1.14) | 0.5668 |
| 0.3 | $1x10^{-5}$ | 1.03 (0.94-1.13) | 0.5662 |
| 0.3 | $5x10^{-5}$ | 1.09 (0.99-1.19) | 0.5684 |
| 0.3 | $1x10^{-4}$ | 1.03 (0.94-1.13) | 0.5664 |
| 0.3 | $5x10^{-4}$ | 0.99 (0.90-1.08) | 0.5668 |
| 0.3 | $1x10^{-3}$ | 0.99 (0.91-1.09) | 0.5665 |
| 0.3 | $5x10^{-3}$ | 1.01 (0.93-1.11) | 0.5663 |
| 0.3 | $1x10^{-2}$ | 1.02 (0.93-1.11) | 0.5662 |
| 0.3 | $5x10^{-2}$ | 1.11 (1.01-1.21) | 0.5717 |
| 0.3 | 0.1 | 1.07 (0.98-1.17) | 0.5685 |
| 0.3 | 0.2 | 1.08 (0.99-1.18) | 0.5693 |
| 0.3 | 0.4 | 1.09 (0.99-1.19) | 0.5697 |
| 0.3 | 0.6 | 1.09 (1.00-1.19) | 0.5699 |
| 0.3 | 0.8 | 1.09 (1.00-1.20) | 0.57 |
| 0.3 | 1.0 | 1.09 (1.00-1.20) | 0.5703 |
| 0.5 | $1x10^{-8}$ | 1.04 (0.95-1.13) | 0.5667 |
| 0.5 | $5x10^{-8}$ | 1.06 (0.96-1.16) | 0.5672 |
| 0.5 | $1x10^{-7}$ | 1.06 (0.96-1.16) | 0.5673 |
| 0.5 | $5x10^{-7}$ | 1.03 (0.95-1.13) | 0.5663 |
| 0.5 | $1x10^{-6}$ | 1.04 (0.95-1.14) | 0.5667 |
| 0.5 | $5x10^{-6}$ | 1.04 (0.95-1.14) | 0.5667 |
| 0.5 | $1x10^{-5}$ | 1.04 (0.95-1.13) | 0.5663 |
| 0.5 | $5x10^{-5}$ | 1.08 (0.99-1.19) | 0.5687 |
| 0.5 | $1x10^{-4}$ | 1.04 (0.95-1.14) | 0.5669 |
| 0.5 | $5x10^{-4}$ | 1.01 (0.92-1.10) | 0.5664 |
| 0.5 | $1x10^{-3}$ | 1.02 (0.94-1.12) | 0.567 |
| 0.5 | $5x10^{-3}$ | 1.02 (0.94-1.12) | 0.5663 |
| 0.5 | $1x10^{-2}$ | 1.01 (0.93-1.11) | 0.5664 |
| **0.5** | **$5x10^{-2}$** | **1.12 (1.02-1.23)** | **0.5733** |
| 0.5 | 0.1 | 1.08 (0.99-1.19) | 0.5693 |
| 0.5 | 0.2 | 1.09 (0.99-1.19) | 0.5694 |
| 0.5 | 0.4 | 1.09 (0.99-1.19) | 0.5697 |
| 0.5 | 0.6 | 1.09 (0.99-1.19) | 0.5696 |
| 0.5 | 0.8 | 1.09 (1.00-1.19) | 0.5702 |
| 0.5 | 1.0 | 1.09 (1.00-1.20) | 0.5704 |
| 0.1 | $1x10^{-8}$ | 1.06 (0.97-1.16) | 0.5681 |
| 0.1 | $5x10^{-8}$ | 1.09 (1.00-1.19) | 0.5707 |
| 0.1 | $1x10^{-7}$ | 1.09 (0.99-1.19) | 0.5703 |
| 0.1 | $5x10^{-7}$ | 1.06 (0.97-1.16) | 0.5682 |
| 0.1 | $1x10^{-6}$ | 1.05 (0.96-1.15) | 0.568 |
| 0.1 | $5x10^{-6}$ | 1.04 (0.95-1.13) | 0.5672 |
| 0.1 | $1x10^{-5}$ | 1.02 (0.93-1.11) | 0.5665 |
| 0.1 | $5x10^{-5}$ | 1.08 (0.99-1.19) | 0.5681 |
| 0.1 | $1x10^{-4}$ | 1.01 (0.92-1.11) | 0.5665 |
| 0.1 | $5x10^{-4}$ | 0.98 (0.90-1.08) | 0.5669 |

| | | | |
|---|---|---|---|
| 0.1 | $1x10^{-3}$ | 0.99 (0.90-1.08) | 0.5666 |
| 0.1 | $5x10^{-3}$ | 1.02 (0.93-1.11) | 0.5664 |
| 0.1 | $1x10^{-2}$ | 1.02 (0.93-1.12) | 0.5663 |
| 0.1 | $5x10^{-2}$ | 1.10 (1.01-1.21) | 0.572 |
| 0.1 | 0.1 | 1.05 (0.96-1.15) | 0.568 |
| 0.1 | 0.2 | 1.07 (0.98-1.17) | 0.569 |
| 0.1 | 0.4 | 1.07 (0.98-1.18) | 0.5696 |
| 0.1 | 0.6 | 1.09 (1.00-1.19) | 0.5709 |
| 0.1 | 0.8 | 1.09 (1.00-1.19) | 0.5705 |
| 0.1 | 1.0 | 1.09 (0.99-1.19) | 0.5707 |

| **Supplementary table 6. Selection of CKD GRS** | | | |
|---|---|---|---|
| **$r^2$** | **P-value threshold** | **OR per SD** | **AUC** |
| 0.3 | $1x10^{-8}$ | 1.02 (0.93-1.11) | 0.5666 |
| 0.3 | $5x10^{-8}$ | 1.01 (0.92-1.10) | 0.5664 |
| 0.3 | $1x10^{-7}$ | 1.02 (0.93-1.12) | 0.5667 |
| 0.3 | $5x10^{-7}$ | 1.03 (0.94-1.12) | 0.5665 |
| 0.3 | $1x10^{-6}$ | 1.03 (0.94-1.13) | 0.5667 |
| 0.3 | $5x10^{-6}$ | 1.07 (0.97-1.17) | 0.571 |
| 0.3 | $1x10^{-5}$ | 1.07 (0.97-1.17) | 0.5712 |
| 0.3 | $5x10^{-5}$ | 1.06 (0.97-1.17) | 0.5712 |
| 0.3 | $1x10^{-4}$ | 1.07 (0.97-1.17) | 0.5715 |
| 0.3 | $5x10^{-4}$ | 1.08 (0.98-1.18) | 0.5725 |
| 0.3 | $1x10^{-3}$ | 1.08 (0.98-1.18) | 0.5726 |
| 0.3 | $5x10^{-3}$ | 1.08 (0.98-1.18) | 0.573 |
| 0.3 | $1x10^{-2}$ | 1.08 (0.98-1.18) | 0.5728 |
| 0.3 | $5x10^{-2}$ | 1.07 (0.98-1.18) | 0.5725 |
| 0.3 | 0.1 | 1.07 (0.98-1.18) | 0.5723 |
| 0.3 | 0.2 | 1.07 (0.98-1.18) | 0.5724 |
| 0.3 | 0.4 | 1.07 (0.98-1.18) | 0.5724 |
| 0.3 | 0.6 | 1.07 (0.98-1.18) | 0.5724 |
| 0.3 | 0.8 | 1.07 (0.98-1.18) | 0.5724 |
| 0.3 | 1.0 | 1.07 (0.98-1.18) | 0.5724 |
| 0.5 | $1x10^{-8}$ | 1.06 (0.97-1.16) | 0.5681 |
| 0.5 | $5x10^{-8}$ | 1.05 (0.96-1.15) | 0.5678 |
| 0.5 | $1x10^{-7}$ | 1.06 (0.97-1.16) | 0.5686 |
| 0.5 | $5x10^{-7}$ | 1.06 (0.97-1.16) | 0.5684 |
| 0.5 | $1x10^{-6}$ | 1.06 (0.96-1.15) | 0.5683 |
| **0.5** | **$5x10^{-6}$** | **1.08 (0.99-1.19)** | **0.5731** |
| 0.5 | $1x10^{-5}$ | 1.08 (0.99-1.18) | 0.5727 |
| 0.5 | $5x10^{-5}$ | 1.07 (0.98-1.17) | 0.5718 |
| 0.5 | $1x10^{-4}$ | 1.07 (0.97-1.17) | 0.5715 |
| 0.5 | $5x10^{-4}$ | 1.08 (0.99-1.19) | 0.5729 |
| 0.5 | $1x10^{-3}$ | 1.08 (0.99-1.18) | 0.5727 |
| 0.5 | $5x10^{-3}$ | 1.08 (0.98-1.18) | 0.573 |
| 0.5 | $1x10^{-2}$ | 1.08 (0.99-1.18) | 0.5731 |
| 0.5 | $5x10^{-2}$ | 1.08 (0.98-1.18) | 0.5727 |
| 0.5 | 0.1 | 1.08 (0.98-1.18) | 0.5726 |
| 0.5 | 0.2 | 1.08 (0.98-1.18) | 0.5728 |
| 0.5 | 0.4 | 1.08 (0.98-1.18) | 0.5727 |
| 0.5 | 0.6 | 1.08 (0.98-1.18) | 0.5726 |
| 0.5 | 0.8 | 1.08 (0.98-1.18) | 0.5726 |
| 0.5 | 1.0 | 1.08 (0.98-1.18) | 0.5726 |
| 0.1 | $1x10^{-8}$ | 1.08 (0.99-1.18) | 0.5709 |
| 0.1 | $5x10^{-8}$ | 1.05 (0.96-1.15) | 0.5679 |
| 0.1 | $1x10^{-7}$ | 1.06 (0.97-1.16) | 0.5688 |
| 0.1 | $5x10^{-7}$ | 1.06 (0.97-1.16) | 0.5684 |
| 0.1 | $1x10^{-6}$ | 1.05 (0.96-1.15) | 0.5678 |
| 0.1 | $5x10^{-6}$ | 1.08 (0.98-1.18) | 0.5725 |
| 0.1 | $1x10^{-5}$ | 1.08 (0.98-1.18) | 0.5724 |
| 0.1 | $5x10^{-5}$ | 1.06 (0.97-1.17) | 0.5712 |
| 0.1 | $1x10^{-4}$ | 1.06 (0.97-1.17) | 0.5713 |
| 0.1 | $5x10^{-4}$ | 1.08 (0.98-1.18) | 0.5727 |

| | | | |
|---|---|---|---|
| 0.1 | $1x10^{-3}$ | 1.08 (0.98-1.18) | 0.5726 |
| 0.1 | $5x10^{-3}$ | 1.08 (0.98-1.18) | 0.573 |
| 0.1 | $1x10^{-2}$ | 1.08 (0.99-1.18) | 0.5731 |
| 0.1 | $5x10^{-2}$ | 1.07 (0.98-1.18) | 0.5725 |
| 0.1 | 0.1 | 1.07 (0.98-1.17) | 0.5722 |
| 0.1 | 0.2 | 1.07 (0.98-1.17) | 0.5722 |
| 0.1 | 0.4 | 1.07 (0.98-1.17) | 0.5722 |
| 0.1 | 0.6 | 1.07 (0.98-1.17) | 0.5722 |
| 0.1 | 0.8 | 1.07 (0.98-1.17) | 0.5722 |
| 0.1 | 1.0 | 1.07 (0.98-1.17) | 0.5722 |

| Supplementary table 7. Selection of GFR GRS | | | |
|---|---|---|---|
| **$r^2$** | **P-value threshold** | **OR per SD** | **AUC** |
| 0.3 | $1x10^{-8}$ | 1.00 (0.92-1.10) | 0.5666 |
| 0.3 | $5x10^{-8}$ | 1.02 (0.93-1.12) | 0.5666 |
| 0.3 | $1x10^{-7}$ | 1.02 (0.93-1.12) | 0.5667 |
| 0.3 | $5x10^{-7}$ | 1.03 (0.94-1.13) | 0.567 |
| 0.3 | $1x10^{-6}$ | 1.03 (0.94-1.12) | 0.567 |
| 0.3 | $5x10^{-6}$ | 1.02 (0.93-1.12) | 0.5668 |
| 0.3 | $1x10^{-5}$ | 1.02 (0.93-1.12) | 0.5668 |
| 0.3 | $5x10^{-5}$ | 1.04 (0.95-1.14) | 0.5677 |
| 0.3 | $1x10^{-4}$ | 1.06 (0.97-1.16) | 0.5688 |
| 0.3 | $5x10^{-4}$ | 1.05 (0.96-1.15) | 0.5675 |
| 0.3 | $1x10^{-3}$ | 1.04 (0.95-1.14) | 0.5662 |
| 0.3 | $5x10^{-3}$ | 1.06 (0.96-1.16) | 0.5672 |
| 0.3 | $1x10^{-2}$ | 1.01 (0.92-1.11) | 0.5662 |
| 0.3 | $5x10^{-2}$ | 0.97 (0.89-1.07) | 0.5681 |
| 0.3 | 0.1 | 0.97 (0.88-1.06) | 0.5679 |
| 0.3 | 0.2 | 0.95 (0.97-1.04) | 0.5698 |
| 0.3 | 0.4 | 0.95 (0.87-1.04) | 0.5703 |
| 0.3 | 0.6 | 0.95 (0.87-1.04) | 0.5704 |
| 0.3 | 0.8 | 0.95 (0.86-1.04) | 0.5705 |
| 0.3 | 1.0 | 0.95 (0.86-1.04) | 0.5705 |
| 0.5 | $1x10^{-8}$ | 0.99 (0.90-1.08) | 0.5667 |
| 0.5 | $5x10^{-8}$ | 1.00 (0.92-1.10) | 0.5666 |
| 0.5 | $1x10^{-7}$ | 1.01 (0.92-1.10) | 0.5665 |
| 0.5 | $5x10^{-7}$ | 1.02 (0.93-1.12) | 0.5665 |
| 0.5 | $1x10^{-6}$ | 1.02 (0.93-1.11) | 0.5664 |
| 0.5 | $5x10^{-6}$ | 1.03 (0.94-1.12) | 0.5667 |
| 0.5 | $1x10^{-5}$ | 1.03 (0.94-1.12) | 0.5667 |
| 0.5 | $5x10^{-5}$ | 1.03 (0.95-1.13) | 0.5674 |
| 0.5 | $1x10^{-4}$ | 1.05 (0.96-1.15) | 0.5681 |
| 0.5 | $5x10^{-4}$ | 1.03 (0.94-1.13) | 0.567 |
| 0.5 | $1x10^{-3}$ | 1.03 (0.94-1.13) | 0.5664 |
| 0.5 | $5x10^{-3}$ | 1.04 (0.95-1.14) | 0.5669 |
| 0.5 | $1x10^{-2}$ | 1.01 (0.93-1.11) | 0.5664 |
| 0.5 | $5x10^{-2}$ | 0.98 (0.89-1.07) | 0.5676 |
| 0.5 | 0.1 | 0.98 (0.89-1.07) | 0.5672 |
| 0.5 | 0.2 | 0.96 (0.87-1.05) | 0.5693 |
| 0.5 | 0.4 | 0.95 (0.87-1.05) | 0.5698 |
| 0.5 | 0.6 | 0.95 (0.87-1.04) | 0.57 |
| 0.5 | 0.8 | 0.95 (0.87-1.04) | 0.5701 |
| 0.5 | 1.0 | 0.95 (0.87-1.04) | 0.5701 |
| 0.1 | $1x10^{-8}$ | 1.00 (0.91-1.09) | 0.5666 |
| 0.1 | $5x10^{-8}$ | 1.01 (0.92-1.10) | 0.5665 |
| 0.1 | $1x10^{-7}$ | 1.01 (0.93-1.11) | 0.5664 |
| 0.1 | $5x10^{-7}$ | 1.04 (0.95-1.14) | 0.5668 |
| 0.1 | $1x10^{-6}$ | 1.03 (0.95-1.13) | 0.5664 |
| 0.1 | $5x10^{-6}$ | 1.01 (0.93-1.11) | 0.5665 |
| 0.1 | $1x10^{-5}$ | 1.03 (0.94-1.12) | 0.5664 |
| 0.1 | $5x10^{-5}$ | 1.05 (0.95-1.14) | 0.5679 |
| 0.1 | $1x10^{-4}$ | 1.07 (0.98-1.17) | 0.5699 |
| 0.1 | $5x10^{-4}$ | 1.03 (0.95-1.13) | 0.5663 |

| | | | |
|---|---|---|---|
| 0.1 | $1x10^{-3}$ | 1.02 (0.93-1.11) | 0.566 |
| 0.1 | $5x10^{-3}$ | 1.06 (0.97-1.16) | 0.5666 |
| 0.1 | $1x10^{-2}$ | 0.99 (0.91-1.09) | 0.5668 |
| 0.1 | $5x10^{-2}$ | 0.97 (0.88-1.06) | 0.5685 |
| 0.1 | 0.1 | 0.97 (0.88-1.06) | 0.5688 |
| 0.1 | 0.2 | 0.95 (0.87-1.04) | 0.5705 |
| 0.1 | 0.4 | 0.95 (0.87-1.04) | 0.5709 |
| 0.1 | 0.6 | 0.95 (0.86-1.04) | 0.571 |
| **0.1** | **0.8** | **0.94 (0.86-1.04)** | **0.571** |
| 0.1 | 1.0 | 0.94 (0.86-1.04) | 0.571 |

| **Supplementary table 8. Selection of UACR GRS** | | | |
|---|---|---|---|
| **$r^2$** | **P-value threshold** | **OR per SD** | **AUC** |
| 0.3 | $1x10^{-8}$ | 1.01 (0.92-1.11) | 0.5669 |
| 0.3 | $5x10^{-8}$ | 1.00 (0.92-1.10) | 0.5667 |
| 0.3 | $1x10^{-7}$ | 0.99 (0.90-1.08) | 0.5663 |
| 0.3 | $5x10^{-7}$ | 0.99 (0.90-1.08) | 0.5662 |
| 0.3 | $1x10^{-6}$ | 0.98 (0.90-1.08) | 0.5661 |
| 0.3 | $5x10^{-6}$ | 1.03 (0.94-1.13) | 0.568 |
| 0.3 | $1x10^{-5}$ | 1.03 (0.94-1.13) | 0.5681 |
| 0.3 | $5x10^{-5}$ | 1.00 (0.92-1.10) | 0.5666 |
| 0.3 | $1x10^{-4}$ | 1.02 (0.93-1.11) | 0.5669 |
| 0.3 | $5x10^{-4}$ | 0.97 (0.88-1.06) | 0.5669 |
| 0.3 | $1x10^{-3}$ | 0.94 (0.86-1.03) | 0.5686 |
| 0.3 | $5x10^{-3}$ | 0.93 (0.85-1.02) | 0.5677 |
| 0.3 | $1x10^{-2}$ | 0.92 (0.84-1.01) | 0.5701 |
| 0.3 | $5x10^{-2}$ | 0.90 (0.81-0.99) | 0.574 |
| 0.3 | 0.1 | 0.88 (0.80-0.98) | 0.5714 |
| 0.3 | 0.2 | 0.87 (0.78-0.96) | 0.5753 |
| 0.3 | 0.4 | 0.85 (0.77-0.95) | 0.5748 |
| 0.3 | 0.6 | 0.85 (0.76-0.94) | 0.5759 |
| 0.3 | 0.8 | 0.85 (0.76-0.94) | 0.5757 |
| 0.3 | 1.0 | 0.85 (0.76-0.94) | 0.5759 |
| 0.5 | $1x10^{-8}$ | 1.00 (0.91-1.10) | 0.5666 |
| 0.5 | $5x10^{-8}$ | 1.00 (0.91-1.10) | 0.5666 |
| 0.5 | $1x10^{-7}$ | 0.99 (0.91-1.09) | 0.5663 |
| 0.5 | $5x10^{-7}$ | 0.99 (0.90-1.08) | 0.5662 |
| 0.5 | $1x10^{-6}$ | 0.98 (0.90-1.08) | 0.5662 |
| 0.5 | $5x10^{-6}$ | 1.02 (0.94-1.12) | 0.5677 |
| 0.5 | $1x10^{-5}$ | 1.02 (0.93-1.12) | 0.5677 |
| 0.5 | $5x10^{-5}$ | 1.00 (0.91-1.10) | 0.5666 |
| 0.5 | $1x10^{-4}$ | 1.01 (0.92-1.10) | 0.5667 |
| 0.5 | $5x10^{-4}$ | 0.98 (0.89-1.07) | 0.5664 |
| 0.5 | $1x10^{-3}$ | 0.95 (0.87-1.04) | 0.5676 |
| 0.5 | $5x10^{-3}$ | 0.94 (0.85-1.03) | 0.5677 |
| 0.5 | $1x10^{-2}$ | 0.93 (0.85-1.02) | 0.5692 |
| 0.5 | $5x10^{-2}$ | 0.90 (0.82-0.99) | 0.5744 |
| 0.5 | 0.1 | 0.87 (0.79-0.96) | 0.5761 |
| 0.5 | 0.2 | 0.87 (0.78-0.96) | 0.5771 |
| 0.5 | 0.4 | 0.84 (0.76-0.94) | 0.5778 |
| 0.5 | 0.6 | 0.83 (0.75-0.92) | 0.581 |
| 0.5 | 0.8 | 0.83 (0.74-0.92) | 0.581 |
| **0.5** | **1.0** | **0.83 (0.74-0.92)** | **0.5812** |
| 0.1 | $1x10^{-8}$ | 1.02 (0.93-1.12) | 0.5672 |
| 0.1 | $5x10^{-8}$ | 1.01 (0.92-1.11) | 0.5668 |
| 0.1 | $1x10^{-7}$ | 1.01 (0.92-1.10) | 0.5667 |
| 0.1 | $5x10^{-7}$ | 1.01 (0.93-1.11) | 0.5671 |
| 0.1 | $1x10^{-6}$ | 0.99 (0.90-1.08) | 0.5662 |
| 0.1 | $5x10^{-6}$ | 1.05 (0.96-1.15) | 0.5702 |
| 0.1 | $1x10^{-5}$ | 1.05 (0.96-1.15) | 0.5702 |
| 0.1 | $5x10^{-5}$ | 0.99 (0.91-1.09) | 0.5666 |
| 0.1 | $1x10^{-4}$ | 1.01 (0.92-1.10) | 0.5666 |
| 0.1 | $5x10^{-4}$ | 0.96 (0.87-1.05) | 0.5678 |

| | | | |
|---|---|---|---|
| 0.1 | $1x10^{-3}$ | 0.94 (0.86-1.03) | 0.5694 |
| 0.1 | $5x10^{-3}$ | 0.94 (0.85-1.03) | 0.5666 |
| 0.1 | $1x10^{-2}$ | 0.92 (0.83-1.01) | 0.57 |
| 0.1 | $5x10^{-2}$ | 0.88 (0.80-0.97) | 0.574 |
| 0.1 | 0.1 | 0.93 (0.84-1.03) | 0.5654 |
| 0.1 | 0.2 | 0.93 (0.83-1.03) | 0.5661 |
| 0.1 | 0.4 | 0.95 (0.86-1.05) | 0.5653 |
| 0.1 | 0.6 | 0.95 (0.86-1.05) | 0.5655 |
| 0.1 | 0.8 | 0.96 (0.87-1.06) | 0.5651 |
| 0.1 | 1.0 | 0.95 (0.86-1.05) | 0.5652 |

| Supplementary table 9. Selection of TC GRS | | | |
|---|---|---|---|
| $r^2$ | **P-value threshold** | **OR per SD** | **AUC** |
| 0.3 | $1x10^{-8}$ | 1.00 (0.91-1.10) | 0.5664 |
| 0.3 | $5x10^{-8}$ | 0.99 (0.90-1.09) | 0.567 |
| 0.3 | $1x10^{-7}$ | 0.99 (0.90-1.09) | 0.567 |
| 0.3 | $5x10^{-7}$ | 1.00 (0.91-1.09) | 0.5668 |
| 0.3 | $1x10^{-6}$ | 1.00 (0.91-1.09) | 0.5668 |
| 0.3 | $5x10^{-6}$ | 1.00 (0.91-1.09) | 0.5668 |
| 0.3 | $1x10^{-5}$ | 1.00 (0.91-1.10) | 0.5665 |
| 0.3 | $5x10^{-5}$ | 0.99 (0.91-1.09) | 0.5669 |
| 0.3 | $1x10^{-4}$ | 0.99 (0.90-1.09) | 0.5671 |
| 0.3 | $5x10^{-4}$ | 1.00 (0.91-1.10) | 0.5667 |
| 0.3 | $1x10^{-3}$ | 0.99 (0.90-1.09) | 0.5671 |
| 0.3 | $5x10^{-3}$ | 0.97 (0.88-1.06) | 0.5687 |
| 0.3 | $1x10^{-2}$ | 0.98 (0.90-1.08) | 0.5675 |
| 0.3 | $5x10^{-2}$ | 0.98 (0.89-1.08) | 0.5676 |
| 0.3 | 0.1 | 0.97 (0.89-1.07) | 0.5682 |
| 0.3 | 0.2 | 0.97 (0.88-1.06) | 0.5685 |
| 0.3 | 0.4 | 0.96 (0.87-1.05) | 0.5699 |
| 0.3 | 0.6 | 0.95 (0.87-1.04) | 0.5703 |
| **0.3** | **0.8** | **0.95 (0.86-1.04)** | **0.5707** |
| 0.3 | 1.0 | 0.95 (0.87-1.04) | 0.5706 |
| 0.5 | $1x10^{-8}$ | 1.01 (0.92-1.11) | 0.5661 |
| 0.5 | $5x10^{-8}$ | 1.00 (0.91-1.10) | 0.5665 |
| 0.5 | $1x10^{-7}$ | 1.00 (0.91-1.10) | 0.5666 |
| 0.5 | $5x10^{-7}$ | 1.01 (0.92-1.11) | 0.5662 |
| 0.5 | $1x10^{-6}$ | 1.01 (0.92-1.11) | 0.5661 |
| 0.5 | $5x10^{-6}$ | 1.01 (0.93-1.11) | 0.5659 |
| 0.5 | $1x10^{-5}$ | 1.02 (0.93-1.12) | 0.5658 |
| 0.5 | $5x10^{-5}$ | 1.01 (0.92-1.10) | 0.5661 |
| 0.5 | $1x10^{-4}$ | 1.01 (0.92-1.10) | 0.5662 |
| 0.5 | $5x10^{-4}$ | 1.01 (0.92-1.11) | 0.566 |
| 0.5 | $1x10^{-3}$ | 1.01 (0.92-1.11) | 0.5661 |
| 0.5 | $5x10^{-3}$ | 0.99 (0.90-1.09) | 0.5673 |
| 0.5 | $1x10^{-2}$ | 1.00 (0.91-1.10) | 0.5667 |
| 0.5 | $5x10^{-2}$ | 0.99 (0.90-1.09) | 0.5668 |
| 0.5 | 0.1 | 0.99 (0.90-1.08) | 0.5673 |
| 0.5 | 0.2 | 0.98 (0.89-1.08) | 0.5676 |
| 0.5 | 0.4 | 0.97 (0.88-1.06) | 0.5689 |
| 0.5 | 0.6 | 0.96 (0.88-1.06) | 0.5691 |
| 0.5 | 0.8 | 0.96 (0.87-1.05) | 0.5696 |
| 0.5 | 1.0 | 0.96 (0.87-1.05) | 0.5696 |
| 0.1 | $1x10^{-8}$ | 1.02 (0.93-1.11) | 0.566 |
| 0.1 | $5x10^{-8}$ | 1.01 (0.92-1.11) | 0.5662 |
| 0.1 | $1x10^{-7}$ | 1.01 (0.92-1.11) | 0.5662 |
| 0.1 | $5x10^{-7}$ | 1.01 (0.92-1.10) | 0.5663 |
| 0.1 | $1x10^{-6}$ | 1.01 (0.92-1.11) | 0.5662 |
| 0.1 | $5x10^{-6}$ | 1.01 (0.93-1.11) | 0.5661 |
| 0.1 | $1x10^{-5}$ | 1.02 (0.93-1.12) | 0.5658 |
| 0.1 | $5x10^{-5}$ | 1.00 (0.91-1.09) | 0.5668 |
| 0.1 | $1x10^{-4}$ | 1.00 (0.91-1.10) | 0.5664 |
| 0.1 | $5x10^{-4}$ | 1.02 (0.93-1.12) | 0.5656 |

| | | | |
|---|---|---|---|
| 0.1 | $1x10^{-3}$ | 1.02 (0.93-1.11) | 0.5657 |
| 0.1 | $5x10^{-3}$ | 0.98 (0.89-1.08) | 0.5679 |
| 0.1 | $1x10^{-2}$ | 0.97 (0.89-1.07) | 0.5685 |
| 0.1 | $5x10^{-2}$ | 0.99 (0.90-1.09) | 0.567 |
| 0.1 | 0.1 | 0.99 (0.90-1.08) | 0.5671 |
| 0.1 | 0.2 | 0.97 (0.88-1.06) | 0.5684 |
| 0.1 | 0.4 | 0.96 (0.88-1.05) | 0.5692 |
| 0.1 | 0.6 | 0.96 (0.87-1.05) | 0.5696 |
| 0.1 | 0.8 | 0.96 (0.87-1.05) | 0.57 |
| 0.1 | 1.0 | 0.96 (0.87-1.05) | 0.5698 |

| Supplementary table 10. Selection of TG GRS | | | |
|---|---|---|---|
| $r^2$ | P-value threshold | OR per SD | AUC |
| 0.3 | 1x10$^{-8}$ | 0.99 (0.91-1.09) | 0.5666 |
| 0.3 | 5x10$^{-8}$ | 0.99 (0.91-1.09) | 0.5665 |
| 0.3 | 1x10$^{-7}$ | 0.99 (0.91-1.09) | 0.5666 |
| 0.3 | 5x10$^{-7}$ | 1.00 (0.91-1.09) | 0.5667 |
| 0.3 | 1x10$^{-6}$ | 1.00 (0.92-1.10) | 0.5665 |
| 0.3 | 5x10$^{-6}$ | 1.00 (0.91-1.10) | 0.5665 |
| 0.3 | 1x10$^{-5}$ | 1.00 (0.91-1.10) | 0.5666 |
| 0.3 | 5x10$^{-5}$ | 1.00 (0.92-1.10) | 0.5665 |
| 0.3 | 1x10$^{-4}$ | 1.01 (0.92-1.10) | 0.5666 |
| 0.3 | 5x10$^{-4}$ | 1.00 (0.92-1.10) | 0.5665 |
| 0.3 | 1x10$^{-3}$ | 1.00 (0.91-1.10) | 0.5666 |
| 0.3 | 5x10$^{-3}$ | 0.99 (0.90-1.08) | 0.5668 |
| 0.3 | 1x10$^{-2}$ | 0.98 (0.89-1.07) | 0.567 |
| 0.3 | 5x10$^{-2}$ | 0.94 (0.86-1.03) | 0.5701 |
| 0.3 | 0.1 | 0.95 (0.86-1.04) | 0.5702 |
| 0.3 | 0.2 | 0.94 (0.86-1.03) | 0.5706 |
| 0.3 | 0.4 | 0.93 (0.85-1.02) | 0.5717 |
| 0.3 | 0.6 | 0.93 (0.85-1.02) | 0.572 |
| 0.3 | 0.8 | 0.93 (0.85-1.02) | 0.5724 |
| 0.3 | 1.0 | 0.93 (0.85-1.02) | 0.5725 |
| 0.5 | 1x10$^{-8}$ | 1.03 (0.94-1.12) | 0.5666 |
| 0.5 | 5x10$^{-8}$ | 1.03 (0.94-1.13) | 0.5668 |
| 0.5 | 1x10$^{-7}$ | 1.03 (0.94-1.13) | 0.5669 |
| 0.5 | 5x10$^{-7}$ | 1.03 (0.94-1.13) | 0.5669 |
| 0.5 | 1x10$^{-6}$ | 1.04 (0.95-1.14) | 0.5673 |
| 0.5 | 5x10$^{-6}$ | 1.04 (0.95-1.14) | 0.5674 |
| 0.5 | 1x10$^{-5}$ | 1.04 (0.95-1.14) | 0.5672 |
| 0.5 | 5x10$^{-5}$ | 1.04 (0.95-1.14) | 0.5675 |
| 0.5 | 1x10$^{-4}$ | 1.04 (0.95-1.14) | 0.5674 |
| 0.5 | 5x10$^{-4}$ | 1.04 (0.95-1.13) | 0.567 |
| 0.5 | 1x10$^{-3}$ | 1.03 (0.94-1.13) | 0.5668 |
| 0.5 | 5x10$^{-3}$ | 1.02 (0.93-1.12) | 0.5664 |
| 0.5 | 1x10$^{-2}$ | 1.01 (0.92-1.10) | 0.5666 |
| 0.5 | 5x10$^{-2}$ | 0.98 (0.89-1.07) | 0.5675 |
| 0.5 | 0.1 | 0.96 (0.88-1.06) | 0.5687 |
| 0.5 | 0.2 | 0.95 (0.87-1.04) | 0.5697 |
| 0.5 | 0.4 | 0.93 (0.85-1.02) | 0.5715 |
| 0.5 | 0.6 | 0.93 (0.85-1.02) | 0.572 |
| 0.5 | 0.8 | 0.93 (0.85-1.02) | 0.5723 |
| 0.5 | 1.0 | 0.93 (0.85-1.01) | 0.5724 |
| 0.1 | 1x10$^{-8}$ | 1.06 (0.97-1.16) | 0.5682 |
| 0.1 | 5x10$^{-8}$ | 1.05 (0.96-1.16) | 0.5684 |
| 0.1 | 1x10$^{-7}$ | 1.06 (0.96-1.16) | 0.5685 |
| 0.1 | 5x10$^{-7}$ | 1.05 (0.96-1.16) | 0.5682 |
| 0.1 | 1x10$^{-6}$ | 1.05 (0.96-1.15) | 0.5681 |
| 0.1 | 5x10$^{-6}$ | 1.04 (0.95-1.14) | 0.5676 |
| 0.1 | 1x10$^{-5}$ | 1.03 (0.94-1.13) | 0.5671 |
| 0.1 | 5x10$^{-5}$ | 1.03 (0.94-1.13) | 0.5674 |
| 0.1 | 1x10$^{-4}$ | 1.03 (0.94-1.13) | 0.5674 |
| 0.1 | 5x10$^{-4}$ | 1.02 (0.93-1.12) | 0.5667 |

| | | | |
|---|---|---|---|
| 0.1 | $1x10^{-3}$ | 1.02 (0.93-1.12) | 0.5665 |
| 0.1 | $5x10^{-3}$ | 0.99 (0.91-1.09) | 0.5667 |
| 0.1 | $1x10^{-2}$ | 0.95 (0.87-1.05) | 0.5682 |
| 0.1 | $5x10^{-2}$ | 0.93 (0.85-1.02) | 0.5718 |
| 0.1 | 0.1 | 0.93 (0.85-1.02) | 0.5722 |
| 0.1 | 0.2 | 0.93 (0.85-1.02) | 0.5724 |
| 0.1 | 0.4 | 0.93 (0.85-1.02) | 0.5723 |
| 0.1 | 0.6 | 0.93 (0.85-1.02) | 0.5723 |
| 0.1 | 0.8 | 0.93 (0.85-1.02) | 0.5726 |
| **0.1** | **1.0** | **0.93 (0.85-1.01)** | **0.5726** |

| **Supplementary table 11. Selection of LDL GRS** | | | |
|---|---|---|---|
| **$r^2$** | **P-value threshold** | **OR per SD** | **AUC** |
| 0.3 | $1x10^{-8}$ | 0.95 (0.87-1.04) | 0.5698 |
| 0.3 | $5x10^{-8}$ | 0.95 (0.87-1.04) | 0.5698 |
| 0.3 | $1x10^{-7}$ | 0.96 (0.88-1.05) | 0.5691 |
| 0.3 | $5x10^{-7}$ | 0.95 (0.87-1.04) | 0.5698 |
| 0.3 | $1x10^{-6}$ | 0.95 (0.87-1.04) | 0.5698 |
| 0.3 | $5x10^{-6}$ | 0.94 (0.85-1.02) | 0.5718 |
| 0.3 | $1x10^{-5}$ | 0.94 (0.86-1.03) | 0.5714 |
| 0.3 | $5x10^{-5}$ | 0.95 (0.87-1.04) | 0.5704 |
| 0.3 | $1x10^{-4}$ | 0.95 (0.87-1.05) | 0.57 |
| 0.3 | $5x10^{-4}$ | 0.93 (0.85-1.02) | 0.5725 |
| 0.3 | $1x10^{-3}$ | 0.94 (0.85-1.03) | 0.5718 |
| 0.3 | $5x10^{-3}$ | 0.94 (0.85-1.03) | 0.5721 |
| **0.3** | **$1x10^{-2}$** | **0.93 (0.85-1.02)** | **0.5728** |
| 0.3 | $5x10^{-2}$ | 0.96 (0.87-1.05) | 0.57 |
| 0.3 | 0.1 | 0.96 (0.88-1.06) | 0.5691 |
| 0.3 | 0.2 | 0.96 (0.87-1.05) | 0.5695 |
| 0.3 | 0.4 | 0.96 (0.87-1.05) | 0.5694 |
| 0.3 | 0.6 | 0.96 (0.88-1.05) | 0.5693 |
| 0.3 | 0.8 | 0.96 (0.87-1.05) | 0.5696 |
| 0.3 | 1.0 | 0.96 (0.87-1.05) | 0.5695 |
| 0.5 | $1x10^{-8}$ | 0.97 (0.88-1.06) | 0.5686 |
| 0.5 | $5x10^{-8}$ | 0.96 (0.88-1.06) | 0.5689 |
| 0.5 | $1x10^{-7}$ | 0.97 (0.89-1.06) | 0.5686 |
| 0.5 | $5x10^{-7}$ | 0.97 (0.88-1.06) | 0.5687 |
| 0.5 | $1x10^{-6}$ | 0.96 (0.88-1.06) | 0.5687 |
| 0.5 | $5x10^{-6}$ | 0.95 (0.87-1.04) | 0.5707 |
| 0.5 | $1x10^{-5}$ | 0.95 (0.87-1.04) | 0.5704 |
| 0.5 | $5x10^{-5}$ | 0.96 (0.87-1.05) | 0.5697 |
| 0.5 | $1x10^{-4}$ | 0.96 (0.88-1.06) | 0.5692 |
| 0.5 | $5x10^{-4}$ | 0.94 (0.86-1.04) | 0.5713 |
| 0.5 | $1x10^{-3}$ | 0.95 (0.87-1.04) | 0.5709 |
| 0.5 | $5x10^{-3}$ | 0.94 (0.86-1.03) | 0.5715 |
| 0.5 | $1x10^{-2}$ | 0.93 (0.85-1.02) | 0.5728 |
| 0.5 | $5x10^{-2}$ | 0.94 (0.86-1.04) | 0.5713 |
| 0.5 | 0.1 | 0.96 (0.87-1.05) | 0.5696 |
| 0.5 | 0.2 | 0.95 (0.87-1.05) | 0.5701 |
| 0.5 | 0.4 | 0.96 (0.88-1.05) | 0.5692 |
| 0.5 | 0.6 | 0.96 (0.88-1.06) | 0.5691 |
| 0.5 | 0.8 | 0.96 (0.88-1.05) | 0.5693 |
| 0.5 | 1.0 | 0.96 (0.88-1.05) | 0.5694 |
| 0.1 | $1x10^{-8}$ | 0.98 (0.89-1.07) | 0.5675 |
| 0.1 | $5x10^{-8}$ | 0.98 (0.90-1.07) | 0.5674 |
| 0.1 | $1x10^{-7}$ | 0.98 (0.90-1.08) | 0.5672 |
| 0.1 | $5x10^{-7}$ | 0.98 (0.89-1.07) | 0.5675 |
| 0.1 | $1x10^{-6}$ | 0.97 (0.89-1.07) | 0.5678 |
| 0.1 | $5x10^{-6}$ | 0.95 (0.87-1.04) | 0.5706 |
| 0.1 | $1x10^{-5}$ | 0.96 (0.88-1.05) | 0.5698 |
| 0.1 | $5x10^{-5}$ | 0.97 (0.89-1.06) | 0.5684 |
| 0.1 | $1x10^{-4}$ | 0.98 (0.90-1.08) | 0.5675 |
| 0.1 | $5x10^{-4}$ | 0.95 (0.86-1.04) | 0.5714 |

| | | | |
|---|---|---|---|
| 0.1 | $1x10^{-3}$ | 0.96 (0.87-1.05) | 0.5701 |
| 0.1 | $5x10^{-3}$ | 0.94 (0.86-1.03) | 0.5724 |
| 0.1 | $1x10^{-2}$ | 0.93 (0.85-1.02) | 0.5728 |
| 0.1 | $5x10^{-2}$ | 0.96 (0.88-1.06) | 0.5692 |
| 0.1 | 0.1 | 0.96 (0.88-1.06) | 0.5689 |
| 0.1 | 0.2 | 0.96 (0.88-1.06) | 0.5691 |
| 0.1 | 0.4 | 0.96 (0.88-1.05) | 0.5691 |
| 0.1 | 0.6 | 0.96 (0.88-1.06) | 0.569 |
| 0.1 | 0.8 | 0.96 (0.88-1.05) | 0.5693 |
| 0.1 | 1.0 | 0.96 (0.88-1.05) | 0.5692 |

| **Supplementary table 12. Selection of HDL GRS** | | | |
|---|---|---|---|
| $r^2$ | **P-value threshold** | **OR per SD** | **AUC** |
| 0.3 | $1x10^{-8}$ | 1.12 (1.02-1.23) | 0.573 |
| 0.3 | $5x10^{-8}$ | 1.13 (1.03-1.24) | 0.5742 |
| 0.3 | $1x10^{-7}$ | 1.12 (1.03-1.23) | 0.5732 |
| 0.3 | $5x10^{-7}$ | 1.12 (1.03-1.23) | 0.5736 |
| 0.3 | $1x10^{-6}$ | 1.12 (1.03-1.23) | 0.5735 |
| 0.3 | $5x10^{-6}$ | 1.13 (1.03-1.23) | 0.5738 |
| 0.3 | $1x10^{-5}$ | 1.13 (1.03-1.24) | 0.5742 |
| 0.3 | $5x10^{-5}$ | 1.13 (1.03-1.24) | 0.5742 |
| 0.3 | $1x10^{-4}$ | 1.13 (1.03-1.23) | 0.5738 |
| 0.3 | $5x10^{-4}$ | 1.12 (1.02-1.22) | 0.5736 |
| 0.3 | $1x10^{-3}$ | 1.12 (1.02-1.23) | 0.5737 |
| 0.3 | $5x10^{-3}$ | 1.05 (0.96-1.15) | 0.5676 |
| 0.3 | $1x10^{-2}$ | 1.00 (0.91-1.09) | 0.5666 |
| 0.3 | $5x10^{-2}$ | 0.89 (0.81-0.97) | 0.578 |
| 0.3 | 0.1 | 0.88 (0.80-0.97) | 0.5795 |
| 0.3 | 0.2 | 0.90 (0.82-0.99) | 0.5761 |
| 0.3 | 0.4 | 0.90 (0.82-0.99) | 0.5762 |
| 0.3 | 0.6 | 0.90 (0.82-0.99) | 0.5764 |
| 0.3 | 0.8 | 0.90 (0.82-0.99) | 0.5764 |
| 0.3 | 1.0 | 0.90 (0.82-0.99) | 0.5764 |
| 0.5 | $1x10^{-8}$ | 1.12 (1.02-1.23) | 0.5732 |
| 0.5 | $5x10^{-8}$ | 1.13 (1.03-1.23) | 0.5739 |
| 0.5 | $1x10^{-7}$ | 1.12 (1.02-1.23) | 0.5732 |
| 0.5 | $5x10^{-7}$ | 1.13 (1.03-1.24) | 0.5743 |
| 0.5 | $1x10^{-6}$ | 1.13 (1.03-1.24) | 0.5745 |
| 0.5 | $5x10^{-6}$ | 1.13 (1.03-1.24) | 0.5738 |
| 0.5 | $1x10^{-5}$ | 1.13 (1.03-1.24) | 0.5744 |
| 0.5 | $5x10^{-5}$ | 1.13 (1.03-1.23) | 0.5742 |
| 0.5 | $1x10^{-4}$ | 1.12 (1.02-1.23) | 0.5734 |
| 0.5 | $5x10^{-4}$ | 1.12 (1.03-1.23) | 0.5746 |
| 0.5 | $1x10^{-3}$ | 1.13 (1.03-1.23) | 0.5646 |
| 0.5 | $5x10^{-3}$ | 1.07 (0.98-1.18) | 0.5693 |
| 0.5 | $1x10^{-2}$ | 1.03 (0.94-1.12) | 0.5663 |
| 0.5 | $5x10^{-2}$ | 0.90 (0.82-0.99) | 0.5751 |
| 0.5 | 0.1 | 0.89 (0.81-0.98) | 0.578 |
| 0.5 | 0.2 | 0.90 (0.82-0.99) | 0.5761 |
| 0.5 | 0.4 | 0.90 (0.82-0.99) | 0.5759 |
| 0.5 | 0.6 | 0.90 (0.82-0.99) | 0.5763 |
| 0.5 | 0.8 | 0.90 (0.82-0.99) | 0.5765 |
| 0.5 | 1.0 | 0.90 (0.82-0.99) | 0.5765 |
| 0.1 | $1x10^{-8}$ | 1.09 (0.99-1.19) | 0.5686 |
| 0.1 | $5x10^{-8}$ | 1.10 (1.00-1.20) | 0.5693 |
| 0.1 | $1x10^{-7}$ | 1.09 (1.00-1.20) | 0.5689 |
| 0.1 | $5x10^{-7}$ | 1.10 (1.00-1.20) | 0.5696 |
| 0.1 | $1x10^{-6}$ | 1.09 (1.00-1.20) | 0.5692 |
| 0.1 | $5x10^{-6}$ | 1.10 (1.00-1.20) | 0.57 |
| 0.1 | $1x10^{-5}$ | 1.11 (1.01-1.21) | 0.5706 |
| 0.1 | $5x10^{-5}$ | 1.11 (1.02-1.22) | 0.5714 |
| 0.1 | $1x10^{-4}$ | 1.12 (1.02-1.22) | 0.5714 |
| 0.1 | $5x10^{-4}$ | 1.09 (0.99-1.19) | 0.5699 |

| | | | |
|---|---|---|---|
| 0.1 | $1x10^{-3}$ | 1.09 (1.00-1.19) | 0.5701 |
| 0.1 | $5x10^{-3}$ | 0.98 (0.89-1.07) | 0.5673 |
| 0.1 | $1x10^{-2}$ | 0.92 (0.84-1.01) | 0.5723 |
| **0.1** | **$5x10^{-2}$** | **0.87 (0.79-0.95)** | **0.5828** |
| 0.1 | 0.1 | 0.88 (0.80-0.96) | 0.5804 |
| 0.1 | 0.2 | 0.90 (0.82-0.99) | 0.5766 |
| 0.1 | 0.4 | 0.91 (0.83-0.99) | 0.5761 |
| 0.1 | 0.6 | 0.91 (0.83-0.99) | 0.5761 |
| 0.1 | 0.8 | 0.91 (0.83-1.00) | 0.576 |
| 0.1 | 1.0 | 0.91 (0.83-0.99) | 0.5761 |

| Supplementary table 13. Selection of T2D GRS | | | |
|---|---|---|---|
| **$r^2$** | **P-value threshold** | **OR per SD** | **AUC** |
| 0.3 | $1x10^{-8}$ | 1.05 (0.96-1.15) | 0.5684 |
| 0.3 | $5x10^{-8}$ | 1.05 (0.96-1.15) | 0.5688 |
| 0.3 | $1x10^{-7}$ | 1.05 (0.96-1.15) | 0.5686 |
| 0.3 | $5x10^{-7}$ | 1.05 (0.96-1.15) | 0.5676 |
| 0.3 | $1x10^{-6}$ | 1.07 (0.98-1.17) | 0.5698 |
| 0.3 | $5x10^{-6}$ | 1.09 (1.00-1.20) | 0.571 |
| 0.3 | $1x10^{-5}$ | 1.11 (1.02-1.22) | 0.5755 |
| 0.3 | $5x10^{-5}$ | 1.12 (1.02-1.22) | 0.5773 |
| 0.3 | $1x10^{-4}$ | 1.12 (1.02-1.22) | 0.5773 |
| 0.3 | $5x10^{-4}$ | 1.10 (1.01-1.21) | 0.5763 |
| 0.3 | $1x10^{-3}$ | 1.10 (1.00-1.20) | 0.5753 |
| 0.3 | $5x10^{-3}$ | 1.09 (0.99-1.19) | 0.5746 |
| 0.3 | $1x10^{-2}$ | 1.09 (0.99-1.19) | 0.5741 |
| 0.3 | $5x10^{-2}$ | 1.08 (0.99-1.19) | 0.5738 |
| 0.3 | 0.1 | 1.08 (0.99-1.19) | 0.5737 |
| 0.3 | 0.2 | 1.08 (0.99-1.19) | 0.5736 |
| 0.3 | 0.4 | 1.08 (0.99-1.19) | 0.5736 |
| 0.3 | 0.6 | 1.08 (0.99-1.19) | 0.5736 |
| 0.3 | 0.8 | 1.08 (0.99-1.19) | 0.5736 |
| 0.3 | 1.0 | 1.08 (0.99-1.19) | 0.5736 |
| 0.5 | $1x10^{-8}$ | 1.02 (0.93-1.12) | 0.5669 |
| 0.5 | $5x10^{-8}$ | 1.02 (0.93-1.12) | 0.5669 |
| 0.5 | $1x10^{-7}$ | 1.02 (0.93-1.12) | 0.5669 |
| 0.5 | $5x10^{-7}$ | 1.02 (0.93-1.11) | 0.5667 |
| 0.5 | $1x10^{-6}$ | 1.04 (0.95-1.13) | 0.5673 |
| 0.5 | $5x10^{-6}$ | 1.05 (0.96-1.15) | 0.5677 |
| 0.5 | $1x10^{-5}$ | 1.06 (0.97-1.17) | 0.5699 |
| 0.5 | $5x10^{-5}$ | 1.09 (0.99-1.19) | 0.5726 |
| 0.5 | $1x10^{-4}$ | 1.10 (1.00-1.20) | 0.5745 |
| 0.5 | $5x10^{-4}$ | 1.10 (1.01-1.21) | 0.576 |
| 0.5 | $1x10^{-3}$ | 1.09 (1.00-1.20) | 0.5751 |
| 0.5 | $5x10^{-3}$ | 1.09 (1.00-1.20) | 0.5746 |
| 0.5 | $1x10^{-2}$ | 1.09 (0.99-1.19) | 0.5742 |
| 0.5 | $5x10^{-2}$ | 1.08 (0.99-1.19) | 0.5739 |
| 0.5 | 0.1 | 1.08 (0.99-1.19) | 0.5737 |
| 0.5 | 0.2 | 1.08 (0.99-1.19) | 0.5736 |
| 0.5 | 0.4 | 1.08 (0.99-1.19) | 0.5735 |
| 0.5 | 0.6 | 1.08 (0.99-1.19) | 0.5735 |
| 0.5 | 0.8 | 1.08 (0.99-1.19) | 0.5735 |
| 0.5 | 1.0 | 1.08 (0.99-1.19) | 0.5735 |
| 0.1 | $1x10^{-8}$ | 1.08 (0.99-1.19) | 0.5719 |
| 0.1 | $5x10^{-8}$ | 1.09 (1.00-1.20) | 0.5737 |
| 0.1 | $1x10^{-7}$ | 1.09 (1.00-1.19) | 0.573 |
| 0.1 | $5x10^{-7}$ | 1.10 (1.00-1.20) | 0.5723 |
| 0.1 | $1x10^{-6}$ | 1.11 (1.01-1.22) | 0.5753 |
| 0.1 | $5x10^{-6}$ | 1.14 (1.04-1.24) | 0.5764 |
| 0.1 | $1x10^{-5}$ | 1.13 (1.03-1.24) | 0.5781 |
| **0.1** | **$5x10^{-5}$** | **1.13 (1.03-1.23)** | **0.5783** |
| 0.1 | $1x10^{-4}$ | 1.12 (1.02-1.22) | 0.5775 |
| 0.1 | $5x10^{-4}$ | 1.10 (1.00-1.20) | 0.5751 |

| | | | |
|---|---|---|---|
| 0.1 | $1x10^{-3}$ | 1.09 (1.00-1.20) | 0.5746 |
| 0.1 | $5x10^{-3}$ | 1.09 (0.99-1.19) | 0.5742 |
| 0.1 | $1x10^{-2}$ | 1.08 (0.99-1.19) | 0.5738 |
| 0.1 | $5x10^{-2}$ | 1.08 (0.99-1.18) | 0.5733 |
| 0.1 | 0.1 | 1.08 (0.99-1.18) | 0.5733 |
| 0.1 | 0.2 | 1.08 (0.99-1.18) | 0.5734 |
| 0.1 | 0.4 | 1.08 (0.99-1.18) | 0.5734 |
| 0.1 | 0.6 | 1.08 (0.99-1.18) | 0.5734 |
| 0.1 | 0.8 | 1.08 (0.99-1.18) | 0.5734 |
| 0.1 | 1.0 | 1.08 (0.99-1.18) | 0.5734 |

| Supplementary table 14. Selection of HbA1c GRS | | | |
|---|---|---|---|
| **$r^2$** | **P-value threshold** | **OR per SD** | **AUC** |
| 0.3 | $1x10^{-8}$ | 1.04 (0.95-1.14) | 0.5664 |
| 0.3 | $5x10^{-8}$ | 1.06 (0.97-1.16) | 0.5673 |
| 0.3 | $1x10^{-7}$ | 1.06 (0.97-1.16) | 0.5674 |
| 0.3 | $5x10^{-7}$ | 1.04 (0.95-1.14) | 0.5666 |
| 0.3 | $1x10^{-6}$ | 1.05 (0.96-1.15) | 0.5671 |
| 0.3 | $5x10^{-6}$ | 1.03 (0.94-1.13) | 0.5663 |
| 0.3 | $1x10^{-5}$ | 1.03 (0.94-1.12) | 0.5663 |
| 0.3 | $5x10^{-5}$ | 1.00 (0.92-1.10) | 0.5665 |
| 0.3 | $1x10^{-4}$ | 1.00 (0.92-1.10) | 0.5665 |
| 0.3 | $5x10^{-4}$ | 1.05 (0.96-1.15) | 0.5673 |
| 0.3 | $1x10^{-3}$ | 1.07 (0.97-1.18) | 0.5685 |
| 0.3 | $5x10^{-3}$ | 1.12 (1.02-1.24) | 0.5741 |
| 0.3 | $1x10^{-2}$ | 1.13 (1.03-1.25) | 0.5757 |
| 0.3 | $5x10^{-2}$ | 1.25 (1.12-1.40) | 0.586 |
| 0.3 | 0.1 | 1.31 (1.16-1.47) | 0.5918 |
| 0.3 | 0.2 | 1.36 (1.21-1.54) | 0.5981 |
| 0.3 | 0.4 | 1.35 (1.19-1.52) | 0.5944 |
| 0.3 | 0.6 | 1.35 (1.19-1.53) | 0.5943 |
| 0.3 | 0.8 | 1.35 (1.19-1.53) | 0.5937 |
| 0.3 | 1.0 | 1.35 (1.19-1.53) | 0.5933 |
| 0.5 | $1x10^{-8}$ | 1.06 (0.97-1.16) | 0.5674 |
| 0.5 | $5x10^{-8}$ | 1.07 (0.98-1.18) | 0.5682 |
| 0.5 | $1x10^{-7}$ | 1.08 (0.99-1.18) | 0.5686 |
| 0.5 | $5x10^{-7}$ | 1.06 (0.97-1.16) | 0.5672 |
| 0.5 | $1x10^{-6}$ | 1.07 (0.97-1.17) | 0.5676 |
| 0.5 | $5x10^{-6}$ | 1.05 (0.96-1.15) | 0.5669 |
| 0.5 | $1x10^{-5}$ | 1.04 (0.94-1.14) | 0.5666 |
| 0.5 | $5x10^{-5}$ | 1.03 (0.94-1.13) | 0.5663 |
| 0.5 | $1x10^{-4}$ | 1.03 (0.94-1.13) | 0.5662 |
| 0.5 | $5x10^{-4}$ | 1.05 (0.96-1.16) | 0.5674 |
| 0.5 | $1x10^{-3}$ | 1.06 (0.97-1.17) | 0.5682 |
| 0.5 | $5x10^{-3}$ | 1.11 (1.01-1.22) | 0.573 |
| 0.5 | $1x10^{-2}$ | 1.13 (1.02-1.25) | 0.5749 |
| 0.5 | $5x10^{-2}$ | 1.24 (1.11-1.39) | 0.5856 |
| 0.5 | 0.1 | 1.29 (1.16-1.45) | 0.5912 |
| **0.5** | **0.2** | **1.37 (1.22-1.54)** | **0.5984** |
| 0.5 | 0.4 | 1.35 (1.20-1.52) | 0.5952 |
| 0.5 | 0.6 | 1.35 (1.19-1.52) | 0.5949 |
| 0.5 | 0.8 | 1.35 (1.20-1.53) | 0.5945 |
| 0.5 | 1.0 | 1.35 (1.19-1.52) | 0.5942 |
| 0.1 | $1x10^{-8}$ | 1.04 (0.95-1.14) | 0.5666 |
| 0.1 | $5x10^{-8}$ | 1.06 (0.97-1.16) | 0.5676 |
| 0.1 | $1x10^{-7}$ | 1.06 (0.97-1.16) | 0.5675 |
| 0.1 | $5x10^{-7}$ | 1.05 (0.96-1.15) | 0.567 |
| 0.1 | $1x10^{-6}$ | 1.06 (0.97-1.17) | 0.5678 |
| 0.1 | $5x10^{-6}$ | 1.05 (0.96-1.15) | 0.5669 |
| 0.1 | $1x10^{-5}$ | 1.05 (0.96-1.15) | 0.567 |
| 0.1 | $5x10^{-5}$ | 1.04 (0.95-1.14) | 0.5665 |
| 0.1 | $1x10^{-4}$ | 1.05 (0.96-1.15) | 0.567 |
| 0.1 | $5x10^{-4}$ | 1.09 (1.00-1.20) | 0.5713 |

| | | | |
|---|---|---|---|
| 0.1 | $1x10^{-3}$ | 1.10 (1.00-1.22) | 0.5717 |
| 0.1 | $5x10^{-3}$ | 1.15 (1.04-1.27) | 0.5778 |
| 0.1 | $1x10^{-2}$ | 1.15 (1.04-1.27) | 0.5778 |
| 0.1 | $5x10^{-2}$ | 1.28 (1.15-1.44) | 0.5896 |
| 0.1 | 0.1 | 1.32 (1.17-1.48 | 0.5931 |
| 0.1 | 0.2 | 1.37 (1.22-1.55) | 0.597 |
| 0.1 | 0.4 | 1.34 (1.19-1.51) | 0.5923 |
| 0.1 | 0.6 | 1.34 (1.19-1.52) | 0.5926 |
| 0.1 | 0.8 | 1.33 (1.18-1.51) | 0.5908 |
| 0.1 | 1.0 | 1.33 (1.17-1.50) | 0.5902 |

| Supplementary table 15. Selection of BMI GRS | | | |
|---|---|---|---|
| $r^2$ | P-value threshold | OR per SD | AUC |
| 0.3 | $1x10^{-8}$ | 1.10 (1.00-1.20) | 0.5682 |
| 0.3 | $5x10^{-8}$ | 1.10 (1.01-1.21) | 0.5683 |
| 0.3 | $1x10^{-7}$ | 1.11 (1.01-1.21) | 0.5687 |
| 0.3 | $5x10^{-7}$ | 1.11 (1.01-1.21) | 0.5694 |
| 0.3 | $1x10^{-6}$ | 1.11 (1.01-1.21) | 0.5686 |
| 0.3 | $5x10^{-6}$ | 1.11 (1.01-1.21) | 0.5677 |
| 0.3 | $1x10^{-5}$ | 1.10 (1.00-1.20) | 0.5663 |
| 0.3 | $5x10^{-5}$ | 1.10 (1.01-1.21) | 0.5665 |
| 0.3 | $1x10^{-4}$ | 1.13 (1.03-1.24) | 0.5698 |
| 0.3 | $5x10^{-4}$ | 1.21 (1.10-1.33) | 0.5852 |
| 0.3 | $1x10^{-3}$ | 1.22 (1.11-1.34) | 0.5876 |
| **0.3** | **$5x10^{-3}$** | **1.23 (1.11-1.35)** | **0.5893** |
| 0.3 | $1x10^{-2}$ | 1.22 (1.11-1.35) | 0.589 |
| 0.3 | $5x10^{-2}$ | 1.18 (1.07-1.30) | 0.584 |
| 0.3 | 0.1 | 1.15 (1.05-1.26) | 0.5808 |
| 0.3 | 0.2 | 1.13 (1.03-1.24) | 0.5792 |
| 0.3 | 0.4 | 1.13 (1.03-1.24) | 0.5793 |
| 0.3 | 0.6 | 1.13 (1.03-1.24) | 0.5793 |
| 0.3 | 0.8 | 1.13 (1.03-1.24) | 0.5792 |
| 0.3 | 1.0 | 1.13 (1.03-1.24) | 0.5792 |
| 0.5 | $1x10^{-8}$ | 1.09 (1.00-1.20) | 0.5691 |
| 0.5 | $5x10^{-8}$ | 1.09 (0.99-1.19) | 0.5684 |
| 0.5 | $1x10^{-7}$ | 1.09 (0.99-1.19) | 0.5687 |
| 0.5 | $5x10^{-7}$ | 1.09 (1.00-1.19) | 0.5686 |
| 0.5 | $1x10^{-6}$ | 1.09 (0.99-1.19) | 0.5679 |
| 0.5 | $5x10^{-6}$ | 1.10 (1.00-1.20) | 0.568 |
| 0.5 | $1x10^{-5}$ | 1.09 (1.00-1.20) | 0.5674 |
| 0.5 | $5x10^{-5}$ | 1.09 (1.00-1.20) | 0.5668 |
| 0.5 | $1x10^{-4}$ | 1.11 (1.01-1.22) | 0.5684 |
| 0.5 | $5x10^{-4}$ | 1.16 (1.06-1.27) | 0.577 |
| 0.5 | $1x10^{-3}$ | 1.18 (1.07-1.29) | 0.5801 |
| 0.5 | $5x10^{-3}$ | 1.21 (1.10-1.33) | 0.5853 |
| 0.5 | $1x10^{-2}$ | 1.21 (1.10-1.33) | 0.5866 |
| 0.5 | $5x10^{-2}$ | 1.17 (1.06-1.28) | 0.5829 |
| 0.5 | 0.1 | 1.14 (1.04-1.26) | 0.5806 |
| 0.5 | 0.2 | 1.13 (1.03-1.24) | 0.5788 |
| 0.5 | 0.4 | 1.12 (1.02-1.23) | 0.5781 |
| 0.5 | 0.6 | 1.12 (1.02-1.23) | 0.578 |
| 0.5 | 0.8 | 1.12 (1.02-1.23) | 0.578 |
| 0.5 | 1.0 | 1.12 (1.02-1.23) | 0.578 |
| 0.1 | $1x10^{-8}$ | 1.12 (1.02-1.23) | 0.569 |
| 0.1 | $5x10^{-8}$ | 1.11 (1.01-1.21) | 0.5674 |
| 0.1 | $1x10^{-7}$ | 1.12 (1.02-1.23) | 0.5688 |
| 0.1 | $5x10^{-7}$ | 1.10 (1.01-1.21) | 0.567 |
| 0.1 | $1x10^{-6}$ | 1.11 (1.01-1.21) | 0.5674 |
| 0.1 | $5x10^{-6}$ | 1.10 (1.00-1.20) | 0.5664 |
| 0.1 | $1x10^{-5}$ | 1.08 (0.99-1.18) | 0.565 |
| 0.1 | $5x10^{-5}$ | 1.09 (1.00-1.20) | 0.5654 |
| 0.1 | $1x10^{-4}$ | 1.12 (1.02-1.23) | 0.5682 |
| 0.1 | $5x10^{-4}$ | 1.22 (1.11-1.34) | 0.5882 |

| | | | |
|---|---|---|---|
| 0.1 | $1x10^{-3}$ | 1.21 (1.10-1.33) | 0.5876 |
| 0.1 | $5x10^{-3}$ | 1.22 (1.11-1.35) | 0.5893 |
| 0.1 | $1x10^{-2}$ | 1.22 (1.11-1.34) | 0.5889 |
| 0.1 | $5x10^{-2}$ | 1.16 (1.06-1.28) | 0.583 |
| 0.1 | 0.1 | 1.14 (1.04-1.25) | 0.5801 |
| 0.1 | 0.2 | 1.12 (1.02-1.23) | 0.5779 |
| 0.1 | 0.4 | 1.12 (1.02-1.23) | 0.578 |
| 0.1 | 0.6 | 1.12 (1.02-1.23) | 0.578 |
| 0.1 | 0.8 | 1.12 (1.02-1.23) | 0.5778 |
| 0.1 | 1.0 | 1.12 (1.02-1.23) | 0.5778 |

| Supplementary table 16. Selection of WHR GRS | | | |
|---|---|---|---|
| $r^2$ | P-value threshold | OR per SD | AUC |
| 0.3 | $1x10^{-8}$ | 1.09 (0.99-1.19) | 0.5713 |
| 0.3 | $5x10^{-8}$ | 1.10 (1.00-1.20) | 0.572 |
| 0.3 | $1x10^{-7}$ | 1.10 (1.00-1.20) | 0.5724 |
| 0.3 | $5x10^{-7}$ | 1.11 (1.02-1.22) | 0.5748 |
| 0.3 | $1x10^{-6}$ | 1.13 (1.03-1.23) | 0.5758 |
| 0.3 | $5x10^{-6}$ | 1.14 (1.04-1.25) | 0.578 |
| 0.3 | $1x10^{-5}$ | 1.14 (1.04-1.25) | 0.5786 |
| 0.3 | $5x10^{-5}$ | 1.16 (1.06-1.28) | 0.5816 |
| 0.3 | $1x10^{-4}$ | 1.17 (1.07-1.13) | 0.5832 |
| 0.3 | $5x10^{-4}$ | 1.17 (1.07-1.29) | 0.583 |
| 0.3 | $1x10^{-3}$ | 1.18 (1.08-1.29) | 0.5844 |
| 0.3 | $5x10^{-3}$ | 1.15 (1.05-1.26) | 0.5815 |
| 0.3 | $1x10^{-2}$ | 1.15 (1.05-1.27) | 0.5819 |
| 0.3 | $5x10^{-2}$ | 1.18 (1.07-1.29) | 0.5848 |
| 0.3 | 0.1 | 1.19 (1.08-1.30) | 0.5862 |
| 0.3 | 0.2 | 1.18 (1.07-1.29) | 0.5851 |
| 0.3 | 0.4 | 1.18 (1.08-1.30) | 0.5857 |
| 0.3 | 0.6 | 1.18 (1.07-1.30) | 0.5853 |
| 0.3 | 0.8 | 1.18 (1.08-1.30) | 0.5854 |
| 0.3 | 1.0 | 1.18 (1.08-1.30) | 0.5856 |
| 0.5 | $1x10^{-8}$ | 1.06 (0.97-1.16) | 0.5689 |
| 0.5 | $5x10^{-8}$ | 1.08 (0.99-1.18) | 0.5705 |
| 0.5 | $1x10^{-7}$ | 1.09 (0.99-1.19) | 0.5709 |
| 0.5 | $5x10^{-7}$ | 1.10 (1.10-1.20) | 0.5728 |
| 0.5 | $1x10^{-6}$ | 1.11 (1.01-1.21) | 0.5732 |
| 0.5 | $5x10^{-6}$ | 1.12 (1.02-1.22) | 0.5744 |
| 0.5 | $1x10^{-5}$ | 1.13 (1.03-1.23) | 0.5758 |
| 0.5 | $5x10^{-5}$ | 1.15 (1.05-1.26) | 0.5787 |
| 0.5 | $1x10^{-4}$ | 1.15 (1.05-1.26) | 0.5788 |
| 0.5 | $5x10^{-4}$ | 1.15 (1.05-1.26) | 0.5793 |
| 0.5 | $1x10^{-3}$ | 1.16 (1.06-1.27) | 0.5811 |
| 0.5 | $5x10^{-3}$ | 1.16 (1.05-1.27) | 0.582 |
| 0.5 | $1x10^{-2}$ | 1.17 (1.07-1.28) | 0.5837 |
| 0.5 | $5x10^{-2}$ | 1.19 (1.09-1.31) | 0.5859 |
| 0.5 | 0.1 | 1.19 (1.08-1.30) | 0.5863 |
| 0.5 | 0.2 | 1.18 (1.08-1.30) | 0.5851 |
| 0.5 | 0.4 | 1.19 (1.08-1.30) | 0.5858 |
| 0.5 | 0.6 | 1.18 (1.07-1.29) | 0.5847 |
| 0.5 | 0.8 | 1.18 (1.07-1.29) | 0.5849 |
| 0.5 | 1.0 | 1.18 (1.08-1.30) | 0.5851 |
| 0.1 | $1x10^{-8}$ | 1.12 (1.03-1.23) | 0.5756 |
| 0.1 | $5x10^{-8}$ | 1.13 (1.03-1.23) | 0.575 |
| 0.1 | $1x10^{-7}$ | 1.14 (1.04-1.25) | 0.5767 |
| 0.1 | $5x10^{-7}$ | 1.15 (1.05-1.26) | 0.579 |
| 0.1 | $1x10^{-6}$ | 1.16 (1.06-1.28) | 0.5807 |
| 0.1 | $5x10^{-6}$ | 1.16 (1.06-1.27) | 0.5808 |
| 0.1 | $1x10^{-5}$ | 1.15 (1.05-1.27) | 0.5805 |
| 0.1 | $5x10^{-5}$ | 1.19 (1.08-1.31) | 0.5846 |
| 0.1 | $1x10^{-4}$ | 1.20 (1.09-1.32) | 0.5862 |
| 0.1 | $5x10^{-4}$ | 1.18 (1.07-1.29) | 0.5832 |

| | | | |
|---|---|---|---|
| 0.1 | $1x10^{-3}$ | 1.17 (1.06-1.28) | 0.5826 |
| 0.1 | $5x10^{-3}$ | 1.13 (1.03-1.24) | 0.5791 |
| 0.1 | $1x10^{-2}$ | 1.12 (1.03-1.23) | 0.5788 |
| 0.1 | $5x10^{-2}$ | 1.16 (1.06-1.28) | 0.5828 |
| 0.1 | 0.1 | 1.19 (1.08-1.30) | 0.5863 |
| 0.1 | 0.2 | 1.19 (1.08-1.31) | 0.5864 |
| 0.1 | 0.4 | 1.18 (1.07-1.29) | 0.5849 |
| 0.1 | 0.6 | 1.19 (1.09-1.31) | 0.5864 |
| 0.1 | 0.8 | 1.19 (1.08-1.31) | 0.5862 |
| **0.1** | **1.0** | **1.19 (1.09-1.31)** | **0.5865** |

| **Supplementary table 17. Selection of SVS GRS** | | | |
|---|---|---|---|
| **$r^2$** | **P-value threshold** | **OR per SD** | **AUC** |
| 0.3 | $1x10^{-8}$ | - | - |
| 0.3 | $5x10^{-8}$ | - | - |
| 0.3 | $1x10^{-7}$ | 1.02 (0.93-1.12) | 0.5663 |
| 0.3 | $5x10^{-7}$ | 0.96 (0.88-1.05) | 0.5673 |
| 0.3 | $1x10^{-6}$ | 0.99 (0.90-1.08) | 0.5668 |
| 0.3 | $5x10^{-6}$ | 1.06 (0.96-1.16) | 0.5705 |
| 0.3 | $1x10^{-5}$ | 1.06 (0.97-1.16) | 0.5664 |
| 0.3 | $5x10^{-5}$ | 1.08 (0.99-1.19) | 0.568 |
| 0.3 | $1x10^{-4}$ | 1.06 (0.96-1.16) | 0.5676 |
| 0.3 | $5x10^{-4}$ | 1.11 (1.01-1.22) | 0.5731 |
| 0.3 | $1x10^{-3}$ | 1.04 (0.95-1.14) | 0.5653 |
| 0.3 | $5x10^{-3}$ | 1.02 (0.93-1.12) | 0.5656 |
| 0.3 | $1x10^{-2}$ | 1.02 (0.93-1.12) | 0.5657 |
| 0.3 | $5x10^{-2}$ | 1.01 (0.92-1.11) | 0.5663 |
| 0.3 | 0.1 | 1.02 (0.93-1.12) | 0.5658 |
| 0.3 | 0.2 | 1.01 (0.92-1.11) | 0.5663 |
| 0.3 | 0.4 | 0.99 (0.90-1.08) | 0.5671 |
| 0.3 | 0.6 | 0.98 (0.89-1.07) | 0.5676 |
| 0.3 | 0.8 | 0.97 (0.90-1.07) | 0.5677 |
| 0.3 | 1.0 | 0.98 (0.89-1.07) | 0.5676 |
| 0.5 | $1x10^{-8}$ | - | - |
| 0.5 | $5x10^{-8}$ | - | - |
| 0.5 | $1x10^{-7}$ | 1.02 (0.93-1.12) | 0.5663 |
| 0.5 | $5x10^{-7}$ | 0.94 (0.86-1.03) | 0.5677 |
| 0.5 | $1x10^{-6}$ | 0.97 (0.89-1.07) | 0.5675 |
| 0.5 | $5x10^{-6}$ | 1.05 (0.96-1.15) | 0.5704 |
| 0.5 | $1x10^{-5}$ | 1.07 (0.97-1.17) | 0.5672 |
| 0.5 | $5x10^{-5}$ | 1.10 (1.00-1.21) | 0.5698 |
| 0.5 | $1x10^{-4}$ | 1.07 (0.98-1.17) | 0.5683 |
| **0.5** | **$5x10^{-4}$** | **1.15 (1.05-1.26)** | **0.5767** |
| 0.5 | $1x10^{-3}$ | 1.06 (0.96-1.16) | 0.5654 |
| 0.5 | $5x10^{-3}$ | 1.04 (0.95-1.14) | 0.5656 |
| 0.5 | $1x10^{-2}$ | 1.03 (0.94-1.13) | 0.5656 |
| 0.5 | $5x10^{-2}$ | 1.03 (0.94-1.13) | 0.5657 |
| 0.5 | 0.1 | 1.05 (0.96-1.15) | 0.5663 |
| 0.5 | 0.2 | 1.04 (0.95-1.14) | 0.5659 |
| 0.5 | 0.4 | 1.02 (0.93-1.11) | 0.5659 |
| 0.5 | 0.6 | 1.01 (0.92-1.10) | 0.5663 |
| 0.5 | 0.8 | 1.00 (0.92-1.10) | 0.5665 |
| 0.5 | 1.0 | 1.01 (0.92-1.10) | 0.5664 |
| 0.1 | $1x10^{-8}$ | - | - |
| 0.1 | $5x10^{-8}$ | - | - |
| 0.1 | $1x10^{-7}$ | 1.02 (0.93-1.12) | 0.5663 |
| 0.1 | $5x10^{-7}$ | 0.96 (0.88-1.05) | 0.5673 |
| 0.1 | $1x10^{-6}$ | 0.99 (0.90-1.08) | 0.5668 |
| 0.1 | $5x10^{-6}$ | 1.06 (0.97-1.16) | 0.5711 |
| 0.1 | $1x10^{-5}$ | 1.07 (0.97-1.17) | 0.5665 |
| 0.1 | $5x10^{-5}$ | 1.09 (1.00-1.20) | 0.5686 |
| 0.1 | $1x10^{-4}$ | 1.07 (0.98-1.17) | 0.5685 |
| 0.1 | $5x10^{-4}$ | 1.12 (1.02-1.23) | 0.5749 |

| | | | |
|---|---|---|---|
| 0.1 | $1x10^{-3}$ | 1.01 (0.93-1.11) | 0.5659 |
| 0.1 | $5x10^{-3}$ | 1.01 (0.92-1.11) | 0.5661 |
| 0.1 | $1x10^{-2}$ | 1.01 (0.92-1.11) | 0.5661 |
| 0.1 | $5x10^{-2}$ | 0.99 (0.90-1.08) | 0.567 |
| 0.1 | 0.1 | 0.98 (0.90-1.08) | 0.5674 |
| 0.1 | 0.2 | 0.97 (0.89-1.07) | 0.568 |
| 0.1 | 0.4 | 0.96 (0.87-1.05) | 0.569 |
| 0.1 | 0.6 | 0.95 (0.87-1.04) | 0.5695 |
| 0.1 | 0.8 | 0.95 (0.97-1.04) | 0.5695 |
| 0.1 | 1.0 | 0.95 (0.87-1.04) | 0.5695 |

| Supplementary table 18. Selection of educational attainment GRS | | | |
|---|---|---|---|
| **$r^2$** | **P-value threshold** | **OR per SD** | **AUC** |
| 0.3 | $1x10^{-8}$ | 0.80 (0.73-0.88) | 0.5938 |
| 0.3 | $5x10^{-8}$ | 0.79 (0.72-0.86) | 0.5962 |
| 0.3 | $1x10^{-7}$ | 0.78 (0.71-0.85) | 0.5994 |
| 0.3 | $5x10^{-7}$ | 0.76 (0.69-0.83) | 0.6034 |
| 0.3 | $1x10^{-6}$ | 0.75 (0.68-0.83) | 0.605 |
| 0.3 | $5x10^{-6}$ | 0.75 (0.68-0.82) | 0.605 |
| 0.3 | $1x10^{-5}$ | 0.74 (0.67-0.81) | 0.607 |
| 0.3 | $5x10^{-5}$ | 0.72 (0.66-0.79) | 0.6131 |
| 0.3 | $1x10^{-4}$ | 0.74 (0.67-0.81) | 0.6088 |
| 0.3 | $5x10^{-4}$ | 0.75 (0.68-0.83) | 0.6018 |
| 0.3 | $1x10^{-3}$ | 0.77 (0.70-0.84) | 0.5974 |
| 0.3 | $5x10^{-3}$ | 0.70 (0.63-0.77) | 0.6178 |
| 0.3 | $1x10^{-2}$ | 0.69 (0.63-0.76) | 0.6178 |
| **0.3** | **$5x10^{-2}$** | **0.62 (0.56-0.69)** | **0.6432** |
| 0.3 | 0.1 | 0.65 (0.59-0.72) | 0.6361 |
| 0.3 | 0.2 | 0.68 (0.62-0.76) | 0.6263 |
| 0.3 | 0.4 | 0.79 (0.72-0.87) | 0.5996 |
| 0.3 | 0.6 | 0.78 (0.71-0.85) | 0.6027 |
| 0.3 | 0.8 | 0.78 (0.71-0.86) | 0.6022 |
| 0.3 | 1.0 | 0.78 (0.71-0.86) | 0.6014 |
| 0.5 | $1x10^{-8}$ | 0.83 (0.76-0.91) | 0.5877 |
| 0.5 | $5x10^{-8}$ | 0.81 (0.74-0.89) | 0.5907 |
| 0.5 | $1x10^{-7}$ | 0.80 (0.73-0.88) | 0.592 |
| 0.5 | $5x10^{-7}$ | 0.79 (0.72-0.86) | 0.5964 |
| 0.5 | $1x10^{-6}$ | 0.78 (0.71-0.86) | 0.5978 |
| 0.5 | $5x10^{-6}$ | 0.77 (0.70-0.84) | 0.6013 |
| 0.5 | $1x10^{-5}$ | 0.75 (0.68-0.83) | 0.6047 |
| 0.5 | $5x10^{-5}$ | 0.73 (0.67-0.81) | 0.6106 |
| 0.5 | $1x10^{-4}$ | 0.74 (0.67-0.81) | 0.6081 |
| 0.5 | $5x10^{-4}$ | 0.74 (0.67-0.81) | 0.6059 |
| 0.5 | $1x10^{-3}$ | 0.74 (0.67-0.81) | 0.6077 |
| 0.5 | $5x10^{-3}$ | 0.66 (0.60-0.73) | 0.6312 |
| 0.5 | $1x10^{-2}$ | 0.65 (0.59-0.72) | 0.6332 |
| 0.5 | $5x10^{-2}$ | 0.66 (0.59-0.72) | 0.6372 |
| 0.5 | 0.1 | 0.72 (0.65-0.79) | 0.6199 |
| 0.5 | 0.2 | 0.76 (0.69-0.83) | 0.6079 |
| 0.5 | 0.4 | 0.81 (0.73-0.89) | 0.5963 |
| 0.5 | 0.6 | 0.80 (0.73-0.88) | 0.5982 |
| 0.5 | 0.8 | 0.81 (0.73-0.89) | 0.5962 |
| 0.5 | 1.0 | 0.81 (0.74-0.89) | 0.5958 |
| 0.1 | $1x10^{-8}$ | 0.82 (0.74-0.89) | 0.5902 |
| 0.1 | $5x10^{-8}$ | 0.79 (0.72-0.87) | 0.5938 |
| 0.1 | $1x10^{-7}$ | 0.78 (0.71-0.85) | 0.5975 |
| 0.1 | $5x10^{-7}$ | 0.77 (0.70-0.84) | 0.5995 |
| 0.1 | $1x10^{-6}$ | 0.76 (0.69-0.83) | 0.6035 |
| 0.1 | $5x10^{-6}$ | 0.74 (0.67-0.81) | 0.6078 |
| 0.1 | $1x10^{-5}$ | 0.75 (0.68-0.82) | 0.604 |
| 0.1 | $5x10^{-5}$ | 0.75 (0.68-0.82) | 0.6043 |
| 0.1 | $1x10^{-4}$ | 0.78 (0.71-0.86) | 0.5942 |
| 0.1 | $5x10^{-4}$ | 0.82 (0.75-0.90) | 0.5818 |

| | | | |
|---|---|---|---|
| 0.1 | $1x10^{-3}$ | 0.83 (0.75-0.91) | 0.5812 |
| 0.1 | $5x10^{-3}$ | 0.77 (0.70-0.85) | 0.5931 |
| 0.1 | $1x10^{-2}$ | 0.80 (0.73-0.88) | 0.5856 |
| 0.1 | $5x10^{-2}$ | 0.64 (0.57-0.71) | 0.6323 |
| 0.1 | 0.1 | 0.64 (0.58-0.71) | 0.6367 |
| 0.1 | 0.2 | 0.71 (0.64-0.78) | 0.6192 |
| 0.1 | 0.4 | 0.76 (0.69-0.84) | 0.606 |
| 0.1 | 0.6 | 0.74 (0.68-0.82) | 0.6107 |
| 0.1 | 0.8 | 0.75 (0.68-0.82) | 0.6102 |
| 0.1 | 1.0 | 0.75 (0.68-0.83) | 0.6092 |

| Supplementary table 19. Selection of alcohol consumption GRS | | | |
|---|---|---|---|
| **$r^2$** | **P-value threshold** | **OR per SD** | **AUC** |
| 0.3 | $1x10^{-8}$ | 1.11 (1.01-1.22) | 0.5727 |
| 0.3 | $5x10^{-8}$ | 1.10 (1.00-1.21) | 0.5717 |
| 0.3 | $1x10^{-7}$ | 1.10 (1.00-1.21) | 0.5719 |
| 0.3 | $5x10^{-7}$ | 1.10 (1.00-1.21) | 0.5718 |
| 0.3 | $1x10^{-6}$ | 1.10 (1.00-1.21) | 0.5727 |
| 0.3 | $5x10^{-6}$ | 1.10 (1.00-1.21) | 0.5733 |
| 0.3 | $1x10^{-5}$ | 1.09 (1.00-1.20) | 0.5726 |
| 0.3 | $5x10^{-5}$ | 1.11 (1.01-1.21) | 0.5732 |
| 0.3 | $1x10^{-4}$ | 1.09 (0.99-1.19) | 0.5711 |
| 0.3 | $5x10^{-4}$ | 1.09 (0.99-1.19) | 0.5705 |
| 0.3 | $1x10^{-3}$ | 1.11 (1.02-1.22) | 0.5728 |
| 0.3 | $5x10^{-3}$ | 1.16 (1.06-1.28) | 0.5792 |
| 0.3 | $1x10^{-2}$ | 1.15 (1.04-1.26) | 0.5768 |
| 0.3 | $5x10^{-2}$ | 1.04 (0.95-1.14) | 0.5656 |
| 0.3 | 0.1 | 1.01 (0.93-1.11) | 0.566 |
| 0.3 | 0.2 | 0.96 (0.87-1.05) | 0.5694 |
| 0.3 | 0.4 | 0.94 (0.86-1.03) | 0.5704 |
| 0.3 | 0.6 | 0.94 (0.86-1.04) | 0.5703 |
| 0.3 | 0.8 | 0.94 (0.86-1.03) | 0.5704 |
| 0.3 | 1.0 | 0.94 (0.86-1.03) | 0.5705 |
| 0.5 | $1x10^{-8}$ | 1.10 (1.00-1.21) | 0.5719 |
| 0.5 | $5x10^{-8}$ | 1.09 (1.00-1.20) | 0.5712 |
| 0.5 | $1x10^{-7}$ | 1.10 (1.00-1.21) | 0.5722 |
| 0.5 | $5x10^{-7}$ | 1.10 (1.00-1.20) | 0.5719 |
| 0.5 | $1x10^{-6}$ | 1.10 (1.00-1.21) | 0.5723 |
| 0.5 | $5x10^{-6}$ | 1.11 (1.01-1.22) | 0.5739 |
| 0.5 | $1x10^{-5}$ | 1.11 (1.01-1.22) | 0.5737 |
| 0.5 | $5x10^{-5}$ | 1.12 (1.02-1.23) | 0.5754 |
| 0.5 | $1x10^{-4}$ | 1.10 (1.00-1.20) | 0.5725 |
| 0.5 | $5x10^{-4}$ | 1.09 (0.99-1.19) | 0.5704 |
| 0.5 | $1x10^{-3}$ | 1.10 (1.01-1.21) | 0.5713 |
| **0.5** | **$5x10^{-3}$** | **1.17 (1.07-1.29)** | **0.5806** |
| 0.5 | $1x10^{-2}$ | 1.14 (1.04-1.25) | 0.5751 |
| 0.5 | $5x10^{-2}$ | 1.06 (0.97-1.17) | 0.5664 |
| 0.5 | 0.1 | 1.04 (0.95-1.14) | 0.5657 |
| 0.5 | 0.2 | 0.97 (0.89-1.06) | 0.568 |
| 0.5 | 0.4 | 0.95 (0.87-1.04) | 0.5692 |
| 0.5 | 0.6 | 0.95 (0.87-1.04) | 0.5696 |
| 0.5 | 0.8 | 0.95 (0.86-1.04) | 0.5696 |
| 0.5 | 1.0 | 0.95 (0.86-1.04) | 0.5695 |
| 0.1 | $1x10^{-8}$ | 1.13 (1.03-1.24) | 0.5743 |
| 0.1 | $5x10^{-8}$ | 1.12 (1.02-1.23) | 0.5733 |
| 0.1 | $1x10^{-7}$ | 1.12 (1.02-1.22) | 0.573 |
| 0.1 | $5x10^{-7}$ | 1.12 (1.02-1.23) | 0.5743 |
| 0.1 | $1x10^{-6}$ | 1.12 (1.02-1.23) | 0.5743 |
| 0.1 | $5x10^{-6}$ | 1.12 (1.02-1.23) | 0.5752 |
| 0.1 | $1x10^{-5}$ | 1.11 (1.01-1.22) | 0.5749 |
| 0.1 | $5x10^{-5}$ | 1.13 (1.03-1.23) | 0.5758 |
| 0.1 | $1x10^{-4}$ | 1.10 (1.00-1.20) | 0.5754 |
| 0.1 | $5x10^{-4}$ | 1.10 (1.00-1.21) | 0.5714 |

| | | | |
|---|---|---|---|
| 0.1 | $1x10^{-3}$ | 1.11 (1.02-1.22) | 0.5721 |
| 0.1 | $5x10^{-3}$ | 1.15 (1.05-1.26) | 0.5773 |
| 0.1 | $1x10^{-2}$ | 1.12 (1.02-1.23) | 0.5732 |
| 0.1 | $5x10^{-2}$ | 1.00 (0.92-1.10) | 0.5664 |
| 0.1 | 0.1 | 0.97 (0.88-1.06) | 0.5682 |
| 0.1 | 0.2 | 0.94 (0.86-1.03) | 0.5712 |
| 0.1 | 0.4 | 0.93 (0.85-1.02) | 0.5722 |
| 0.1 | 0.6 | 0.93 (0.85-1.02) | 0.5725 |
| 0.1 | 0.8 | 0.93 (0.84-1.02) | 0.5727 |
| 0.1 | 1.0 | 0.93 (0.84-1.01) | 0.5728 |

| Supplementary table 20. Selection of smoking GRS | | | |
|---|---|---|---|
| **$r^2$** | **P-value threshold** | **OR per SD** | **AUC** |
| 0.3 | $1x10^{-8}$ | 1.04 (0.95-1.14) | 0.5666 |
| 0.3 | $5x10^{-8}$ | 1.05 (0.96-1.15) | 0.5674 |
| 0.3 | $1x10^{-7}$ | 1.06 (0.97-1.16) | 0.5678 |
| 0.3 | $5x10^{-7}$ | 1.09 (0.99-1.19) | 0.57 |
| 0.3 | $1x10^{-6}$ | 1.10 (1.01-1.21) | 0.5719 |
| 0.3 | $5x10^{-6}$ | 1.11 (1.01-1.21) | 0.5715 |
| 0.3 | $1x10^{-5}$ | 1.13 (1.03-1.24) | 0.5746 |
| 0.3 | $5x10^{-5}$ | 1.11 (1.01-1.22) | 0.5705 |
| 0.3 | $1x10^{-4}$ | 1.11 (1.01-1.22) | 0.5701 |
| 0.3 | $5x10^{-4}$ | 1.18 (1.08-1.30) | 0.5789 |
| 0.3 | $1x10^{-3}$ | 1.24 (1.13-1.36) | 0.5891 |
| 0.3 | $5x10^{-3}$ | 1.27 (1.15-1.39) | 0.599 |
| **0.3** | **$1x10^{-2}$** | **1.32 (1.20-1.45)** | **0.6063** |
| 0.3 | $5x10^{-2}$ | 1.23 (1.12-1.35) | 0.594 |
| 0.3 | 0.1 | 1.22 (1.11-1.34) | 0.5928 |
| 0.3 | 0.2 | 1.21 (1.10-1.33) | 0.5913 |
| 0.3 | 0.4 | 1.22 (1.11-1.34) | 0.593 |
| 0.3 | 0.6 | 1.22 (1.11-1.34) | 0.5925 |
| 0.3 | 0.8 | 1.22 (1.11-1.34) | 0.5923 |
| 0.3 | 1.0 | 1.22 (1.11-1.34) | 0.5924 |
| 0.5 | $1x10^{-8}$ | 1.05 (0.96-1.15) | 0.5669 |
| 0.5 | $5x10^{-8}$ | 1.06 (0.97-1.16) | 0.568 |
| 0.5 | $1x10^{-7}$ | 1.08 (0.98-1.18) | 0.5688 |
| 0.5 | $5x10^{-7}$ | 1.09 (0.99-1.19) | 0.5697 |
| 0.5 | $1x10^{-6}$ | 1.11 (1.01-1.22) | 0.5721 |
| 0.5 | $5x10^{-6}$ | 1.14 (1.04-1.25) | 0.5752 |
| 0.5 | $1x10^{-5}$ | 1.14 (1.04-1.25) | 0.5765 |
| 0.5 | $5x10^{-5}$ | 1.12 (1.02-1.23) | 0.5722 |
| 0.5 | $1x10^{-4}$ | 1.12 (1.02-1.23) | 0.5726 |
| 0.5 | $5x10^{-4}$ | 1.19 (1.08-1.30) | 0.5804 |
| 0.5 | $1x10^{-3}$ | 1.24 (1.13-1.36) | 0.5906 |
| 0.5 | $5x10^{-3}$ | 1.26 (1.14-1.38) | 0.5969 |
| 0.5 | $1x10^{-2}$ | 1.29 (1.17-1.42) | 0.6019 |
| 0.5 | $5x10^{-2}$ | 1.24 (1.13-1.37) | 0.596 |
| 0.5 | 0.1 | 1.23 (1.12-1.36) | 0.595 |
| 0.5 | 0.2 | 1.24 (1.13-1.37) | 0.5964 |
| 0.5 | 0.4 | 1.26 (1.14-1.39) | 0.5986 |
| 0.5 | 0.6 | 1.26 (1.14-1.38) | 0.5984 |
| 0.5 | 0.8 | 1.26 (1.14-1.38) | 0.5982 |
| 0.5 | 1.0 | 1.26 (1.14-1.39) | 0.5987 |
| 0.1 | $1x10^{-8}$ | 1.07 (0.98-1.17) | 0.5679 |
| 0.1 | $5x10^{-8}$ | 1.07 (0.98-1.18) | 0.569 |
| 0.1 | $1x10^{-7}$ | 1.08 (0.99-1.19) | 0.5696 |
| 0.1 | $5x10^{-7}$ | 1.12 (1.02-1.22) | 0.5732 |
| 0.1 | $1x10^{-6}$ | 1.14 (1.04-1.25) | 0.5764 |
| 0.1 | $5x10^{-6}$ | 1.15 (1.05-1.26) | 0.5762 |
| 0.1 | $1x10^{-5}$ | 1.15 (1.05-1.26) | 0.5772 |
| 0.1 | $5x10^{-5}$ | 1.10 (1.00-1.20) | 0.5697 |
| 0.1 | $1x10^{-4}$ | 1.10 (1.00-1.21) | 0.5696 |
| 0.1 | $5x10^{-4}$ | 1.20 (1.10-1.32) | 0.5817 |

| | | | |
|---|---|---|---|
| 0.1 | $1x10^{-3}$ | 1.27 (1.16-1.40) | 0.5963 |
| 0.1 | $5x10^{-3}$ | 1.24 (1.13-1.36) | 0.5953 |
| 0.1 | $1x10^{-2}$ | 1.31 (1.19-1.45) | 0.6058 |
| 0.1 | $5x10^{-2}$ | 1.22 (1.11-1.34) | 0.5931 |
| 0.1 | 0.1 | 1.20 (1.10-1.32) | 0.5907 |
| 0.1 | 0.2 | 1.19 (1.08-1.30) | 0.5881 |
| 0.1 | 0.4 | 1.20 (1.09-1.32) | 0.5899 |
| 0.1 | 0.6 | 1.20 (1.09-1.32) | 0.5897 |
| 0.1 | 0.8 | 1.19 (1.09-1.31) | 0.5887 |
| 0.1 | 1.0 | 1.19 (1.09-1.31) | 0.589 |

| **Supplementary table 21. Selection of insomnia GRS** | | | |
|---|---|---|---|
| **$r^2$** | **P-value threshold** | **OR per SD** | **AUC** |
| 0.3 | $1x10^{-8}$ | 1.08 (0.99-1.19) | 0.572 |
| 0.3 | $5x10^{-8}$ | 1.05 (0.96-1.14) | 0.569 |
| 0.3 | $1x10^{-7}$ | 1.05 (0.96-1.15) | 0.5696 |
| 0.3 | $5x10^{-7}$ | 1.05 (0.96-1.15) | 0.5699 |
| 0.3 | $1x10^{-6}$ | 1.05 (0.96-1.14) | 0.569 |
| 0.3 | $5x10^{-6}$ | 1.00 (0.92-1.10) | 0.5665 |
| 0.3 | $1x10^{-5}$ | 1.05 (0.96-1.15) | 0.5673 |
| 0.3 | $5x10^{-5}$ | 1.11 (1.01-1.21) | 0.5751 |
| 0.3 | $1x10^{-4}$ | 1.08 (0.99-1.19) | 0.572 |
| 0.3 | $5x10^{-4}$ | 1.09 (1.00-1.19) | 0.5744 |
| 0.3 | $1x10^{-3}$ | 1.12 (1.02-1.22) | 0.5775 |
| 0.3 | $5x10^{-3}$ | 1.08 (0.99-1.18) | 0.5718 |
| 0.3 | $1x10^{-2}$ | 1.03 (0.94-1.12) | 0.5664 |
| 0.3 | $5x10^{-2}$ | 1.08 (0.98-1.18) | 0.5728 |
| 0.3 | 0.1 | 1.06 (0.97-1.17) | 0.5711 |
| 0.3 | 0.2 | 1.05 (0.96-1.15) | 0.5686 |
| 0.3 | 0.4 | 0.98 (0.89-1.07) | 0.5674 |
| 0.3 | 0.6 | 0.96 (0.88-1.06) | 0.5683 |
| 0.3 | 0.8 | 0.95 (0.87-1.05) | 0.5692 |
| 0.3 | 1.0 | 0.95 (0.87-1.05) | 0.5691 |
| 0.5 | $1x10^{-8}$ | 1.05 (0.96-1.15) | 0.5683 |
| 0.5 | $5x10^{-8}$ | 1.03 (0.94-1.13) | 0.5675 |
| 0.5 | $1x10^{-7}$ | 1.04 (0.95-1.14) | 0.5679 |
| 0.5 | $5x10^{-7}$ | 1.05 (0.96-1.15) | 0.5693 |
| 0.5 | $1x10^{-6}$ | 1.06 (0.96-1.16) | 0.5696 |
| 0.5 | $5x10^{-6}$ | 1.03 (0.94-1.13) | 0.5665 |
| 0.5 | $1x10^{-5}$ | 1.07 (0.97-1.17) | 0.5687 |
| 0.5 | $5x10^{-5}$ | 1.11 (1.02-1.22) | 0.5761 |
| 0.5 | $1x10^{-4}$ | 1.11 (1.01-1.21) | 0.5753 |
| 0.5 | $5x10^{-4}$ | 1.09 (1.00-1.20) | 0.5748 |
| 0.5 | $1x10^{-3}$ | 1.12 (1.02-1.23) | 0.5776 |
| **0.5** | **$5x10^{-3}$** | **1.15 (1.05-1.26)** | **0.5811** |
| 0.5 | $1x10^{-2}$ | 1.12 (1.03-1.23) | 0.5766 |
| 0.5 | $5x10^{-2}$ | 1.08 (0.99-1.19) | 0.572 |
| 0.5 | 0.1 | 1.01 (0.92-1.10) | 0.5664 |
| 0.5 | 0.2 | 0.96 (0.88-1.06) | 0.5684 |
| 0.5 | 0.4 | 0.95 (0.87-1.04) | 0.57 |
| 0.5 | 0.6 | 0.95 (0.86-1.04) | 0.5708 |
| 0.5 | 0.8 | 0.94 (0.86-1.03) | 0.5709 |
| 0.5 | 1.0 | 0.94 (0.86-1.03) | 0.5709 |
| 0.1 | $1x10^{-8}$ | 1.07 (0.98-1.17) | 0.5698 |
| 0.1 | $5x10^{-8}$ | 1.01 (0.92-1.11) | 0.5669 |
| 0.1 | $1x10^{-7}$ | 1.01 (0.92-1.10) | 0.5668 |
| 0.1 | $5x10^{-7}$ | 1.02 (0.93-1.11) | 0.5671 |
| 0.1 | $1x10^{-6}$ | 0.99 (0.90-1.08) | 0.5663 |
| 0.1 | $5x10^{-6}$ | 0.98 (0.90-1.08) | 0.567 |
| 0.1 | $1x10^{-5}$ | 1.01 (0.93-1.11) | 0.5664 |
| 0.1 | $5x10^{-5}$ | 1.06 (0.97-1.16) | 0.5703 |
| 0.1 | $1x10^{-4}$ | 1.03 (0.94-1.13) | 0.568 |
| 0.1 | $5x10^{-4}$ | 1.08 (0.98-1.18) | 0.5728 |

| | | | |
|---|---|---|---|
| 0.1 | $1x10^{-3}$ | 1.10 (1.01-1.21) | 0.5753 |
| 0.1 | $5x10^{-3}$ | 1.02 (0.93-1.12) | 0.5669 |
| 0.1 | $1x10^{-2}$ | 1.02 (0.93-1.11) | 0.5668 |
| 0.1 | $5x10^{-2}$ | 1.06 (0.97-1.16) | 0.571 |
| 0.1 | 0.1 | 1.03 (0.94-1.14) | 0.5683 |
| 0.1 | 0.2 | 1.02 (0.93-1.12) | 0.5672 |
| 0.1 | 0.4 | 0.95 (0.87-1.04) | 0.5686 |
| 0.1 | 0.6 | 0.93 (0.85-1.02) | 0.5713 |
| 0.1 | 0.8 | 0.93 (0.85-1.02) | 0.5713 |
| 0.1 | 1.0 | 0.93 (0.85-1.02) | 0.5713 |

| Supplementary table 22. Selection of sleep duration GRS | | | |
|---|---|---|---|
| **$r^2$** | **P-value threshold** | **OR per SD** | **AUC** |
| 0.3 | $1x10^{-8}$ | 0.99 (0.90-1.08) | 0.5664 |
| 0.3 | $5x10^{-8}$ | 1.00 (0.91-1.09) | 0.5664 |
| 0.3 | $1x10^{-7}$ | 1.00 (0.91-1.09) | 0.5666 |
| 0.3 | $5x10^{-7}$ | 1.01 (0.92-1.11) | 0.5668 |
| 0.3 | $1x10^{-6}$ | 1.00 (0.91-1.09) | 0.5664 |
| 0.3 | $5x10^{-6}$ | 1.01 (0.92-1.10) | 0.5667 |
| 0.3 | $1x10^{-5}$ | 1.01 (0.92-1.10) | 0.5667 |
| 0.3 | $5x10^{-5}$ | 0.96 (0.88-1.05) | 0.568 |
| 0.3 | $1x10^{-4}$ | 0.94 (0.86-1.03) | 0.571 |
| 0.3 | $5x10^{-4}$ | 0.92 (0.84-1.01) | 0.5737 |
| 0.3 | $1x10^{-3}$ | 0.92 (0.84-1.01) | 0.5735 |
| 0.3 | $5x10^{-3}$ | 0.92 (0.84-1.01) | 0.5734 |
| 0.3 | $1x10^{-2}$ | 0.92 (0.84-1.01) | 0.5735 |
| 0.3 | $5x10^{-2}$ | 0.92 (0.84-1.01) | 0.5734 |
| 0.3 | 0.1 | 0.92 (0.84-1.01) | 0.5734 |
| 0.3 | 0.2 | 0.93 (0.84-1.01) | 0.5732 |
| 0.3 | 0.4 | 0.93 (0.84-1.01) | 0.5732 |
| 0.3 | 0.6 | 0.93 (0.85-1.01) | 0.5732 |
| 0.3 | 0.8 | 0.93 (0.85-1.01) | 0.5732 |
| 0.3 | 1.0 | 0.93 (0.85-1.01) | 0.5732 |
| 0.5 | $1x10^{-8}$ | 0.96 (0.88-1.05) | 0.5672 |
| 0.5 | $5x10^{-8}$ | 0.99 (0.90-1.08) | 0.5664 |
| 0.5 | $1x10^{-7}$ | 0.99 (0.91-1.09) | 0.5664 |
| 0.5 | $5x10^{-7}$ | 1.00 (0.91-1.09) | 0.5665 |
| 0.5 | $1x10^{-6}$ | 1.00 (0.92-1.10) | 0.5667 |
| 0.5 | $5x10^{-6}$ | 0.99 (0.91-1.09) | 0.5664 |
| 0.5 | $1x10^{-5}$ | 0.99 (0.91-1.09) | 0.5666 |
| 0.5 | $5x10^{-5}$ | 0.98 (0.90-1.07) | 0.567 |
| 0.5 | $1x10^{-4}$ | 0.96 (0.87-1.05) | 0.5688 |
| 0.5 | $5x10^{-4}$ | 0.93 (0.85-1.02) | 0.5719 |
| 0.5 | $1x10^{-3}$ | 0.93 (0.85-1.02) | 0.5711 |
| 0.5 | $5x10^{-3}$ | 0.92 (0.84-1.01) | 0.574 |
| **0.5** | **$1x10^{-2}$** | **0.92 (0.84-1.01)** | **0.5744** |
| 0.5 | $5x10^{-2}$ | 0.92 (0.84-1.01) | 0.5741 |
| 0.5 | 0.1 | 0.92 (0.84-1.01) | 0.5738 |
| 0.5 | 0.2 | 0.92 (0.84-1.01) | 0.5736 |
| 0.5 | 0.4 | 0.92 (0.84-1.01) | 0.5735 |
| 0.5 | 0.6 | 0.92 (0.84-1.01) | 0.5735 |
| 0.5 | 0.8 | 0.92 (0.84-1.01) | 0.5735 |
| 0.5 | 1.0 | 0.92 (0.84-1.01) | 0.5735 |
| 0.1 | $1x10^{-8}$ | 0.99 (0.90-1.08) | 0.5666 |
| 0.1 | $5x10^{-8}$ | 1.02 (0.93-1.12) | 0.5666 |
| 0.1 | $1x10^{-7}$ | 1.01 (0.92-1.11) | 0.5665 |
| 0.1 | $5x10^{-7}$ | 1.01 (0.92-1.11) | 0.5668 |
| 0.1 | $1x10^{-6}$ | 0.99 (0.90-1.08) | 0.5665 |
| 0.1 | $5x10^{-6}$ | 1.00 (0.91-1.09) | 0.5665 |
| 0.1 | $1x10^{-5}$ | 1.01 (0.92-1.11) | 0.5667 |
| 0.1 | $5x10^{-5}$ | 0.97 (0.89-1.07) | 0.5675 |
| 0.1 | $1x10^{-4}$ | 0.94 (0.86-1.03) | 0.5716 |
| 0.1 | $5x10^{-4}$ | 0.92 (0.84-1.01) | 0.5739 |

| | | | |
|---|---|---|---|
| 0.1 | $1x10^{-3}$ | 0.93 (0.85-1.02) | 0.573 |
| 0.1 | $5x10^{-3}$ | 0.93 (0.85-1.02) | 0.5728 |
| 0.1 | $1x10^{-2}$ | 0.93 (0.85-1.01) | 0.5732 |
| 0.1 | $5x10^{-2}$ | 0.93 (0.84-1.01) | 0.5732 |
| 0.1 | 0.1 | 0.93 (0.85-1.01) | 0.5731 |
| 0.1 | 0.2 | 0.93 (0.85-1.02) | 0.5729 |
| 0.1 | 0.4 | 0.93 (0.85-1.02) | 0.5728 |
| 0.1 | 0.6 | 0.93 (0.85-1.02) | 0.5728 |
| 0.1 | 0.8 | 0.93 (0.85-1.02) | 0.5728 |
| 0.1 | 1.0 | 0.93 (0.85-1.02) | 0.5728 |

**Supplementary Table 23. Characteristics of optimized genomic risk scores.**

| Trait | Tuning parameters | N variants in score (% availability in datasets) | OR per SD* (95% CI) | Beta* | AUCº |
|---|---|---|---|---|---|
| SBP | $r^2 < 0.1$, $p \leq 1x10^{-5}$ | 3,387 (99, 99, 99.1) | 1.16 (1.04-1.29) | 0.1460 | 0.5734 |
| DBP | $r^2 < 0.1$, $p \leq 5x10^{-5}$ | 5,065 (98.7, 98.7, 99.2) | 1.15 (0.90-1.47) | 0.1415 | 0.5752 |
| PP | $r^2 < 0.1$, $p \leq 0.8$ | 255,171 (94, 93.7, 96.7) | 1.04 (0.90-1.21) | 0.0405 | 0.5748 |
| WMH | $r^2 < 0.5$, $p \leq 5x10^{-2}$ | 87,951 (79.7, 79.5, 81.8) | 1.10 (1.00-1.20) | 0.094 | 0.5733 |
| CKD | $r^2 < 0.5$, $p \leq 5x10^{-6}$ | 213 (86.4, 85.4, 86.4) | 1.06 (0.95-1.19) | 0.0578 | 0.5731 |
| eGFR | $r^2 < 0.1$, $p \leq 0.8$ | 432,924 (68.4, 69.1, 69.7) | 1.06 (0.96-1.18) | 0.062 | 0.571 |
| UACR | $r^2 < 0.5$, $p \leq 1$ | 1,148,192 (81.7, 81.9, 83) | 1.01 (0.90-1.13) | 0.0058 | 0.5812 |
| TC | $r^2 < 0.3$, $p \leq 0.8$ | 225,988 (98, 97.6, 97.6) | 0.98 (0.81-1.19) | -0.0158 | 0.5707 |
| TG | $r^2 < 0.1$, $p \leq 1$ | 132,538 (97.2, 96.6, 96.7) | 1.02 (0.92-1.12) | 0.0154 | 0.5726 |
| LDL | $r^2 < 0.3$, $p \leq 1x10^{-2}$ | 6,242 (98.1, 98, 98.3) | 0.92 (0.84-1.02) | -0.0803 | 0.5728 |
| HDL | $r^2 < 0.1$, $p \leq 5x10^{-2}$ | 17,425 (98.1, 97.7, 97.8) | 0.96 (0.77-1.20) | -0.0363 | 0.5828 |
| T2D | $r^2 < 0.1$, $p \leq 5x10^{-5}$ | 2,204 (87.6, 87.7, 87.3) | 1.08 (0.98-1.18) | 0.0724 | 0.5783 |
| HbA1c | $r^2 < 0.5$, $p \leq 0.2$ | 106,654 (98.3, 98, 98) | 1.16 (1.02-1.31) | 0.1476 | 0.5984 |
| BMI | $r^2 < 0.3$, $p \leq 5x10^{-3}$ | 63,162 (77.3, 76.9, 77.7) | 1.12 (1.01-1.24) | 0.1134 | 0.5893 |
| WHR | $r^2 < 0.1$, $p \leq 1$ | 954,291 (33.1, 32.9, 33.4) | 1.15 (1.04-1.28) | 0.1415 | 0.5865 |
| SVS | $r^2 < 0.5$, $p \leq 5x10^{-4}$ | 2,162 (82.9, 83.7, 83.9) | 1.08 (0.98-1.19) | 0.0767 | 0.5767 |
| Educational attainment | $r^2 < 0.3$, $p \leq 5x10^{-2}$ | 147,505 (71.8, 71.5, 71.8) | 0.64 (0.40-1.04) | -0.4448 | 0.6432 |
| Alcohol consumption | $r^2 < 0.5$, $p \leq 5x10^{-3}$ | 14,627 (97, 96.2, 96.4) | 1.18 (1.07-1.31) | 0.1654 | 0.5806 |
| Smoking | $r^2 < 0.3$, $p \leq 1x10^{-2}$ | 34,347 (95, 94.7, 96.4) | 1.31 (0.90-1.89) | 0.2666 | 0.6063 |
| Insomnia | $r^2 < 0.5$, $p \leq 5x10^{-3}$ | 31,813 (60.6, 59.9, 59.9) | 1.12 (1.01-1.24) | 0.1116 | 0.5811 |
| Sleep duration | $r^2 < 0.5$, $p \leq 1x10^{-2}$ | 60,031 (57.4, 56.6, 56.7) | 0.96 (0.82-1.12) | -0.0433 | 0.5744 |

*ORs and betas are estimates after inverse-variance, random-effects meta-analysis across the training datasets. ºAUC values are derived from merged training datasets. Availability in datasets in the following order: GOCHA, EUR/ISGC, GERFHS.

SBP systolic blood pressure; DBP diastolic blood pressure; PP pulse pressure; WMH white matter hyperintensities; CKD chronic kidney disease; eGFR estimated glomerular filtration rate; UACR urine albumin-to-creatinine ratio; TC total cholesterol; TG triglycerides; LDL low-density lipoprotein; HDL high-density lipoprotein; T2D type 2 diabetes mellitus; HbA1c hemoglobin A1c; BMI body mass index; WHR waist-to-hip ratio; SVS small vessel stroke; GRS genomic risk score; AUC area under the receiving-operating characteristics curve

**Supplementary Table 24. Exploratory factor analysis of trait-specific GRS.**

| | Factor 1 | Factor 2 | Factor 3 | Factor 4 | Factor 5 | Uniqueness |
|---|---|---|---|---|---|---|
| **SBP** | 0.05 | 0.72 | -0.02 | 0.14 | 0.30 | 0.37 |
| **DBP** | 0.05 | 0.99 | 0.06 | 0.11 | -0.03 | 0.01 |
| **PP** | -0.19 | 0.24 | -0.24 | 0.17 | 0.90 | 0.01 |
| **WMH** | 0.22 | 0.08 | 0.01 | 0.23 | 0.01 | 0.89 |
| **T2D** | 0.39 | 0.06 | 0.06 | 0.05 | -0.03 | 0.84 |
| **HbA1c** | 0.56 | -0.07 | 0.13 | -0.12 | -0.14 | 0.63 |
| **TC** | -0.05 | -0.04 | 0.99 | -0.04 | -0.12 | 0.01 |
| **HDL** | -0.44 | 0.01 | 0.07 | 0.14 | 0.18 | 0.75 |
| **LDL** | 0.06 | -0.03 | 0.59 | 0.01 | -0.08 | 0.65 |
| **TG** | 0.01 | 0.11 | 0.29 | 0.01 | 0.20 | 0.87 |
| **BMI** | 0.72 | -0.07 | 0.00 | 0.11 | 0.00 | 0.46 |
| **WHR** | 0.56 | 0.02 | 0.01 | -0.09 | 0.06 | 0.67 |
| **UACR** | -0.51 | 0.11 | -0.04 | 0.30 | 0.19 | 0.60 |
| **CKD** | 0.06 | -0.06 | -0.01 | -0.08 | -0.06 | 0.98 |
| **eGFR** | -0.23 | 0.04 | -0.05 | 0.30 | 0.04 | 0.85 |
| **SVS** | 0.21 | 0.00 | 0.01 | -0.06 | -0.06 | 0.95 |
| **EA** | -0.61 | -0.06 | 0.00 | 0.18 | 0.00 | 0.60 |
| **Alcohol** | -0.03 | -0.01 | -0.09 | 0.04 | 0.07 | 0.99 |
| **Smoking** | 0.64 | 0.06 | -0.01 | 0.04 | 0.02 | 0.59 |
| **Insomnia** | 0.05 | 0.00 | 0.03 | -0.41 | -0.03 | 0.83 |
| **Sleep duration** | -0.05 | 0.12 | -0.02 | 0.75 | 0.08 | 0.42 |
| **Variance explained** | 0.13 | 0.077 | 0.072 | 0.053 | 0.051 | |
| **Cumulative variance explained** | 0.13 | 0.208 | 0.279 | 0.333 | 0.384 | |

**Supplementary Table 25. Weights and GRS included in the different versions of the metaGRS.**

| GRS | *All traits metaGRS | *Causal metaGRS | *Factor metaGRS | °Stepwise metaGRS | °Lasso metaGRS |
|---|---|---|---|---|---|
| | **OR (95% CI)** | **OR (95% CI)** | **OR (95% CI)** | **OR (95% CI)** | **Penalized OR** |
| **SBP** | 1.16 (1.04-1.29) | 1.16 (1.04-1.29) | . | 1.18 (1.06-1.31) | 1.10 |
| **DBP** | 1.15 (0.90-1.47) | 1.15 (0.90-1.47) | 1.15 (0.90-1.47) | . | 1.05 |
| **PP** | 1.04 (0.90-1.21) | . | 1.04 (0.90-1.21) | 0.78 (0.70-0.87) | 0.83 |
| **WMH** | 1.10 (1.00-1.20) | . | . | 1.08 (0.98-1.18) | 1.07 |
| **SVS** | 1.08 (0.98-1.19) | . | . | . | 1.05 |
| **TC** | 0.98 (0.81-1.19) | 0.98 (0.81-1.19) | 0.98 (0.81-1.19) | . | 1.06 |
| **TG** | 1.02 (0.92-1.12) | . | . | 0.93 (0.84-1.02) | 0.93 |
| **LDL** | 0.92 (0.84-1.02) | 0.92 (0.84-1.02) | . | 0.92 (0.84-1.01) | 0.90 |
| **HDL** | 0.96 (0.77-1.20) | . | . | . | 0.99 |
| **CKD** | 1.06 (0.95-1.19) | 1.06 (0.95-1.19) | . | . | 1.04 |
| **GFR** | 1.06 (0.96-1.18) | . | . | . | 1.06 |
| **UACR** | 1.01 (0.90-1.13) | 1.01 (0.90-1.13) | . | . | 1.03 |
| **T2D** | 1.08 (0.98-1.18) | . | . | . | 1.03 |
| **HbA1c** | 1.16 (1.02-1.31) | . | . | 1.09 (0.98-1.20) | 1.07 |
| **BMI** | 1.12 (1.01-1.24) | . | 1.12 (1.01-1.24) | . | . |
| **WHR** | 1.15 (1.04-1.28) | 1.15 (1.04-1.28) | . | . | 1.03 |
| **Education** | 0.64 (0.40-1.04) | 0.64 (0.40-1.04) | 0.64 (0.40-1.04) | 0.74 (0.67-0.82) | 0.76 |
| **EtOH** | 1.18 (1.07-1.31) | . | . | 1.20 (1.09-1.31) | 1.18 |
| **Smoking** | 1.31 (0.90-1.89) | . | . | . | 1.02 |
| **Insomnia** | 1.12 (1.01-1.24) | . | . | 1.09 (1.00-1.20) | 1.07 |
| **Sleep duration** | 0.96 (0.82-1.12) | . | 0.96 (0.82-1.12) | . | 0.96 |

*Estimates from the 'All-trait metaGRS', 'Causal metaGRS', and 'Factor metaGRS' are from inverse-variance random-effects meta-analysis across the training datasets. °Estimates from 'Stepwise metaGRS' and 'Lasso metaGRS' are from merged training datasets. Estimates for the 'All-trait metaGRS', 'Causal metaGRS' and 'Factor metaGRS' are adjusted for age, sex, and two PCs.

SBP systolic blood pressure; DBP diastolic blood pressure; PP pulse pressure; WMH white matter hyperintensities; CKD chronic kidney disease; eGFR estimated glomerular filtration rate; UACR urine albumin-to-creatinine ratio; TC total cholesterol; TG triglycerides; LDL low-density lipoprotein; HDL high-density lipoprotein; T2D type 2 diabetes mellitus; HbA1c hemoglobin A1c; BMI body mass index; WHR waist-to-hip ratio; SVS small vessel stroke; GRS genomic risk score

**Supplementary Table 26. Effect size estimates and predictive performance of different metaGRS versions**

| metaGRS version | OR (95% CI) | P-value | C-index (95% CI) |
|---|---|---|---|
| All-trait metaGRS | 1.45 (1.30-1.63) | $6.2x10^{-11}$ | 0.5936 (0.5723-0.6219) |
| Causal metaGRS | 1.36 (1.23-1.51) | $2.47x10^{-9}$ | 0.5850 (0.5637-0.6150) |
| Factor metaGRS | 1.40 (1.26-1.56) | $1.95x10^{-10}$ | 0.5887137 (0.5648-0.6209) |
| Stepwise metaGRS | 1.28 (1.14-1.44) | $2.14x10^{-5}$ | 0.561 (0.5415-0.5959) |
| Lasso metaGRS | 1.34 (1.19-1.50) | $5.55x10^{-7}$ | 0.5721 (0.5503-0.6043) |

All models include age, sex, and two PCs. The 95% CIs of the c-indices are calculated after bootstrapping over 1000 iterations.

**Supplementary Table 27. Significant clinical predictors of ICH in validation dataset after backward elimination**

| | **OR (95% CI)** | **p-value** |
|---|---|---|
| History of Ischemic stroke | 2.37 (1.40, 4.02) | 0.0014 |
| Hypertension | 1.65 (1.32, 2.07) | <0.0001 |
| Diabetes | 1.32 (1.00, 1.75) | 0.0500 |
| High Cholesterol | 0.64 (0.51, 0.79) | <0.0001 |
| Heavy alcohol | 1.89 (1.19, 2.99) | 0.0065 |
| Anticoagulant use | 2.65 (1.84, 3.80) | <0.0001 |
| Education: | | |
| Less than Equal High School | 2.25 (1.82, 2.79) | <0.0001 |
| Greater than High School | Reference | |

**Supplementary Table 28. Effect size estimates of all metaGRS versions after adjusting for clinical predictors and improvement in predictive ability.**

| metaGRS version | OR (95% CI) | P-value | $AIC_{clin}$ , $AIC_{clin+metaGRS}$, LRT |
|---|---|---|---|
| All-trait metaGRS | 1.31 (1.16-1.48) | < 0.0001 | 1989.65, 1972.86, 0.000041702 |
| Causal metaGRS | 1.24 (1.11-1.39) | 0.0002 | 1989.65, 1977.36, 0.000454161 |
| Factor metaGRS | 1.29 (1.15-1.44) | < 0.0001 | 1989.65, 1972.78, 0.000040045 |
| Stepwise metaGRS | 1.21 (1.07-1.37) | 0.0032 | 1989.65, 1982.87, 0.009226219 |
| Lasso metaGRS | 1.24 (1.09-1.40) | 0.0008 | 1989.65, 1980.18, 0.002094090 |

All effect size estimates are adjusted for age, sex, two principal components, history of ischemic stroke, hypertension, diabetes, high cholesterol, heavy alcohol use, anticoagulant use and less than high school education.

$AIC_{clin}$ = AIC of model containing baseline (age, sex, 2 PCs) and clinical predictors. $AIC_{clin+metaGRS}$ = AIC of model containing baseline, clinical predictors and each metaGRS version respectively.

AIC = Akaike information criterion, LRT = likelihood ratio test

**Supplementary Table 29. Lobar ICH characteristics in validation dataset.**

| | Controls N=796 | | Cases N=334 | | P-value |
|---|---|---|---|---|---|
| | **N Avail** | **N (%)** | **N Avail** | **N (%)** | |
| Age: Mean(SD) | 796 | 68.4 (13.4) | 334 | 71.0 (13.5) | 0.0033 |
| Female | 796 | 400 (50.2) | 334 | 180 (53.9) | 0.2638 |
| Race: | 796 | | 334 | | |
| White | | 796 (100.0) | | 334 (100.0) | |
| American Indian/Alaskan Native | | 0 (0) | | 0 (0) | |
| BMI: Median (IQR)) | 794 | 27.1 (23.8, 31.0) | 325 | 25.7 (22.9, 29.6) | 0.0008 |
| Ischemic History | 796 | 21 (2.6) | 334 | 23 (6.9) | 0.0008 |
| Hypertension | 796 | 422 (53.0) | 330 | 199 (60.3) | 0.0252 |
| No Hypertension | | 374 (47.0) | | 131 (39.7) | |
| Treated Hypertension | | 393 (49.4) | | 144 (43.6) | |
| Untreated Hypertension | | 29 (3.6) | | 55 (16.7) | <0.0001 |
| Diabetes | 795 | 131 (16.5) | 334 | 54 (16.2) | 0.8977 |
| High Cholesterol | 790 | 407 (51.5) | 327 | 158 (48.3) | 0.3302 |
| Smoking: | 795 | | 333 | | |
| Never | | 373 (46.9) | | 145 (43.5) | |
| Former | | 325 (40.9) | | 139 (41.7) | |
| Current | | 97 (12.2) | | 49 (14.7) | 0.4127 |
| Heavy Alcohol Use | 795 | 38 (4.8) | 319 | 18 (5.6) | 0.5513 |
| Anticoagulant Use | 796 | 51 (6.4) | 334 | 60 (18.0) | <0.0001 |
| Education: | 796 | | 332 | | |
| <=HS | | 357 (44.8) | | 217 (65.4) | |
| >HS | | 439 (55.2) | | 115 (34.6) | <0.0001 |
| MetaGRS Mean (SD): | | | | | |
| All-trait metaGRS | 796 | -0.15 (0.91) | 334 | 0.08 (0.93) | <0.0001 |

**Supplementary Table 30. Non-lobar ICH characteristics in validation dataset.**

| | Controls N=796 | | Cases N=508 | | P-value |
|---|---|---|---|---|---|
| | N Avail | N (%) | N Avail | N (%) | |
| Age: Mean(SD) | 796 | 68.4 (13.4) | 508 | 68.1 (14.4) | 0.6814 |
| Female | 796 | 400 (50.2) | 508 | 237 (46.6) | 0.2050 |
| Race: | 796 | | 508 | | 0.3896 |
| White | | 796 (100.0) | | 507 (99.8) | |
| American Indian/Alaskan Native | | 0 (0) | | 1 (0.2) | |
| BMI: Median (IQR)) | 794 | 27.1 (23.8, 31.0) | 494 | 27.4 (23.0, 32.5) | 0.6592 |
| Ischemic History | 796 | 21 (2.6) | 508 | 53 (10.4) | <0.0001 |
| Hypertension | 796 | 422 (53.0) | 502 | 363 (72.3) | <0.0001 |
| No Hypertension | | 374 (47.0) | | 139 (27.7) | |
| Treated Hypertension | | 393 (49.4) | | 259 (51.6) | |
| Untreated Hypertension | | 29 (3.6) | | 104 (20.7) | <0.0001 |
| Diabetes | 795 | 131 (16.5) | 507 | 136 (26.8) | <0.0001 |
| High Cholesterol | 790 | 407 (51.5) | 488 | 208 (42.6) | 0.0020 |
| Smoking: | 795 | | 505 | | 0.1768 |
| Never | | 373 (46.9) | | 220 (43.6) | |
| Former | | 325 (40.9) | | 206 (40.8) | |
| Current | | 97 (12.2) | | 79 (15.6) | |
| Heavy Alcohol Use | 795 | 38 (4.8) | 472 | 36 (7.6) | 0.0367 |
| Anticoagulant Use | 796 | 51 (6.4) | 508 | 83 (16.3) | <0.0001 |
| Education: | 796 | | 502 | | |
| <=HS | | 357 (44.8) | | 323 (64.3) | |
| >HS | | 439 (55.2) | | 179 (35.7) | <0.0001 |
| MetaGRS Mean (SD): | | | | | |
| All-trait metaGRS | 796 | -0.15 (0.91) | 508 | 0.18 (0.86) | <0.0001 |

**Supplementary Table 31. Significant clinical predictors of lobar ICH in validation dataset after backward elimination.**

| | **OR (95% CI)** | **P-value** |
|---|---|---|
| BMI | 0.96 (0.94, 0.98) | 0.0007 |
| History of Ischemic stroke | 2.19 (1.17, 4.10) | 0.0147 |
| Anticoagulant use | 3.12 (2.07, 4.71) | <0.0001 |
| Education: | | |
| Less than Equal HS | 2.19 (1.66, 2.88) | <0.0001 |
| Greater than HS | Reference | |

**Supplementary Table 32. Significant clinical predictors of non-lobar ICH in validation dataset after backward elimination.**

| | **OR (95% CI)** | **P-value** |
|---|---|---|
| History of Ischemic stroke | 2.61 (1.47, 4.62) | 0.0010 |
| Hypertension | 2.12 (1.61, 2.77) | <0.0001 |
| Diabetes | 1.70 (1.24, 2.33) | 0.0011 |
| High Cholesterol | 0.54 (0.42, 0.71) | <0.0001 |
| Heavy alcohol | 2.32 (1.38, 3.90) | 0.0015 |
| Anticoagulant use | 2.53 (1.67, 3.83) | <0.0001 |
| Education: | | |
| Less than Equal HS | 2.31 (1.78, 2.98) | <0.0001 |
| Greater than HS | Reference | |

**Supplementary Table 33. Associations between *APOE* genotype, metaGRS and odds of any ICH, lobar, and non-lobar ICH in the validation dataset.**

| | **OR (95% CI)** | **P-value** |
|---|---|---|
| **Any ICH** | | |
| APOE ε2 allele | 1.28 (0.99-1.66) | 0.064 |
| APOE ε4 allele | 1.34 (1.09-1.64) | 0.006 |
| metaGRS (1-SD increment) | 1.45 (1.29-1.62) | $1.3 \times 10^{-10}$ |
| APOE ε2*metaGRS | | 0.868 |
| APOE ε4*metaGRS | | 0.253 |
| **Lobar ICH** | | |
| APOE ε2 allele | 1.60 (1.16-2.21) | 0.004 |
| APOE ε4 allele | 1.74 (1.36-2.24) | $1.3 \times 10^{-5}$ |
| metaGRS (1-SD increment) | 1.36 (1.18-1.57) | $3.6 \times 10^{-5}$ |
| APOE ε2*metaGRS | | 0.452 |
| APOE ε4*metaGRS | | 0.685 |
| **Non-lobar ICH** | | |
| APOE ε2 allele | 1.08 (0.79-1.47) | 0.627 |
| APOE ε4 allele | 1.09 (0.85-1.39) | 0.511 |
| metaGRS (1-SD increment) | 1.51 (1.32-1.72) | $7.5 \times 10^{-10}$ |
| APOE ε2*metaGRS | | 0.654 |
| APOE ε4*metaGRS | | 0.138 |

All effect size estimates are adjusted for age, sex, and two principal components.
